# MRMU: A New Paradigm for Mendelian Randomization by Accounting for Measured Covariates and Unmeasured Confounders

**DOI:** 10.64898/2026.06.15.26355649

**Authors:** Xianghong Hu, Jiashun Xiao, Zhiwei Wang, Xinrui Huang, Hongyu Zhao, Can Yang

**Affiliations:** School of Mathematical Sciences, Shenzhen University, Shenzhen 518060, China; School of Public Health, Sun Yat-sen University, Guangzhou 510080, China; Department of Biostatistics, Yale School of Public Health, New Haven, CT 06520, USA; Department of Mathematics, The Hong Kong University of Science and Technology, Hong Kong SAR, China

## Abstract

Mendelian randomization (MR) is a powerful approach for causal inference, however, its reliability is frequently compromised by unadjusted covariates and unmeasured confounders, such as unmeasured pleiotropy and sample structure. To address these challenges, we introduce MRMU, a novel paradigm for the MR framework. Unlike traditional single-variable or multivariable MR methods, MRMU selects instrumental variables only from the exposure of interest and estimates one exposure effect at a time, while jointly accounting for measured covariates and unmeasured confounders. This design improves the reliability of MR analyses. In simulations and real data, MRMU achieved better type I error control, higher statistical power, and more accurate effect estimation than existing MR methods. Applying to coronary artery disease (CAD), MRMU identified robust cardiometabolic risk factors, including LDL-C, APOB, systolic blood pressure, body mass index, and smoking initiation, with consistent evidence across multiple CAD datasets. In contrast, traits such as HDL-C, height, and educational attainment, which were found to be significant by existing MR methods, were no longer supported by MRMU. MRMU further supported blood pressure-related traits, rather than lipid traits, as the more relevant pathway linking urate to CAD. Finally, by integrating large-scale plasma proteomics data, MRMU identified candidate CAD drug targets beyond established HMGCR- and PCSK9-related pathways, highlighting its utility for therapeutic target prioritization.

## Introduction

Inferring causal relationships between candidate risk factors or therapeutic targets and disease outcomes is central to understanding disease biology and informing drug discovery. Although randomized controlled trials (RCTs) provide the gold standard for causal inference [1], they are often costly, time-consuming, and sometimes infeasible or unethical [2]. Mendelian randomization (MR) offers an alternative framework for causal inference using genetic instruments, and with the increasing availability of large-scale GWAS summary statistics, has become widely used in epidemiological research [3] and drug target discovery [4, 5].

The validity of causal inferences derived from MR relies on strong assumptions on genetic instruments [6]: the genetic variant must be strongly associated with the exposure (relevance), independent of any confounders of the exposure-outcome relationship (independence), and affect the outcome exclusively through the primary exposure (exclusion restriction) [6]. A major challenge to these assumptions is pleiotropy, where genetic variants affect the outcome through pathways other than the exposure, potentially leading to false-positive causal findings [7]. Measured covariates from large-scale GWAS can help account for known pleiotropic pathways and relax the strict IV assumption that genetic instruments influence the outcome only through the primary exposure [8]. For example, when evaluating the effect of high-density lipoprotein cholesterol (HDL-C) on coronary artery disease (CAD), triglycerides (TG) should be considered as a measured covariate because many HDL-C-associated variants also influence TG [9, 10]. Beyond measured sources of bias, MR is also vulnerable to unmeasured confounders, which may arise from unmeasured pleiotropy or from sample structure embedded in GWAS summary statistics. Unmeasured pleiotropy arises when genetic variants influence the outcome through biological pathways that are not captured by the modeled exposure or the included measured covariates, generating correlations between SNP effect sizes for the exposure and outcome even in the absence of a true causal relationship[11, 12, 13]. Unmeasured confounding may also reflect sample structure and study design artifacts, including population stratification, assortative mating, sample overlap, and cryptic relatedness [14, 15, 16]. If unaccounted for, these unmeasured confounders can lead to spurious MR findings.

Existing MR methods can be broadly classified into single-variable MR (SVMR) and multivariable MR (MVMR), depending on whether measured covariates are incorporated. SVMR methods, like MR-Egger [17], MR-PRESSO [18], GSMR [19], RAPS [20], BWMR [21], MRMix [22], cML-MA [23], and CAUSE [24], attempt to address pleiotropy through assumptions on invalid instruments but do not explicitly model measured covariates. However, these models often fail in complex biomedical scenarios where major IVs are invalid due to pleiotropy. Within the SVMR framework, our previous method MRAPSS [15] improves robustness to unmeasured sources of bias by leveraging genome-wide summary statistics. Nevertheless, MRAPSS, like other SVMR methods, cannot explicitly adjust for measured covariates. In contrast, MVMR jointly models multiple related exposures [8], with extensions such as MVMR-IVW, MVMR-Egger [25], MVMR-Median, MVMR-Lasso, and MVMR-Robust [26]. However, MVMR typically uses the union of IVs across exposures, which may introduce additional invalid IVs as the number of exposures increases. Moreover, most existing methods do not explicitly account for unmeasured confounding embedded in GWAS summary statistics, such as sample structure, which can undermine the reliability of MR results.

Here, we propose MRMU as a novel paradigm for MR, aiming to overcome the limitations of SVMR and MVMR. Unlike existing MR methods, MRMU MRMU addresses this limitation by extending MRAPSS in two directions: it retains genome-wide modeling of unmeasured bias through genetic correlation and the LDSC intercept, while introducing an inferential model that explicitly incorporates measured covariates. effectively accounts for both measured covariates and unmeasured confounders through four designs. First, it estimates the causal effect of one exposure at a time, selecting IVs solely based on the exposure of interest. This approach minimizes the risk of invalid IV inclusion that can arise when instruments are aggregated across multiple exposures. Second, MRMU integrates measured covariates and genome-wide information into a unified framework. Building on our previous method, MRAPSS [15], it models unmeasured confounding while explicitly incorporating measured covariates for robust estimation of the direct causal effect. Third, MRMU facilitates the evaluation of measured covariate effects by comparing exposure effect estimates before and after covariate adjustment using the same set of IVs. This method ensures that observed changes are less likely to be influenced by variations in IV selection and can be more directly attributed to the adjusted covariates. Fourth, MRMU is robust to selection bias induced by IV selection, commonly referred to as the winner’s curse, by incorporating a likelihood conditional on the IV selection rule. These features significantly enhance the reliability and interpretability of MR analyses by reducing bias from both measured and unmeasured sources, while distinguish direct causal effects from those driven by underlying pathways.

To evaluate the performance of MRMU, we conducted comprehensive simulations and applied it to a systematic CAD analysis for causal risk factor identification and candidate drug target prioritization. In simulations, MRMU outperformed existing MR methods across diverse settings, with better type I error control, higher statistical power, and more accurate effect estimation. A real-data negative control analysis further showed that MRMU can reduce bias arising from measured covariates and unmeasured confounding. Applied to 88 traits across multiple CAD datasets, MRMU identified LDL-C, APOB, systolic blood pressure (SBP), body mass index, and smoking initiation as robust risk factors after adjustment for 22 cardiometabolic covariates. In contrast, traits such as HDL-C, height, and educational attainment, which were significant in existing MR methods, were no longer supported by MRMU after accounting for measured covariates and unmeasured confounders. Pathway-related analyses showed that the estimated effects of many traits on CAD were sensitive to adjustment for lipid or metabolic syndrome-related covariate sets, highlighting the role of shared cardiometabolic mechanisms. In particular, MRMU supported blood pressure traits, rather than lipid traits, as plausible contributors to the urate–CAD relationship. Finally, by integrating large-scale plasma proteomics data, MRMU prioritized candidate CAD drug targets, including SLC5A8, PLA2G7, C5orf38, ITM2B, and ADIPOQ, which appear to act independently of pathways represented by established lipid-lowering targets such as HMGCR and PCSK9. These findings demonstrate the utility of MRMU for robust causal inference, pathway interpretation, and drug target prioritization.

## Results

### Study overview

MRMU is an MR method that uses GWAS summary statistics to infer the causal effect of an exposure (*X*) on an outcome (*Y*) while explicitly accounting for both measured covariates 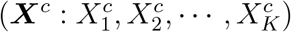 and unmeasured confounders (*U*). Figure 1-A illustrates the design of MRMU with two key features: its IV selection strategy and its modeling framework. Unlike MVMR methods that estimate causal effects from multiple exposures simultaneously, MRMU focuses on the effect of a single exposure at a time. This targeted approach enables MRMU to select IVs that are specifically aligned with the exposure of interest, thereby reducing the risk of incorporating invalid IVs. Once the IVs are identified, the MRMU probabilistic model efficiently accounts for both measured covariates and unmeasured confounders, offering a significant advantage over SVMR methods, which often ignore measured covariates. MRMU achieves its objectives through three interlinked components: (i) an inferential model for causal estimation that explicitly adjusts for measured covariates, (ii) a polygenic effect model that captures correlated pleiotropy arising from shared genetic architecture, and (iii) an estimation error model that accounts for biases induced by sample structure. Technical details of MRMU are described in the Methods section.

**Figure 1:**
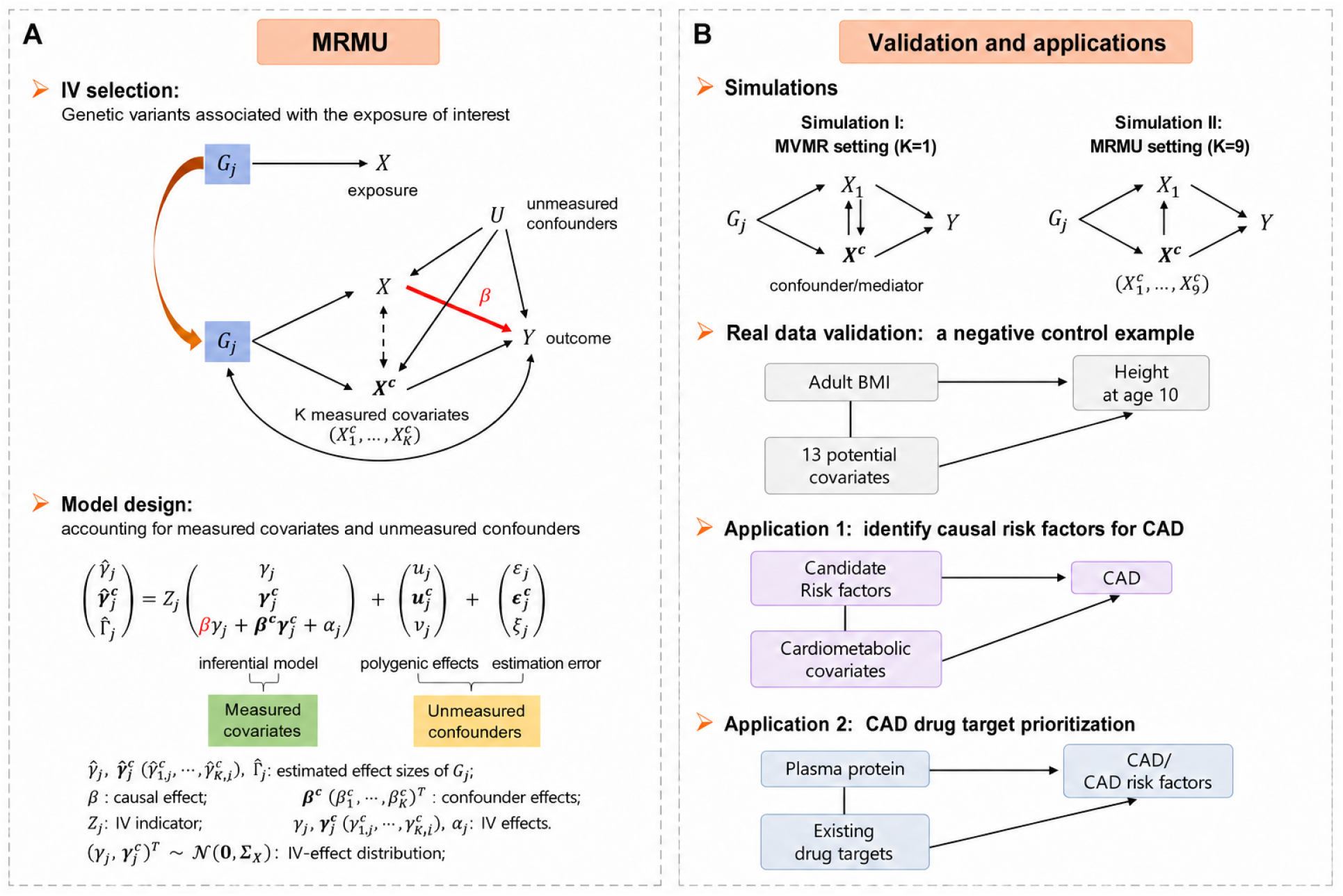
Study Overview. (**A**) Overview of the MRMU framework. IVs are selected based on their associations with the exposure of interest (*X*), and are used to estimate the causal effect (*β*) of *X* on the outcome *Y*. MRMU jointly models the exposure (*X*), measured covariates 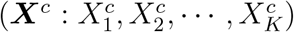, and outcome (*Y*) by incorporating three components: an inferential model for causal effect while adjusting for measured covariates, a polygenic effect model and an estimation error model for unmeasured confounders (unmeasured pleiotropy and sample structure). Within the inferential model, MRMU assumes that the IV effect on the exposure (*γ*_*j*_) and the IV effects on the measured covariates 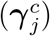 follow a multivariate normal distribution with variance–covariance matrix **Σ**_*X*_. From **Σ**_*X*_, MRMU derives two IV-level quantities to describe pleiotropic overlap between the exposure and measured covariates: the variance ratio and the IV-effect correlation. For the *k*-th measured covariate, the variance ratio 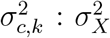 measures the relative magnitude of IV effects on 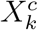 compared with *X*, whereas the IV-effect correlation measures the extent of pleiotropic overlap between *γ*_*j*_ and 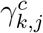. These quantities assess the extent to which exposure-selected IVs also capture measured covariate signals. Larger variance ratios or stronger IV-effect correlations indicate greater IV-level pleiotropic overlap with the corresponding covariates, providing support for including measured covariates in the MRMU framework. Formal definitions of these two quantities are provided in the Methods section. (**B**) MRMU was evaluated through simulations under different model settings and a real-data negative control analysis, and was further applied to coronary artery disease (CAD) analyses for causal risk factor identification and drug target prioritization.

The effectiveness of MRMU was comprehensively evaluated through both simulations and real data applications (Figure 1-B). Specifically, we conducted simulations to evaluate the calibration, power, and estimation accuracy of MRMU in the presence of measured covariates and unmeasured confounders. Simulation I considered an MVMR setting with one measured covariate (*K* = 1), examining cases where the covariate acted either as a confounder or as a mediator. Simulation II considered an MRMU setting with nine measured confounders (*K* = 9), allowing us to assess the impact of including both true and null confounders. Details of the simulation setting are provided in the supplementary note 1.4. We then used a real-data negative control example, with adult BMI as the exposure and height at age 10 as the outcome, to evaluate whether MRMU can control false-positive findings in a setting where a direct causal effect is biologically implausible. In real applications, we applied MRMU to identify causal risk factors for CAD among 88 traits while accounting for cardiometabolic covariates, including blood and urate biomarkers, lifestyle factors, and metabolic syndrome components such as body mass index (BMI), systolic blood pressure (SBP), and diastolic blood pressure (DBP). To evaluate replicability, we analyzed three CAD datasets. Finally, we integrated large-scale plasma proteomics data to prioritize drug targets that may act directly on CAD or its risk factors, beyond pathways targeted by existing CAD therapies. To facilitate wider adoption and reproducibility, we have implemented MRMU in an efficient and freely available R package, accessible at https://github.com/hxh0928/MRMU. The data and code required to reproduce the real-data applications are also publicly available on GitHub.

### Simulations assessing calibration, statistical power, and point estimate

In Simulation I, we considered a setting with one measured covariate (*K* = 1), acting either as a confounder or as a mediator, and compared MRMU with six SVMR methods (IVW, Egger, RAPS, CAUSE, Weighted-mode, and Weighted-median) and five MVMR methods (MVMR-IVW, MVMR-Egger, MVMR-Median, MVMR-Lasso, and MVMR-Robust) in terms of type I error control, statistical power, and causal effect estimation. To assess type I error control, we generated quantile–quantile plots under the null hypothesis (*β* = 0), as shown in Figures 2A and 2B. In the confounder setting, MRMU, Egger, CAUSE, and MVMR-Egger produced well-calibrated *p*-values and effectively controlled type I error. For power comparisons, we focused on MRMU, Egger, CAUSE, and MVMR-Egger, as the other methods tended to produce inflated false postives. As shown in Figure 2C, MRMU achieved the highest power among these methods. These results suggest that many existing SVMR methods struggle when a substantial proportion of IVs are invalid due to confounding, whereas many current MVMR methods remain susceptible to unmeasured confounding. In contrast, MRMU jointly accounts for measured covariates and unmeasured confounders and leverages a large number of pleiotropic variants with moderate instrument strength to improve causal inference. As a result, MRMU can better avoid false positives induced by confounding while maintaining higher statistical power. Results under the mediator setting showed a similar overall pattern and are presented in the Fig S2A-C.

**Figure 2:**
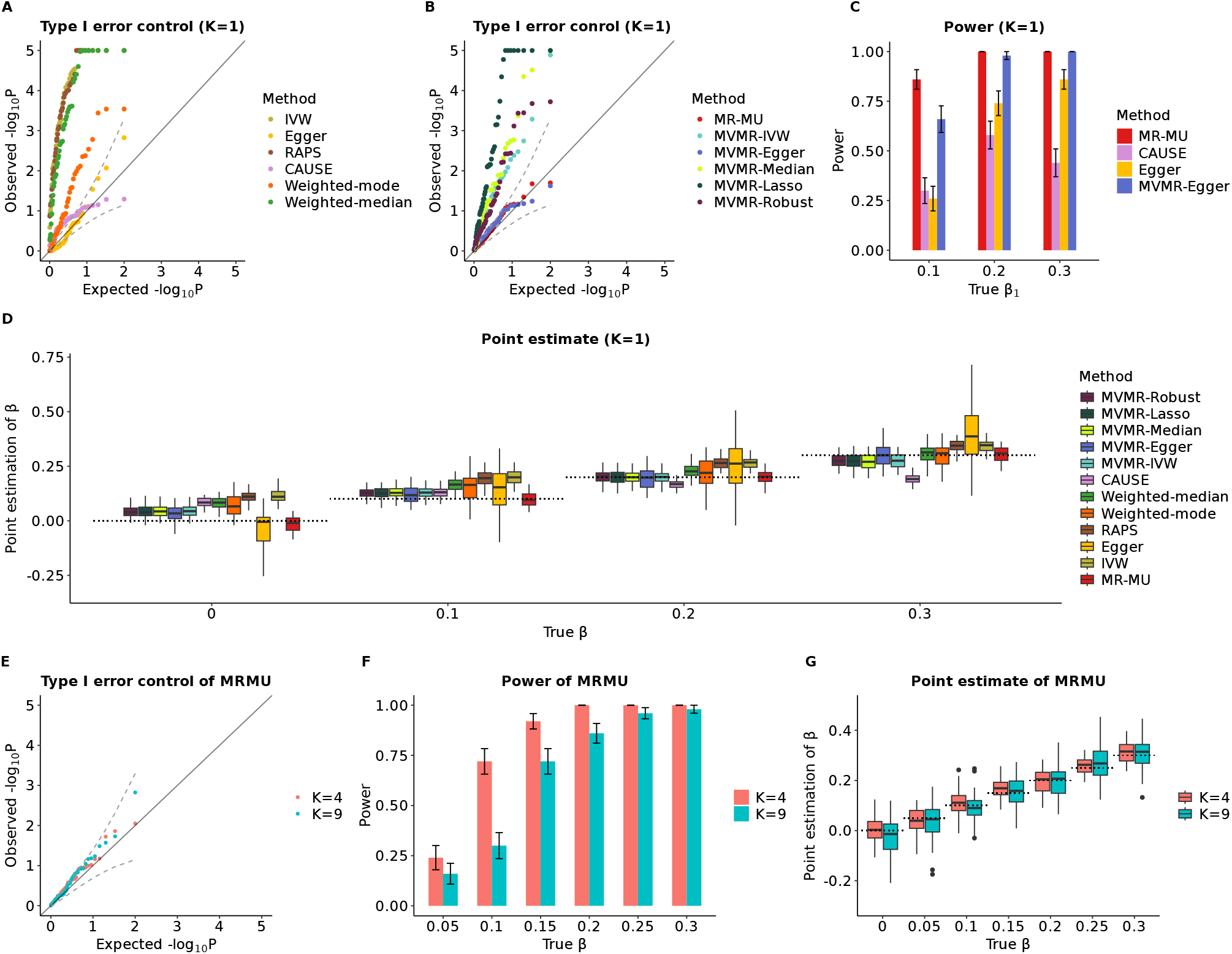
Performance of MRMU and compared SVMR and MVMR methods in simulated data. (**A**–**D**) Results under the simulation setting with one measured confounder (*K* = 1). (**A, B**) Quantile–quantile plots of − log_10_(*P*) values under the null setting (*β* = 0) for MRMU and SVMR methods (**A**) and for MRMU and MVMR methods (**B**). MRMU, MR-Egger, CAUSE, and MVMR-Egger showed well-calibrated type I error. (**C**) Statistical power of MRMU, MR-Egger, CAUSE, and MVMR-Egger under alternative settings with *β* ∈ {0.1, 0.2, 0.3}. (**D**) Causal effect estimates across methods under settings with *β* ∈ {0, 0.1, 0.2, 0.3}; dotted horizontal lines indicate the true causal effects. (**E**–**G**) Performance of MRMU in settings with multiple measured covariates (*K* = 4 or *K* = 9). (**E**) Quantile–quantile plots of − log_10_(*P*) values under the null setting (*β* = 0). (**F**) Statistical power of MRMU under alternative settings with *β* ranging from 0.05 to 0.3. (**G**) Causal effect estimates from MRMU under settings with *β* ranging from 0 to 0.3; Results for each setting were summarized from 50 replications.

We next evaluated the accuracy of point estimation in Simulation I. In the confounder setting, MRMU provided more accurate effect estimates than the compared methods (Figures 2D). Although Egger and MVMR-Egger yielded less biased estimates, they showed substantially larger estimation errors. The improved performance of MRMU can be attributed to two key features. First, by accounting for both measured covariates and unmeasured confounders, MRMU reduces bias arising from confounding and thereby improves estimation accuracy. Second, MRMU addresses the winner’s curse, or selection bias, which arises when the same summary statistics are used for both IV selection and effect estimation. Specifically, MRMU uses a conditional likelihood approach to alleviate this bias, leading to more reliable effect estimates. Among the compared methods, CAUSE appeared particularly vulnerable to selection bias because it includes weaker instruments using a relatively liberal IV selection threshold of 1 × 10^−3^. Similar findings for the mediator setting are provided in the Supplementary Fig S2D. We extended simulation I by setting the causal effect of the confounder on the outcome to zero (i.e., *β*^*c*^ = 0). In this specific scenario, the presence of the confounder does not introduce confounding effect in the MR testing between the exposure and outcome. Nevertheless, unmeasured confounders remain influential and must be taken into account. We observed that MRMU consistently demonstrated satisfactory performance in terms of well-calibrated p-values, higher statistical power, and accurate estimation (see Fig. S3). However, most SVMR methods still exhibited inflated p-values and biased causal estimates, as did the MVMR methods. It is worth noting that when only unmeasured confounders were relevant, the extent of p-value inflation and bias in causal estimates from SVMR methods decreased. Combined with the previous simulation results, our findings highlight the critical importance of considering both measured and unmeasured confounders in MR analysis to obtain accurate estimates and draw meaningful conclusions in MR studies.

In Simulation II, we evaluated the performance of MRMU in the presence of multiple measured confounders. We considered a setting with nine measured confounders, including four true confounders and five null confounders with no causal effect on the outcome. We examined two cases: one where MRMU accounted for all nine measured confounders (*K* = 9), and another where it accounted for only the four true confounders (*K* = 4), allowing us to assess the impact of including null confounders. The summarized results of MRMU for both cases are presented in Figs. 2E-G. The results show that MRMU effectively controls type I errors(2 E), achieves satisfactory power (Fig. 2 F), and demonstrates high estimation accuracy in both cases (Fig. 2G). We observed a slight reduction in the power of MRMU in the case of *K* = 9 where null confounders are involved (Fig. 2 B). This highlights the importance of carefully selecting confounders and avoiding the inclusion of too many irrelevant ones in MRMU to improve statistical power and estimation efficiency. We also compared the performance of MRMU with SVMR methods (Fig. S4). Among the compared methods, MRMU and Egger produced well-calibrated p-values, indicating their ability to effectively control type I errors. However, MRMU exhibited higher statistical power and estimation precision compared to Egger. CAUSE, which had previously shown no type I error inflation in Simulation I, exhibited a mild inflation in Simulation II. Other SVMR methods, such as IVW, RAPS, weighted-mode, and weighted-median, demonstrated inflated type I error rates.

### MRMU distinguishes direct effects from pathway-driven effects

In this section, we aim to highlight two key points. First, MRMU can recover the direct causal effect of the exposure on the outcome after adjusting for measured covariates included in the model. Second, comparing MRMU results obtained with and without adjustment for a measured covariate can help clarify the role of measured covariates in shaping exposure–outcome relationships. Notably, MRMU without adjustment for a measured covariate is equivalent to MRAPSS, in which no *X*^*c*^ is included.

To illustrate these points, we performed analysis under the setting of Simulation I by comparing MRMU results with and without adjustment for the measured covariate under two scenarios, in which *X*^*c*^ acted either as a confounder or as a mediator. We considered both the null setting for the causal effect (*β* = 0) and the alternative setting (*β* = 0.2), while allowing the covariate effect to be nonzero (*β*^*c*^≠ 0). When *X*^*c*^ acted as a confounder, the unadjusted analysis, MRAPSS, produced larger effect estimates than the adjusted MRMU analysis (Fig.3A-B). Under the null setting, failure to adjust for *X*^*c*^ induced an apparent nonzero effect, whereas the adjusted MRMU analysis remained centered around the null (Fig.3A). Under the alternative setting, the MRMU analysis recovered an effect close to the true effect, while the MR-APSS again yielded inflated estimates (Fig.3B). These results indicate that, when *X*^*c*^ is a confounder, the discrepancy between MR-APSS and MRMU reflects confounding bias, and adjustment for *X*^*c*^ enables MRMU to recover the unbiased causal effect of *X* on *Y*. When *X*^*c*^ acted as a mediator, MR-APSS again produced larger effect estimates than the adjusted MRMU analysis (Fig.3C-D). Under the null setting, the MR-APSS showed an apparent nonzero effect, whereas the MRMU analysis remained centered around the null (Fig.3C). Under the alternative setting, the MRMU analysis recovered an effect close to the target value, while the MR-APSS analysis yielded a larger estimate (Fig.3D). In this setting, however, the discrepancy between the MR-APSS and MRMU analyses has a different interpretation: the MR-APSS analysis captures the total effect of *X* on *Y* (the sum of the direct effect and the indirect effect mediated by *X*^*c*^), whereas the MRMU analysis recovers the direct effect conditional on the mediator. Thus, the difference between the two analyses reflects the contribution of the indirect pathway through *X*^*c*^. Our results show that MRMU can recover the true (direct)causal effect after accounting for measured covariates, and that comparing MRMU and MR-APSS can help reveal whether an observed effect between exposure and outcome is driven by confounding or by a mediated biological pathway.

**Figure 3:**
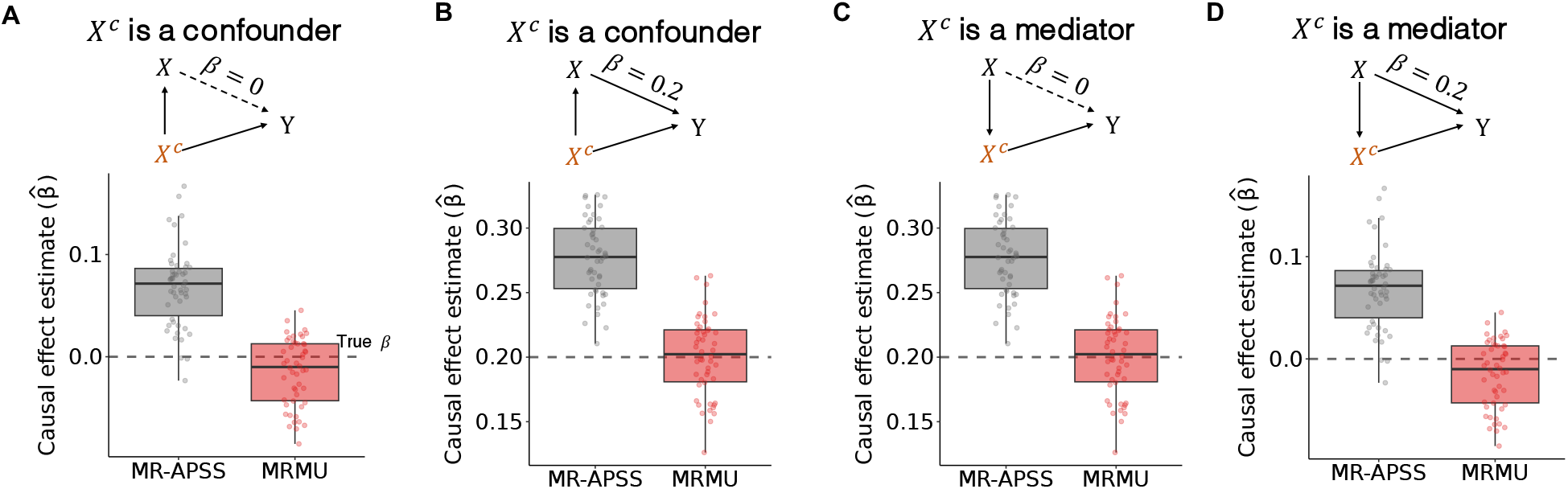
Simulations comparing MRMU with MR-APSS. (**A**)-(**B**) consider the measured covaraite *X*^*c*^ as a confounder, and (**C**)-(**D**) consider *X*^*c*^ as a mediator. In each case, the null setting (*β* = 0) and the alternative setting (*β* = 0.2) with *β*^*c*^≠ 0 are shown.

### Performance validation using a real-data negative control example

To validate MRMU in a real-world setting, we conducted a negative control analysis using adult body mass index (adult BMI) as the exposure and height at age 10 as the outcome. Because height at age 10 is measured before adult BMI, a direct causal effect of adult BMI on childhood height is biologically implausible. However, adult BMI-associated genetic variants may be linked to childhood height through growth, developmental, socioeconomic, dietary, or lifestyle-related pathways. This setting provides a challenging negative control example in which spurious causal signals may arise from measured pleiotropic pathways or unmeasured confounding rather than a true direct causal effect.

We analyzed GWAS summary statistics for adult BMI, height at age 10, and 13 candidate measured covariates representing potential pleiotropic pathways. These covariates covered socioeconomic, diet-related, pubertal timing, reproductive, and lifestyle-related domains, and were selected to capture plausible measured pathways linking adult BMI-associated variants to childhood height (see Supplementary Note 1.5 for details). The GWAS sources are summarized in Supplementary Table S2. MRMU was then used to estimate the direct effect of adult BMI on height at age 10 while accounting for both measured covariates and unmeasured confounders. We compared MRMU with seven SVMR methods and five MVMR methods. Method-specific IV selection thresholds and LD clumping procedures are provided in Supplementary Note 1.4.2 and summarized in Supplementary Table S1.

We first assessed the potential influence of unmeasured confounding using genetic correlations and LDSC intercepts. The genetic correlation between adult BMI and height at age 10 was not significant at the nominal level of 0.05, consistent with the negative control design (Figs. 4A–B). In contrast, substantial genetic correlations and non-zero LDSC intercepts were observed between adult BMI or height at age 10 and the measured covariates, and most covariate pairs also showed evidence of correlated genetic effects and sample-structure-related inflation. These results indicate that the surrounding covariate network contains substantial pleiotropic and sample-structure signals, making this a useful setting for evaluating whether MR methods can distinguish a temporally implausible direct effect from confounding-driven associations. We next examined the extent to which adult BMI instruments overlapped with the measured covariates. Under the MRMU inferential model, we estimated the variance–covariance matrix of IV effects on the exposure and measured covariates, and derived the variance ratio and correlation between IV–covariate and IV–exposure effects. The variance ratio is the estimated variance of IV effects on a measured covariate divided by the estimated variance of IV effects on adult BMI, so a larger value indicates that adult BMI-selected IVs also have stronger effects on that covariate. The IV-effect correlation measures the extent of pleiotropic overlap between IV effects on adult BMI and IV effects on a given measured covariate. As shown in Fig. 4C, the variance ratios ranged from 0.055 to 0.435, with an average of 0.147, and IV effects on adult BMI were correlated with IV effects on most measured covariates. These results indicate that adult BMI instruments were not exclusively associated with the exposure, but also captured substantial measured pleiotropic signals. This supports the use of measured covariate adjustment in MRMU and helps explain why methods that do not jointly account for these covariate signals and unmeasured confounding may produce spurious effects in this negative control example.

**Figure 4:**
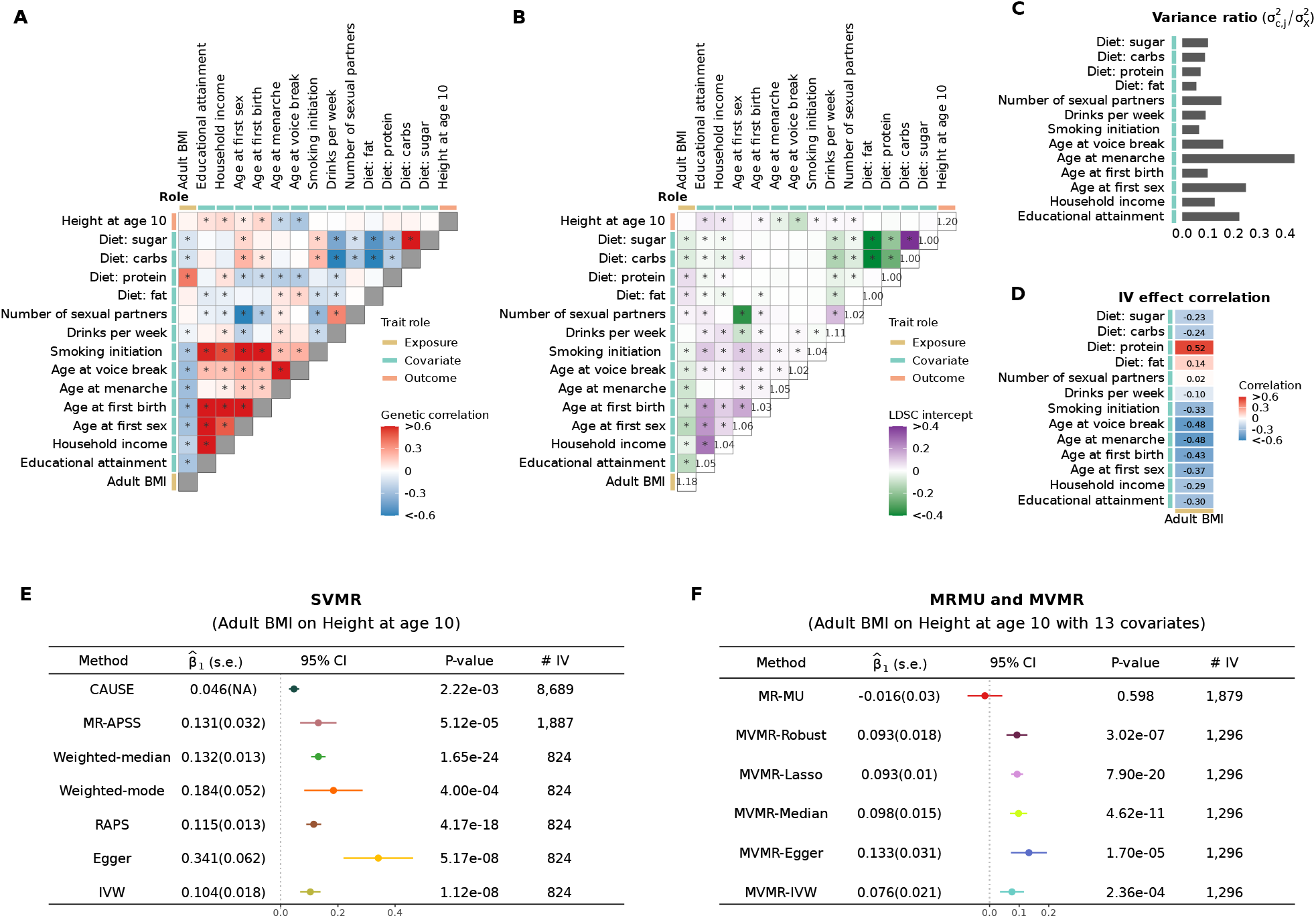
Real-data negative control analysis using adult BMI as the exposure and height at age 10 as the outcome. (**A**) Genetic correlation estimates among adult BMI, 13 measured covariates, and height at age 10. (**B**) LDSC intercept estimates among adult BMI, 13 measured covariates, and height at age 10. (**C**) Variance ratio of IV effects on each measured covariate relative to IV effects on adult BMI. A larger value indicates stronger IV-covariate effects among adult BMI-selected IVs. (**D**) Correlation between IV effects on adult BMI and IV effects on each measured covariate, reflecting the extent of IV-level pleiotropic overlap. (**E**) Causal effect estimates from seven SVMR methods. (**F**) Causal effect estimates from MRMU and five MVMR methods.

Accounting for both measured covariates and unmeasured confounders, MRMU did not support a direct causal effect of adult BMI on height at age 10 (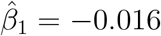, s.e. = 0.030), consistent with the temporal negative control design (Fig. 4F). In contrast, all competing SVMR and MVMR methods reported significant positive effects (Figs. 4E–F). For example, IVW estimated 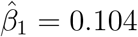(s.e. = 0.018), while MR-APSS, which accounts for unmeasured confounding only, and MVMR-IVW, which adjusts for measured covariates only, both yielded positive estimates of 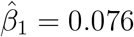(s.e. = 0.021). These results suggest that adjusting for either measured covariates or unmeasured confounding alone may be insufficient in this negative control setting. By jointly modeling both sources of bias, MRMU reduced the spurious positive association and produced results consistent with the expected absence of a direct causal effect.

### MRMU identifies robust cardiometabolic risk factors for CAD

We applied MRMU to investigate potential causal risk factors for coronary artery disease (CAD). We collected 88 complex traits as candidate risk factors and broadly classified them into two groups: cardiometabolic (52 traits) and non-cardiometabolic traits (36 traits). The cardiometabolic group included metabolic syndrome-related traits, blood and urate biomarkers, and lifestyle-related factors, whereas the non-cardiometabolic group comprised traits from other domains, including socioeconomic, reproductive, anthropometric, mental health, gastrointestinal, autoimmune, cancer, blood cell, and other disease-related traits. For the outcome, we used CAD GWAS summary statistics from the UK Biobank in the discovery stage. Detailed information on the GWAS sources for all traits is provided in Supplementary Table S3. After collecting GWAS summary statistics for the candidate risk factors and CAD, we performed standard preprocessing and harmonization across datasets. We then prescreened the candidate traits using MR-APSS and retained 37 risk factors as exposures (25 cardiometabolic traits and 12 non-cardiometabolic traits) with a test P value less than 0.05.

We included the 25 prescreened cardiometabolic traits as measured covariates to help distinguish independent causal effects from associations driven by shared cardiometabolic mechanisms. MRMU was then applied to these prescreened traits while adjusting for the measured cardiometabolic covariates. The robustness of the identified associations was further evaluated in two independent CAD GWAS datasets, including CARDIoGRAMplusC4D and a larger CAD meta-analysis. The analytical workflow is illustrated in Figure S5 and described in the Methods section. To minimize the risk of including irrelevant measured covariates in MRMU, prescreened cardiometabolic traits showing weak pleiotropic evidence with the instruments of a given exposure were excluded from the candidate covariate set. Specifically, for each exposure, IVs were first selected from the exposure GWAS, and each candidate covariate was then evaluated by counting how many of these exposure-selected IVs reached genome-wide significance (*P <* 5 × 10^−8^) in the corresponding covariate GWAS. Covariates with fewer than five such variants were excluded, as they provided limited evidence of pleiotropic relevance for adjustment. When LDL-C is used as the exposure, APOB was further removed from the covariate set because of its high correlation with LDL-C 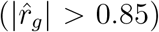. The final measured covariate set used for each exposure in MRMU is shown in Supplementary Figure S6. We compared the results from MRMU with those from existing SVMR and MVMR methods. To ensure a fair comparison with MRMU, all cardiometabolic traits were jointly included in the MVMR model, so that their effects were estimated conditional on one another. For non-cardiometabolic traits, each trait was added one at a time to this cardiometabolic set, and its effect was estimated conditional on the jointly modeled cardiometabolic traits

As shown in Fig. 5A, MRMU identified a relatively focused set of causal risk factors for CAD, all of which were cardiometabolic traits. In particular, systolic blood pressure (SBP), body mass index (BMI), low-density lipoprotein cholesterol (LDL-C), apolipoprotein B (APOB), and Smoking initiation showed positive effects on CAD (UKBB) which are significant after Bonferroni correction in the discovery stage (red colored in 5A). Among them, SBP, BMI, LDL-C, and APOB exhibited strong and highly consistent evidence across all CAD datasets. For SBP, the estimated effects were 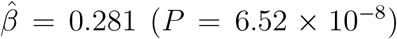 in CAD (UKB), 0.275 (*P* = 7.40 × 10^−7^) in CAD (2015), and 0.258 (*P* = 1.79 × 10^−7^) in CAD (meta). For BMI, the corresponding estimates were 0.131 (*P* = 4.22 × 10^−4^), 0.106 (*P* = 0.011), and 0.108 (*P* = 6.88 × 10^−4^). LDL-C showed highly consistent and strong effects across datasets, with estimates of 0.189 (*P* = 4.87 × 10^−12^), 0.176 (*P* = 1.88 × 10^−9^), and 0.187 (*P* = 7.67 × 10^−14^), whereas APOB yielded estimates of 0.132 (*P* = 6.57 × 10^−7^), 0.174 (*P* = 3.95 × 10^−8^), and 0.147 (*P* = 1.15 × 10^−7^). Smoking initiation showed a positive effect in CAD (UKB) and CAD (meta). MRMU not only recovered several cardiometabolic determinants of CAD but also prioritized those with strong replication across multiple CAD datasets, demonstrating its ability to distinguish robust independent effects from a larger set of potentially pathway-driven signals. The traits prioritized by MRMU span four major CAD-related domains: blood pressure regulation, lipid/apolipoprotein metabolism, adiposity, and smoking behavior, a pattern consistent with independent clinical and epidemiological evidence linking these pathways to CAD risk. Blood-pressure lowering reduces major cardiovascular events and cardiovascular mortality [27]. LDL-C lowering in randomized statin trials reduces major vascular events, supporting the clinical relevance of LDL-C and apoB-containing lipoprotein pathways [28, 29]. Obesity, particularly abdominal obesity, has been identified as an important modifiable risk factor for myocardial infarction, and weight-loss pharmacotherapy reduces cardiovascular events in individuals with overweight or obesity and established cardiovascular disease [30, 31, 32]. Smoking initiation is also consistent with large epidemiological evidence identifying smoking behavior as a major modifiable risk factor for myocardial infarction, coronary heart disease, cardiovascular mortality, and premature death [30, 33]. T2D and HbA1c also showed glycemic-related signals in the CAD analysis. T2D showed nominal evidence in the discovery dataset and stronger evidence in CAD (2015), with a positive effect also observed in CAD (meta). HbA1c showed modest but directionally consistent positive effects across the three CAD datasets. These glycemic-related signals are consistent with large prospective evidence showing that diabetes confers about a two-fold excess risk for vascular diseases, including coronary heart disease, and with cohort evidence linking higher HbA1c levels to increased cardiovascular disease and mortality risk [34, 35]. The positive T2D and HbA1c estimates after cardiometabolic covariate adjustment suggest that glycemic dysregulation may contribute to CAD risk, while also indicating that part of this relationship may overlap with broader cardiometabolic pathways

**Figure 5:**
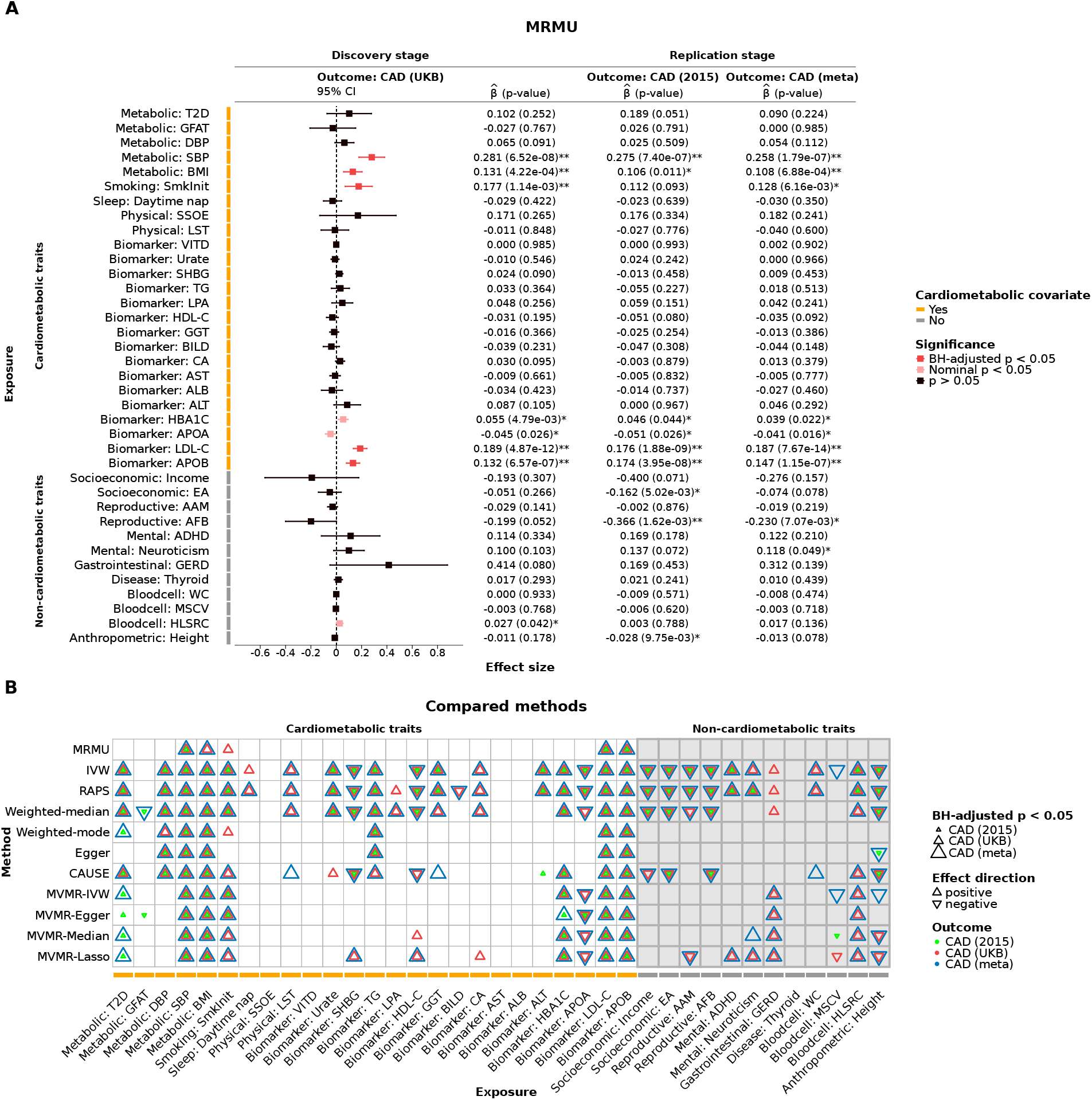
Identification of causal risk factors for CAD. (**A**) MRMU estimates for the 37 prescreened traits across three CAD datasets, including the discovery dataset CAD (UKB) and two replication datasets, CAD (2015) and CAD (meta). Point estimates and 95% confidence intervals are shown for the discovery stage, with corresponding effect estimates and *p*-values listed for all three datasets. Traits are grouped into cardiometabolic and non-cardiometabolic categories. Orange labels indicate exposures for which cardiometabolic covariates were included in the adjustment set. Red points denote significance after Bonferroni correction (*P <* 0.05*/*37) in the discovery stage, pink points denote nominal significance (*P <* 0.05), and black points denote non-significant results. (**B**) Comparison of significant findings from MRMU, SVMR methods, and MVMR methods across the 37 prescreened traits. Symbols indicate significant effects after Bonferroni correction (*P <* 0.05*/*37), with symbol orientation indicating effect direction and color indicating the CAD dataset in which the effect was detected.

Figure 5B compares significant findings from MRMU with those from competing SVMR and MVMR methods. Conventional SVMR methods, including IVW, RAPS, weighted median, weighted mode, MR-Egger, and CAUSE, identified a broader set of Bonferroni-significant associations with CAD across both cardiometabolic and non-cardiometabolic traits. Within the cardiometabolic group, these methods frequently implicated calcium (CA), alanine aminotransferase (ALT), diastolic blood pressure (DBP), high-density lipoprotein cholesterol (HDL-C), triglycerides (TG), urate, apolipoprotein A (APOA), sex hormone-binding globulin (SHBG), and leisure screen time (LST), with significant effects detected in at least one CAD dataset. These associations were not supported by MRMU after adjustment for measured cardiometabolic covariates and unmeasured confounders. MVMR methods showed a more limited set of significant findings than SVMR methods, but several traits, including apolipoprotein A (APOA), sex hormone-binding globulin (SHBG), high-density lipoprotein cholesterol (HDL-C), and calcium (CA), remained significant in at least one MVMR analysis. Compared with MVMR methods, MRMU reported fewer Bonferroni-significant associations among cardiometabolic traits. The contrast between MRMU and competing methods was even more pronounced for non-cardiometabolic traits. Across the three CAD datasets, MRMU did not provide Bonferroni-significant evidence for direct effects of any non-cardiometabolic trait after adjustment for cardiometabolic covariates. In contrast, several SVMR methods reported significant associations for educational attainment (EA), age at menarche (AAM), age at first birth (AFB), attention-deficit/hyperactivity disorder (ADHD), neuroticism, high light scatter reticulocyte count (HLSRC), height, gastroesophageal reflux disease (GERD), and thyroid disease. Some MVMR methods also retained a subset of these signals, including GERD, HLSRC, AFB, ADHD, neuroticism, and height. However, these non-cardiometabolic signals detected by competing methods showed limited consistency across CAD datasets and across methods.

To better understand why competing methods produced a broader set of significant findings, we further conducted a sensitivity analysis comparing MRMU with its variants that adjusted for measured covariates only or unmeasured confounding only. This comparison was motivated by the different sources of bias addressed by existing methods: SVMR methods estimate one exposure at a time and generally do not condition on measured covariates, whereas MVMR methods adjust for measured covariates but do not explicitly model unmeasured confounding in GWAS summary statistics, such as sample structure. Therefore, removing one adjustment component at a time provides a direct way to assess whether the broader signals detected by SVMR and MVMR methods are sensitive to measured pathway adjustment, unmeasured-confounding adjustment, or their joint modeling. The unmeasured-confounding-only variant was implemented using MR-APSS, whereas the measured-covariate-only variant was implemented in MRMU by setting **Ω** = **0** and ***C*** = ***I***. The results are summarized in Supplementary Figure S7. The unmeasured-confounding-only variant produced a broader set of significant findings, resembling the pattern observed for SVMR methods. Specifically, 19 traits remained significant after Bonferroni correction in this variant, indicating that accounting for unmeasured confounding alone may be insufficient when measured cardiometabolic pathways are present. In contrast, the measured-covariate-only variant attenuated many associations after adjustment for cardiometabolic covariates, with 8 traits remaining significant after Bonferroni correction. Several non-cardiometabolic traits, including height, neuroticism, and educational attainment (EA), remained significant in both variants but were no longer significant in the full MRMU model. This pattern suggests that the broader signals detected by SVMR and MVMR methods may arise, at least in part, from incomplete adjustment for measured pathways, unmeasured confounding, or both. By jointly accounting for measured covariates and unmeasured confounders within an exposure-specific IV framework, MRMU may reduce pathway-driven and confounding-induced signals.

As a further illustration, HDL-C provides a representative example of a signal detected by several competing methods but not supported by the full MRMU model. In the sensitivity analysis, the unmeasured-confounding-only variant estimated a strong negative association between HDL-C and CAD 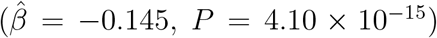, and the measured-covariate-only variant also retained a nominally significant association 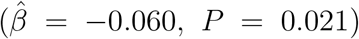 (Supplementary Figure S7). After jointly accounting for measured covariates and unmeasured confounders, the HDL-C estimate was further attenuated and became non-significant in the full MRMU model 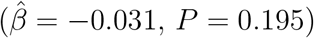. This result suggests that the HDL-C signal detected by several competing methods may be sensitive to both measured pathway adjustment and unmeasured-confounding adjustment.This pattern is consistent with prior evidence suggesting that HDL-C concentration itself is unlikely to be a robust independent causal determinant of CAD. In a classical Mendelian randomization study, Voight et al. used both a functional LIPG variant and a genetic score composed of variants selected to be specifically associated with HDL-C, but genetically increased HDL-C was not associated with lower CAD risk [36]. Beyond genetic evidence, randomized trials and meta-analyses have shown that HDL-targeted treatments, including niacin and CETP inhibitors, did not reduce major cardiovascular outcomes despite effectively raising HDL-C levels [37, 38, 39]. These findings further support the interpretation that HDL-C associations detected by several competing methods may reflect shared lipid pathways or unmeasured confounding rather than a direct HDL-C effect.

### MRMU characterizes pathway-related covariate dependence in exposure–CAD relationships

We applied MRMU to examine how three biologically motivated covariate subgroups affected the estimated effects of the 37 prescreened traits on CAD. These covariate sets included lipid traits (APOB, APOA, LDL-C, HDL-C, and TG), metabolic syndrome–related traits (BMI, SBP, DBP, T2D), and lifestyle factors (daytime napping, SSOE, LST, and smoking initiation). These three covariate groups were chosen because they represent major biologically interpretable pathways relevant to CAD: lipid metabolism and atherogenic lipoprotein pathways, metabolic syndrome– related processes involving adiposity, blood pressure, and glycemic regulation, and behavioral or lifestyle-related pathways [30, 29, 27]. They also correspond to the main cardiometabolic domains highlighted in the preceding CAD risk factor analysis, allowing us to evaluate whether the estimated effects of prescreened traits on CAD were sensitive to different classes of measured covariate adjustment. We then compared covariate-adjusted MRMU estimates with MR-APSS estimates. This comparison helps identify exposure–CAD relationships whose estimated effects are sensitive to specific measured covariates, while reducing the possibility that the observed differences are driven by changes in IV selection. A “defined difference” was recorded when the covariate-adjusted MRMU estimate differed significantly from the MR-APSS estimate, or when an association changed from significant to non-significant after MRMU adjustment. These exposure–CAD pairs are marked by † in Fig. 6A–C. The detailed definition and testing procedure for a defined difference are provided in Supplementary Note 1.7.

**Figure 6:**
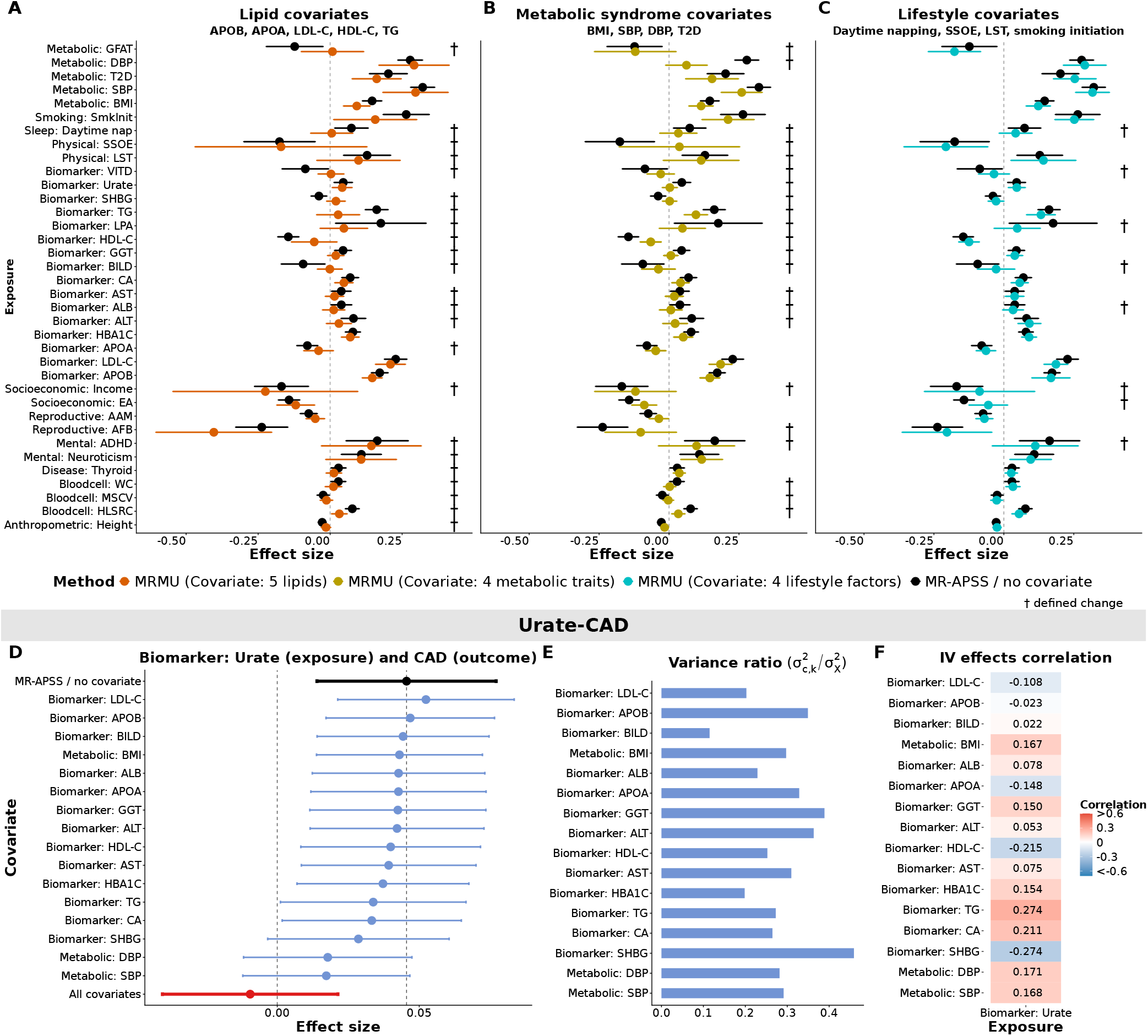
MRMU characterizes the impact of different covariate sets on exposure– CAD relationships. (**A**)–(**C**) Effect estimates from MR-APSS with those from MRMU after separate adjustment for three covariate sets: lipid traits (**A**; APOB, APOA, LDL-C, HDL-C, and TG), metabolic syndrome–related traits (**B**; BMI, SBP, DBP, and T2D), and lifestyle factors (**C**; daytime napping, SSOE, LST, and smoking initiation). Each point and error bar represents the estimated effect and confidence interval for one exposure–CAD pair. (**D**)–(**F**) The urate–CAD relationship case: **D** Causal effect estimates after adjustment for one covariate at a time and for all covariates jointly, (**E**) The variance ratio of IV effects on each covariate relative to urate, and (**F**) The IV-effect correlation between urate and each covariate.

Among the 37 prescreened exposures, 23, 22, and 8 showed a defined difference after adjustment for lipid traits, metabolic syndrome-related traits, and lifestyle factors, respectively. This pattern indicates that measured covariate adjustment can substantially alter the estimated effects of prescreened traits on CAD. In particular, lipid and metabolic syndrome-related covariates affected a larger number of estimated effects than lifestyle covariates, suggesting that these two covariate groups captured broader covariate-dependent variation in this CAD analysis. Our analysis further revealed exposure-specific patterns of covariate dependence. Height, APOA, neuroticism, and thyroid disease showed defined differences only after adjustment for lipid traits, suggesting that their estimated effects on CAD were primarily sensitive to lipid-related covariates. In contrast, EA and daytime napping were more clearly altered after adjustment for lifestyle factors. These trait-specific patterns illustrate that MRMU can help identify which classes of measured covariates are most relevant to the estimated effect of each trait on CAD.

The urate–CAD relationship provides a specific example of how MRMU can characterize covariate-dependent patterns in estimated exposure effects. In the subgroup analysis, a defined difference was observed only after adjustment for the metabolic syndrome–related covariate set, whereas little change was seen after adjustment for the lipid or lifestyle covariate sets. We therefore performed a one-covariate-at-a-time analysis to localize this covariate dependence and identify which measured covariates contributed most to the change. The clearest blood pressure-related changes in the estimated urate effect on CAD were observed after adjustment for SBP and DBP (Fig. 6D), suggesting that blood pressure-related covariates may be particularly relevant to the urate–CAD relationship. To further interpret this blood pressure-related pattern, we examined IV-level signals using MRMU. We selected 698 IVs for urate using an IV threshold of *P <* 5 × 10^−8^. Under the MRMU inferential model, we estimated the variance–covariance matrix of IV effects on urate and the measured covariates, from which we derived the variance ratio and the IV-effect correlation. These metrics quantify pleiotropic overlap between urate-associated IVs and measured covariates. Urate-associated IVs showed substantial overlap with both SBP and DBP, with variance ratios close to 0.3 (Fig. 6E) and IV-effect correlations of 0.168 for SBP and 0.171 for DBP (Fig. 6F). These findings suggest that urate-associated IVs also capture blood pressure-related pleiotropy signals. These blood pressure-related findings are consistent with existing evidence linking urate to blood pressure regulation. In a randomized trial, allopurinol lowered blood pressure in hyperuricemic adolescents with newly diagnosed hypertension, supporting a potential causal effect of urate on blood pressure [40]. Additional genetic and clinical evidence has suggested that blood pressure may account for part of the relationship between urate and cardiovascular disease risk [41, 42]. Consistent with this evidence, our results support blood pressure-related pathways as a plausible mediator of the urate–CAD relationship.

### MRMU prioritizes candidate CAD protein targets beyond established lipid-lowering pathways

We applied MRMU to prioritize candidate protein targets for CAD using large-scale plasma proteomics GWAS summary statistics. The analysis was designed to identify two groups of candidates. The first group included proteins with estimated effects on CAD that persisted after adjustment for LDL-C or APOB, suggesting CAD-related effects not fully explained by major apoB/LDL-related pathways. The second group included proteins with estimated effects on LDL-C or APOB that persisted after adjustment for genetically predicted HMGCR or PCSK9 protein levels, suggesting lipid-related mechanisms not fully captured by these established lipid-lowering target proxies. These two groups capture complementary therapeutic opportunities, including novel non-lipid CAD targets and lipid-related targets that may still provide benefit beyond established interventions.

We analyzed GWAS summary statistics for 4,907 plasma proteins measured in 35,559 Icelanders from the deCODE plasma proteomics study [43]. GWAS summary statistics were preprocessed and harmonized following the same procedures described above. For each protein, IVs were selected from genome-wide pQTLs using a significance threshold of *P <* 5 × 10^−5^, followed by LD clumping in PLINK (*r*^2^ = 0.01, 1,000 kb window). Therefore, the instrument set could include both cis-pQTLs and trans-pQTLs. Among the 4,907 plasma proteins, 875 with a sufficient number of IVs (*>* 10) were retained for downstream analysis. To identify the first group of candidate protein targets, we first applied MR-APSS, without measured covariate adjustment, to these 875 proteins using CAD as the outcome. Proteins significant after FDR correction (*FDR <* 0.05) were retained as the initial CAD-prioritized protein set and were then evaluated by MRMU with separate adjustment for LDL-C or APOB. This step was used to prioritize proteins whose estimated effects on CAD were not fully explained by these major lipid-related pathways. To identify the second group, we applied MR-APSS to the same 875 proteins using LDL-C and APOB as outcomes, and proteins significant after FDR correction (*FDR <* 0.05) were retained for follow-up. These prescreened proteins were then evaluated by MRMU with separate adjustment for genetically predicted HMGCR or PCSK9 protein levels. HMGCR and PCSK9 were selected because they were the only established lipid-lowering drug target proxies with available protein GWAS data in the current dataset.

As is shown in Fig 7A, six proteins (SLC5A8, CYP3A4, CRIP1, PLA2G7, WNT5A, and HTATIP2) were identified among the 875 proteins to have a significant causal effect on CAD at FDR *<* 0.05 without covariate adjustment. Fig 7B further shows whether these CAD signals persisted after adjustment for APOB and LDL-C using MRMU. Among the six proteins, SLC5A8 exhibited the most stable positive effect under both adjustments of LDL-C and CAD, suggesting that its estimated effect on CAD was not fully explained by these major lipid traits. Prior studies have shown that SLC5A8 is required for butyrate-driven tolerogenic immune programs [44, 45], while butyrate itself has been linked to reduced atherosclerotic inflammation and improved vascular metabolic homeostasis [46]. These findings support SLC5A8 as a biologically plausible CAD protein target for follow-up research. For the other five of the six proteins (CYP3A4, CRIP1, PLA2G7, WNT5A, and HTATIP2), the estimated effects were no longer significant after adjustment for LDL-C or APOB (*P >* 0.05), suggesting that their CAD-related signals may be explained by major apoB/LDL pathways.

**Figure 7:**
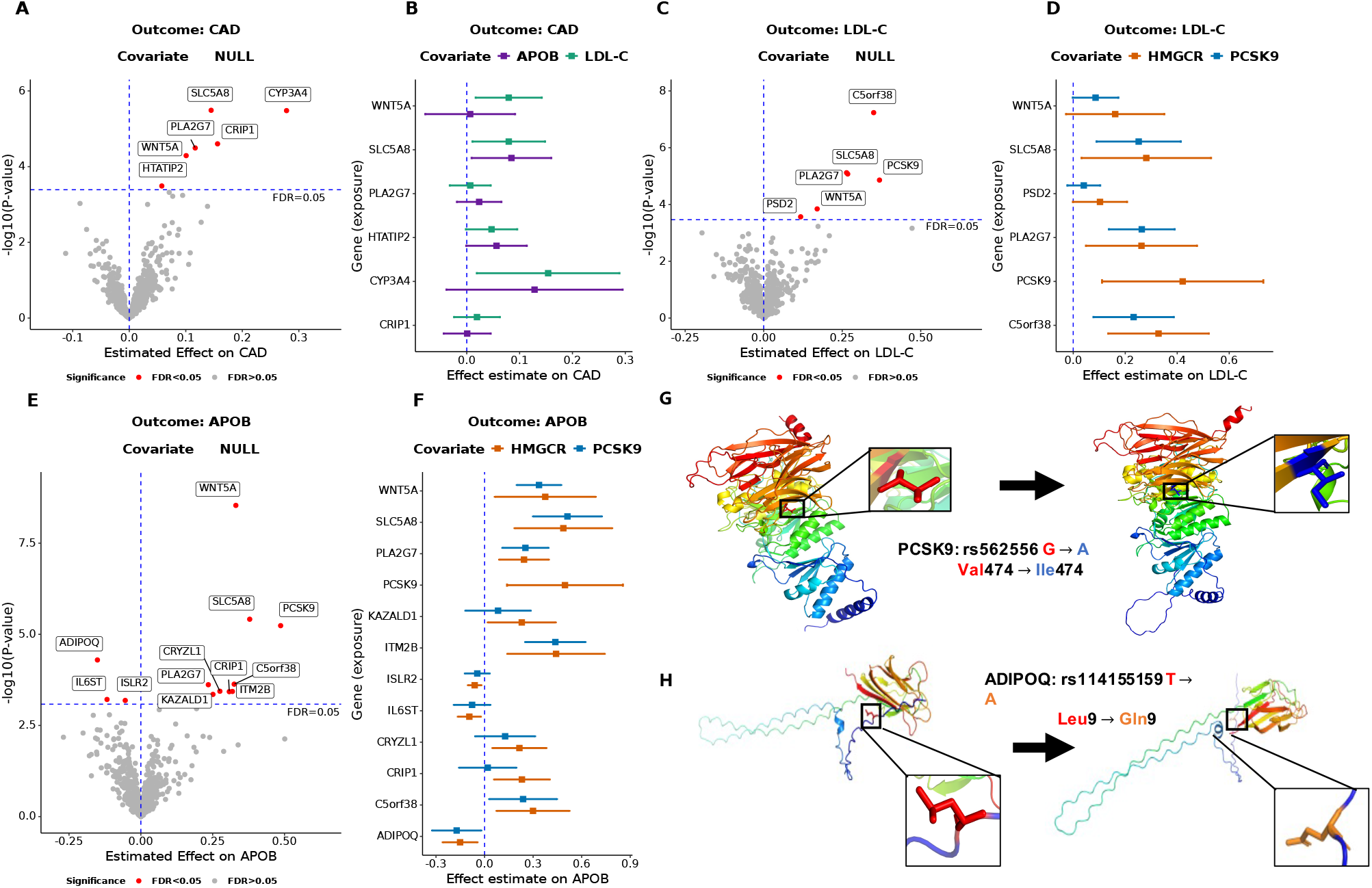
Identification of candidate protein targets for CAD. (**A**) MR-APSS screening analysis of plasma proteins for CAD, with significant proteins defined at FDR *<* 0.05. (**B**) MRMU estimates for proteins highlighted in panel A after separate adjustment for APOB or LDL-C, showing whether the CAD effects persist beyond major lipid-related pathways. (**C**) MR-APSS analysis of plasma proteins on LDL-C identifies proteins with significant effects on LDL-C at FDR *<* 0.05. (**D**) MRMU estimates for proteins highlighted in panel C after separate adjustment for HMGCR or PCSK9, showing whether their effects on LDL-C remain after accounting for established lipid-lowering drug targets. (**E**) MR-APSS analysis of plasma proteins on APOB identifies proteins with significant effects on APOB at FDR *<* 0.05. (**F**) MRMU estimates for proteins highlighted in panel E after separate adjustment for HMGCR or PCSK9, showing whether their effects on APOB persist independently of these established targets. (**G**) AlphaFold3-predicted structural alteration of PCSK9 associated with rs562556 (Val474Ile). (**H**) AlphaFold3-predicted structural alteration of ADIPOQ associated with rs114155159 (Leu9Gln).

We next summarized the second group of candidates, namely proteins with effects on LDL-C or APOB that persisted after adjustment for HMGCR or PCSK9 protein levels. When LDL-C was used as the outcome, MR-APSS identified C5orf38, SLC5A8, PLA2G7, PCSK9, WNT5A, and PSD2 at FDR *<* 0.05 (Fig. 7C). After separate adjustment for HMGCR or PCSK9 using MRMU, SLC5A8, PLA2G7, C5orf38, and WNT5A continued to show positive estimates for LDL-C (Fig. 7D), suggesting that their lipid-related effects were not fully captured by these established lipid-lowering targets. When APOB was used as the outcome, MR-APSS prioritized WNT5A, SLC5A8, PCSK9, C5orf38, CRIP1, ITM2B, PLA2G7, CRYZL1, KAZALD1, ISLR2, IL6ST, and ADIPOQ at FDR *<* 0.05 (Fig. 7E). After adjustment for HMGCR or PCSK9, several proteins, including WNT5A, SLC5A8, PLA2G7, ITM2B, and C5orf38, retained positive estimates, whereas ADIPOQ showed a negative estimate (Fig. 7F). These results highlight candidate lipid-related proteins whose effects on LDL-C or APOB may not be fully explained by the HMGCR or PCSK9 pathways.

PCSK9 served as a positive control for the lipid-related target analysis. PCSK9 remained significantly associated with LDL-C and APOB after adjustment for HMGCR, consistent with extensive genetic and clinical evidence showing that PCSK9 independently regulates LDL metabolism by reducing LDL receptor recycling [47, 48] and PCSK9 inhibition lowers LDL-C substantially even after statin therapy [49]. In Fig. 7G, we present the AlphaFold3-predicted structural alteration of PCSK9 associated with the variant *rs*562556, a PCSK9 pQTL in the deCODE plasma proteomics study (pQTL *P* = 1.093 × 10^−16^). corresponding to the missense mutation *NM*_1_74936.4 : *c*.1420*G > A*. This variant corresponds to the missense mutation *NM*_1_74936.4 : *c*.1420*G > A*, leading to the amino acid substitution Val474Ile. AlphaFold3-based prediction suggested a local structural difference around residue 474. Because PCSK9 regulates LDL metabolism by promoting LDLR degradation and limiting its recycling to the hepatocyte surface [48], this local structural change may have functional consequences for PCSK9-mediated LDLR regulation. Consistent with this possibility, previous population-based studies have linked rs562556 to lipid-related phenotypes. A meta-analysis showed that G-allele carriers had lower total cholesterol and LDL-C than non-carriers [50], and a longitudinal cohort study found that the A allele was associated with higher circulating PCSK9 concentrations and increased carotid plaque risk [51]. These findings suggest that rs562556 may be associated with local structural differences in PCSK9 and provide additional biological context for its link to lipid-related phenotypes.

ADIPOQ provides another example in which a missense variant can be used to explore potential local protein structural changes. In the APOB analysis, ADIPOQ showed a negative effect on APOB after adjustment for HMGCR or PCSK9, suggesting that its effect on APOB was not fully explained by these established lipid-lowering targets. This direction is consistent with previous evidence from type 2 diabetes populations linking adiponectin to a more favorable lipid profile, including inverse relationships with apoB100 levels [52, 53]. We further examined rs114155159, an ADIPOQ cis-pQTL identified in the deCODE plasma proteomics study (pQTL *P* = 6.984 × 10^−48^). This variant is also annotated as a missense variant, leading to the amino acid substitution Leu9Gln. AlphaFold3-based prediction further suggested a local structural difference around the variant site(Fig. 7H), highlighting ADIPOQ as a candidate for follow-up research.

## Discussion

The central contribution of MRMU is to jointly account for measured covariates that capture pleiotropic pathways and unmeasured confounders in GWAS summary statistics, thereby improving the reliability of MR analyses. By using exposure-selected IVs, MRMU allows covariate-adjusted and unadjusted estimates to be compared within the same IV framework. This helps interpret pathway-related effects. In simulations and negative control analyses, MRMU showed better performance than existing MR methods. In the CAD analysis, MRMU prioritized a set of cardiometabolic risk factors and showed that many estimated effects on CAD changed after adjustment for lipid and metabolic syndrome-related covariates. MRMU also supported blood pressure-related traits mediating the urate–CAD relationship and nominated candidate drug targets beyond established lipid-lowering pathways. These findings highlight the utility of MRMU for robust causal inference, pathway interpretation, and therapeutic prioritization.

MRMU identified SBP, BMI, smoking initiation, APOB, and LDL-C as CAD risk factors with relatively stable estimates across different covariate-adjustment settings. In the pathway-related covariate analyses, these traits did not meet our definition of a difference after adjustment for lipid traits, metabolic syndrome-related traits, or lifestyle factors. This pattern further supports their role as CAD risk factors, suggesting that their estimated effects were not primarily driven by the measured covariate groups considered in this analysis, but instead reflected direct effects on CAD within the MRMU framework.

In the urate–CAD analysis, sex hormone-binding globulin (SHBG) showed a covariate-dependent pattern. Adjustment for SHBG changed the estimated urate effect on CAD, and urate-associated IVs showed a high variance ratio and a negative IV-effect correlation with SHBG (Fig. 6). Because SHBG is more closely related to endocrine and metabolic regulation than to blood pressure directly, this signal may reflect a broader endocrine-metabolic component of the urate–CAD relationship. This complements the stronger blood pressure-related pattern observed for systolic and diastolic blood pressure, suggesting that urate-associated genetic instruments may capture both blood pressure-related and broader metabolic signals.

The candidate protein target analysis provided additional support for the importance of LDL-C and APOB in CAD risk. Among proteins prioritized for CAD, several MR-APSS signals became non-significant after adjustment for LDL-C or APOB using MRMU. This pattern suggests that these protein effects on CAD may be partly explained by major apoB/LDL-related pathways. Together with the CAD risk factor analysis, where LDL-C and APOB showed robust effects across datasets, this result provides further evidence that apoB-containing lipoprotein pathways play a central role in CAD risk.

The choice of measured covariates is an important practical consideration in MRMU. Because MRMU estimates the direct effect of an exposure conditional on the included covariates, using different subsets of measured covariates may lead to different effect estimates and biological interpretations. Omitting relevant covariates may leave residual pathway-related confounding and increase the risk of false positive findings, whereas including too many irrelevant or null covariates may reduce statistical power and estimation efficiency, as also observed in our simulations with multiple measured covariates. In practice, we recommend starting from a broad set of biologically plausible covariates informed by prior literature and domain knowledge, followed by data-driven prescreening to reduce the inclusion of irrelevant traits. In our CAD analysis, we first used MRAPSS as a marginal screening step to identify traits with evidence of association with the outcome, and then further excluded candidate covariates that showed weak pleiotropic relevance to the exposure-selected IVs. Specifically, for each exposure, we selected IVs from the exposure GWAS and retained only candidate covariates for which at least five exposure-selected IVs reached genome-wide significance in the corresponding covariate GWAS. This strategy aims to balance confounding control and statistical efficiency by prioritizing covariates that are both biologically relevant and genetically connected to the exposure instruments. In addition, we recommend sensitivity analyses using biologically defined covariate subsets, such as lipid traits, metabolic syndrome-related traits, or lifestyle factors, as well as one covariate at a time analyses when appropriate. Such analyses can help determine whether an exposure–outcome association is robust across covariate choices or primarily depends on specific pathway-related covariates, as illustrated in our pathway-related covariate dependence analysis of exposure–CAD relationships.

Several points should be considered when interpreting our results. First, covariate adjustment may introduce collider bias when inappropriate variables are conditioned on, especially when a covariate is influenced by both the exposure and outcome. MRMU does not explicitly identify collider structures, but we reduced this risk by prescreening measured covariates before model fitting. In our analysis, MR-APSS was used to retain covariates with evidence of causal relevance to the outcome, rather than adjusting for traits only because they were related to the exposure. Although collider bias cannot be fully eliminated, this step helps avoid inappropriate adjustment and supports more interpretable pathway-adjusted inference. Second, the sensitivity analysis comparing MRMU variants should be viewed as a diagnostic evaluation of measured-covariate adjustment and unmeasured-confounding adjustment, not as a strict decomposition of effect changes. These two components may interact, especially for correlated cardiometabolic traits. Therefore, changes observed in the MRMU variants cannot be uniquely attributed to either measured pathways or unmeasured confounding alone. Still, the comparison helps show how incomplete adjustment for either component may lead to broader sets of significant findings and supports the need to account for both measured covariates and unmeasured confounders in MR analyses using GWAS summary statistics.

## Methods

### The MRMU model

Let *X* be the exposure variable, *Y* be the outcome variable, and 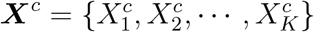 be the *K* measured covariates that may be associated with both the exposure *X* and the outcome *Y*. Let 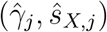 and 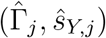 be the effect estimates and standard errors at the *j*-th SNP for *X* and *Y*, respectively. For the measured covariates, let 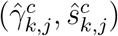 denote the effect estimate and standard error of the *j*-th SNP on the *k*-th measured covariate 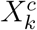, for *k* = 1, · · ·, *K*, and define 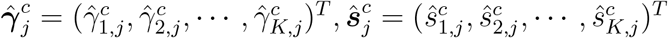. To obtain a reliable estimate of the causal effect that is robust to both measured covariates and unmeasured confounders, MRMU integrates three components to model the relationships among the estimated effect sizes 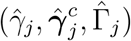, namely the inferential model, the polygenic effect model, and the estimation error model:

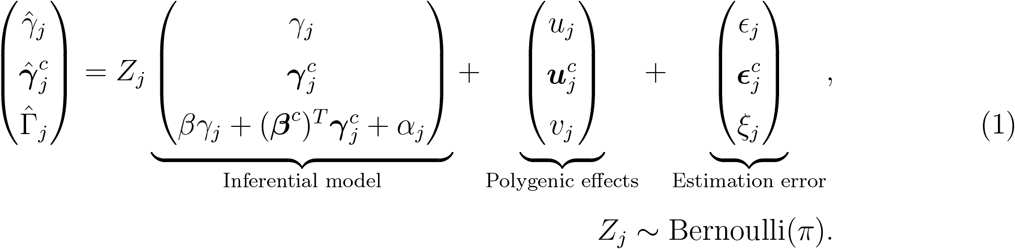

Here *u*_*j*_, 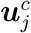, and *v*_*j*_ are the polygenic effects of SNP *G*_*j*_ on the exposure *X*, the measured covariates ***X***^*c*^, and the outcome *Y*, respectively; *ϵ*_*j*_, 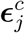, and *ξ*_*j*_ are the corresponding estimation errors. The parameter *γ*_*j*_ denotes the extra effect of *G*_*j*_ on the exposure *X*, measuring IV strength, and 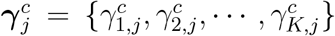 denotes the IV effects of *G*_*j*_ on the *K* measured covariates. The parameter *α*_*j*_ is the direct effect of *G*_*j*_ on the outcome *Y, β* is the causal effect of *X* on *Y*, and 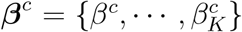 denotes the effects of the measured covariates ***X***^*c*^ on *Y*. The latent indicator *Z*_*j*_ takes value 1 if *G*_*j*_ has nonzero IV strength (*γ*_*j*_≠ 0) and is therefore used for causal inference under the inferential model, and 0 otherwise.

To account for possible unmeasured confounders including polygenicity, unmeasured pleiotropy, and sample structure, we model the polygenic effects and estimation errors as follows:

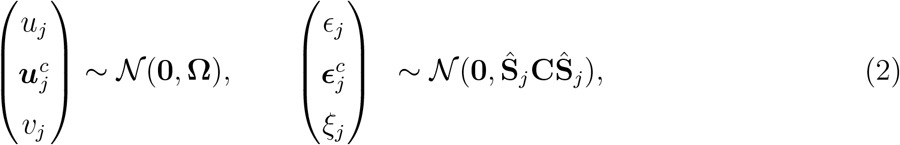

where 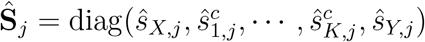, *N* (**0, Ω**) is a multivariate normal density with mean **0** and covariance **Ω**, and *N* (**0, Ŝ**_*j*_ **CŜ**_*j*_) is a multivariate normal density with mean **0** and covariance **S**_*j*_. We allow the off-diagonal elements of **Ω** to be nonzero to account for correlations among polygenic effects induced by shared genetic architecture and unmeasured pleiotropy. We further model the estimation error covariance through **C**, allowing its diagonal elements to exceed one to capture residual inflation in GWAS summary statistics, and its off-diagonal elements to be nonzero to account for correlations induced by sample structure, including population stratification, assortative mating, sample overlap, and cryptic relatedness.

After accounting for unmeasured confounders through the polygenic effect model and the estimation error model, MRMU performs causal inference with an inferential component analogous to multivariable regression, while using IVs associated with the exposure variable *X*. Specifically, for *G*_*j*_ as an IV for *X* (*Z*_*j*_ = 1), the inferential model relies on the following assumptions: (1) *G*_*j*_ is associated with the exposure, that is, *γ*_*j*_≠ 0; (2) *G*_*j*_ is independent of any confounders beyond the measured covariates ***X***^*c*^; (3) the IV effect on the exposure and the IV effects on the measured covariates 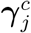 may be correlated, reflecting the close relationship between the exposure and the measured covariates; and (4) the direct effect of *G*_*j*_ on the outcome, *α*_*j*_, representing uncorrelated pleiotropy, is independent of 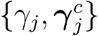. With these assumptions, we model

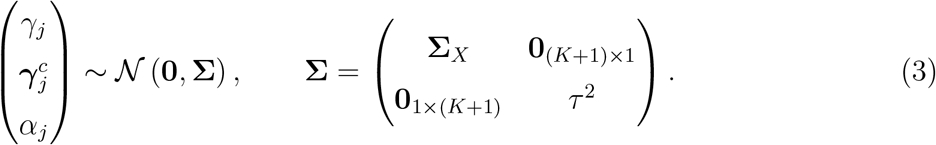

Here, **Σ**_*X*_ is the variance–covariance matrix of the IV effects on the exposure *X* and the *K* measured covariates ***X***^*c*^. Its diagonal elements correspond to 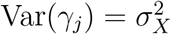 for the exposure and 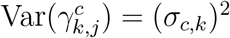 for the *k*-th measured covariate 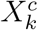, *k* = 1, · · ·, *K*. We set the covariance between *α*_*j*_ and 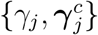 to zero because *α*_*j*_ is assumed to be independent of these IV effects.

To quantify IV-level pleiotropic overlap between the exposure and measured covariates, we define two quantities: the variance ratio and the IV-effect correlation. Here, IV-level pleiotropic overlap refers to the extent to which IVs selected for the exposure also act on measured covariates. For the *k*-th measured covariate, the variance ratio is defined as

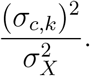

This quantity measures the relative magnitude of IV effects on *X*^*c*^ compared with their effects on *X*. A larger value indicates that exposure-selected IVs have stronger effects on the measured covariate relative to the exposure. We further define the IV-effect correlation between *X* and 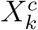 as

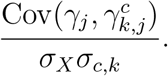

The IV-effect correlation quantifies the extent of IV-level pleiotropic overlap between exposure-selected IV effects on *X* and on 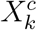. These correlations should be interpreted among exposure-selected IVs, rather than as genome-wide genetic correlations captured by the polygenic component. The variance ratio and IV-effect correlation provide quantities of how strongly exposure-selected IVs capture measured covariate signals and help assess the relevance of measured covariates in the MRMU model.

So far, we have assumed that SNPs are independent. In practice, however, SNPs are correlated because of linkage disequilibrium (LD), so GWAS summary statistics represent marginal rather than conditional SNP effects. Under LD, both the inferential effects and the polygenic background effects are transformed into LD-tagged weighted sums over correlated SNPs. Following the same principle as in MR-APSS, but extending the trait system from (*X, Y*) to (*X*, ***X***^*c*^, *Y*), the MRMU summary-level model can be rewritten in an LD-adjusted form. The full derivation of the LD-adjusted MRMU model is provided in Section 1.1 of the Supplementary Note. After integrating out the LD-tagged inferential and polygenic effects together with the latent indicator *Z*_*j*_, the observed summary statistics follow the marginal model

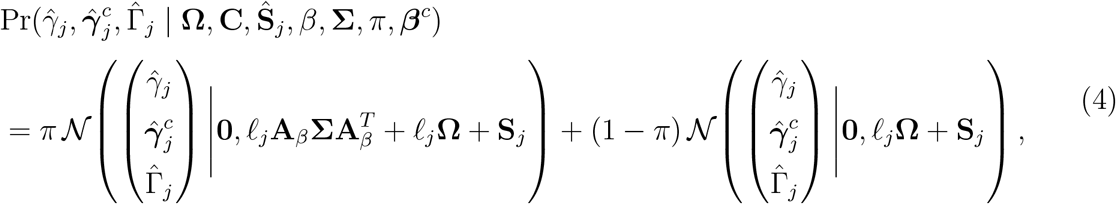

where 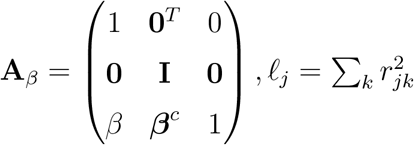 is the LD score of SNP *j*, **0** is a zero vector of length *K*, and **I** is the *K* × *K* identity matrix. Equation(4) clarifies why **Ω** and **C** can be pre-estimated using LD score regression (LDSC). Under the polygenic model, the second moment of GWAS summary statistics can be decomposed into an LD-dependent component and an LD-independent component. The LD-dependent component scales linearly with LD score and reflects background polygenic covariance across traits, corresponding to **Ω** in MRMU. In contrast, the LD-independent component is captured by the LDSC intercept and reflects residual inflation and cross-trait error correlation induced by sample structure, including population stratification, assortative mating, sample overlap, and cryptic relatedness; this corresponds to **C** in MRMU. Because MRMU assumes that the polygenic component explains the majority of genome-wide variation, this separation allows **Ω** and **C** to be estimated from genome-wide summary statistics using LDSC and then treated as fixed background quantities in the MRMU likelihood.

### IV selection

In MRMU, IVs are selected based on the exposure *X* of interest. In practice, MRMU uses a moderate *p*-value threshold (typically 5 × 10^−5^), or equivalently the selection criterion 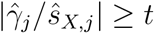 with *t* = 4.06, to screen candidate IVs from the GWAS summary statistics of *X*. The PLINK clumping procedure (–clump-kb 1000 –clump-r2 0.001) is then applied to retain approximately independent variants.

We denote the observed GWAS summary statistics after IV selection by

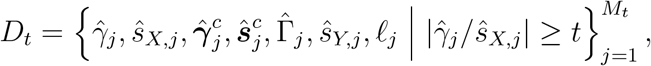

where *M*_*t*_ is the number of SNPs retained after IV selection and *ℓ*_*j*_ is the LD score of the *j*-th selected SNP.

We define

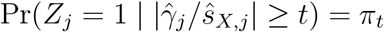

as the conditional probability that the *j*-th selected SNP belongs to the inferential component, that is, has a nonzero effect on the exposure after IV selection.

### Parameter estimation

Estimation based on GWAS summary statistics after IV selection can be challenging in MRMU. Because IVs are selected based on the exposure of interest *X*, their effects on the measured covariates ***X***^*c*^ are not necessarily strong, so some elements of 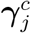 may be close to zero. In addition, not all measured covariates involved are expected to play substantial roles in a given exposure-outcome relationship, implying that many elements of ***β***^*c*^ may also be small or close to zero. These features can lead to weak identification and unstable estimation if ***β***^*c*^ is treated as a fixed-effect parameter. To address this issue, we model ***β***^*c*^ as a random effect and assign the prior

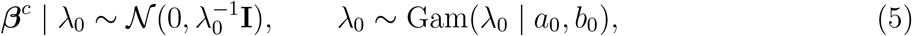

where we set *a*_0_ = 1 and *b*_0_ = 1.

Another issue in the selected IV dataset is the “winner’s curse”, whereby IV selection based on statistical significance distorts the distribution of GWAS summary statistics for the selected SNPs and may bias causal effect estimation. To correct for winner’s curse, rather than maximizing the ordinary likelihood [54], we maximize a likelihood conditional on the selection rule. Under models (4) and (5), the conditional likelihood for MRMU is

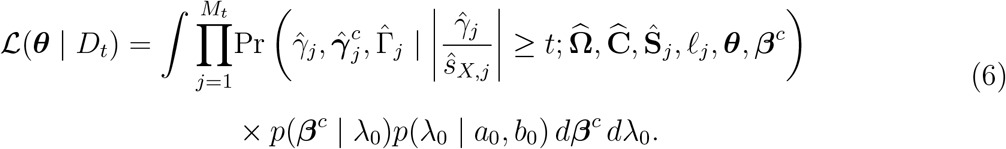

where ***θ*** = {*β*, **Σ**_*X*_, *τ* ^2^, *π*_*t*_} is the set of unknown parameters to be estimated, 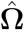 and 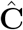 are pre-estimates of **Ω** and **C**, and *t* denotes the IV selection threshold. The detailed derivation of this conditional likelihood is provided in Supplementary Note Section 1.2. Direct maximization of Eq. (6) is difficult because the likelihood involves integration over the latent variables and the random effects ***β***^*c*^ and *λ*_0_. We therefore develop a variational EM algorithm to estimate ***θ***. For simplicity, we omit the known quantities 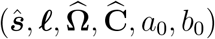 in the derivation below.

Let 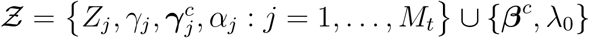 denote the collection of latent variables, Let *q*(Ƶ | *D*_*t*_) be a variational distribution for these latent variables conditional on the selected IV dataset *D*_*t*_. Then the conditional log-likelihood in Eq. (6) can be decomposed as

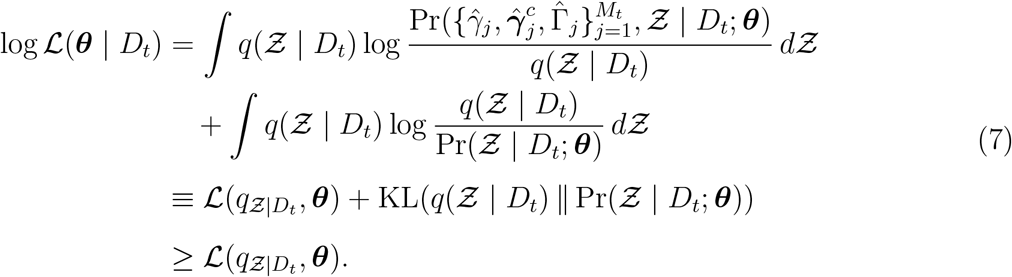

The inequality follows from the non-negativity of the Kullback–Leibler divergence. The quantity 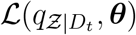 is the evidence lower bound (ELBO) of the conditional log-likelihood.

We assume that the variational distribution factorizes as

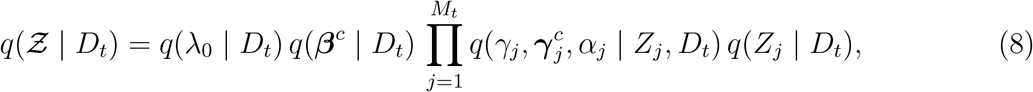

With this factorization, the ELBO becomes tractable. In the E-step, we optimize the ELBO with respect to the variational distribution while holding ***θ*** fixed. In the M-step, we update ***θ*** by maximizing the ELBO with the variational distribution fixed. These two steps are repeated until convergence. Details of the ELBO derivation and the full variational EM updates are provided in Supplementary Note Section 1.3.

### Inference

To test for the existence of a causal effect *β* between *X* and *Y*, we perform a likelihood ratio test comparing the model under the null hypothesis ℋ_0_ : *β* = 0 with the model under the alternative hypothesis ℋ_1_ : *β /*= 0. Let 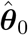 denote the parameter estimates under ℋ_0_ and 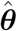 denote the parameter estimates under ℋ_1_. The test statistic is

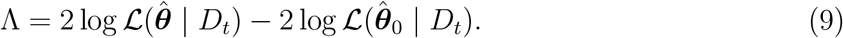

We approximate the likelihood terms above by the converged ELBO values under H_0_ and H_1_, respectively.

## Data Availability

All GWAS summary statistics analyzed in this study are publicly available. Detailed information on the GWAS sources is provided in Supplementary Tables S2 and S3. The MRMU software is available at https://github.com/hxh0928/MRMU. The datasets and source code for reproducing the analyses are available at https://github.com/hxh0928/MRMU-reproduce.

https://github.com/hxh0928/MRMU-reproduce

## Acknowledgements

This work was supported in part by the National Natural Science Foundation of China (NSFC) (Grant No. 12501401); the Innovation and Technology Commission (ITCPD/17-9, PRP/066/23FX); Research Grants Council, University Grants Committee (16307322, 16302823, 16309424, and 16308925); The Hong Kong University of Science and Technology Startup Grants (R9405 and Z0428 from the Big Data Institute); Guangdong Major Project of Basic Research (2026B0303000015); Shenzhen Newly Introduced High-end Talents Scientific Research Start-up Project (Pengcheng Peacock Plan, Grant No. 892007202);

## Notes

### Competing Interest Statement

The authors have declared no competing interest.

### Clinical Protocols

https://github.com/hxh0928/MRMU

https://github.com/hxh0928/MRMU-reproduce

