## supplemental notes tables, and figures for "MRMU: A New Paradigm for Mendelian Randomization by Accounting for Measured Covariates and Unmeasured Confounders"

### Supplementary Information:

**Figure** S7: Comparison of MRMU model variants in the CAD risk factor analysis 25

### Supplementary Tables S1-S2

**Table** S1: Instrument variable selection and LD clumping settings across MR methods. 26

**Table** S2: GWAS sources for traits in the negative control analysis. . . . . . . . . 27

**Table** S3: GWAS sources for candidate traits used in the CAD real-data analysis. 28

### 1 Supplementary notes

#### 1.1 Derivation of the LD-adjusted MRMU model

For independent SNPs, the MRMU summary-level model can be written as

$$\begin{pmatrix}\hat{\gamma}_j\\ \hat{\boldsymbol{\gamma}}_j^c\\ \hat{\Gamma}_j\end{pmatrix} = Z_j \underbrace{\begin{pmatrix}\gamma_j\\ \boldsymbol{\gamma}_j^c\\ \beta\gamma_j + (\boldsymbol{\beta}^c)^T\boldsymbol{\gamma}_j^c + \alpha_j\end{pmatrix}}_{\text{inferential component}} + \underbrace{\begin{pmatrix}u_j\\ \boldsymbol{u}_j^c\\ v_j\end{pmatrix}}_{\text{polygenic component}} + \underbrace{\begin{pmatrix}\epsilon_j\\ \boldsymbol{\epsilon}_j^c\\ \xi_j\end{pmatrix}}_{\text{estimation error}}, \qquad Z_j \sim \text{Bernoulli}(\pi), \tag{1}$$

For notational convenience, define

$$\hat{\mathbf{b}}_j = \begin{pmatrix}\hat{\gamma}_j\\ \hat{\boldsymbol{\gamma}}_j^c\\ \hat{\Gamma}_j\end{pmatrix}, \qquad \boldsymbol{\eta}_j = \begin{pmatrix}\gamma_j\\ \boldsymbol{\gamma}_j^c\\ \alpha_j\end{pmatrix}, \qquad \mathbf{A}_\beta = \begin{pmatrix}1 & \mathbf{0}^T & 0\\ \mathbf{0} & \mathbf{I} & \mathbf{0}\\ \beta & (\boldsymbol{\beta}^c)^T & 1\end{pmatrix},$$

so that the inferential component in Eq. (1) can be written compactly as

$$Z_j\mathbf{A}_\beta\boldsymbol{\eta}_j.$$

Under linkage disequilibrium (LD), GWAS summary statistics estimate marginal SNP effects, so the observed effect at SNP $j$ is a weighted sum of effects contributed by neighboring SNPs. Let $r_{jk}$ denote the LD correlation between the $j$-th and $k$-th SNPs. Then the marginal summary statistics satisfy

$$\hat{\mathbf{b}}_j = \sum_k r_{jk}\Big(Z_k\mathbf{A}_\beta\boldsymbol{\eta}_k + \boldsymbol{\omega}_k\Big) + \mathbf{e}_j, \tag{2}$$

where

$$\boldsymbol{\omega}_k = \begin{pmatrix}u_k\\ \boldsymbol{u}_k^c\\ v_k\end{pmatrix}, \qquad \mathbf{e}_j = \begin{pmatrix}\epsilon_j\\ \boldsymbol{\epsilon}_j^c\\ \xi_j\end{pmatrix}.$$

Following the same LD-adjustment strategy as in MRAPSS [1], we define the LD-tagged latent effects

$$\gamma_j^* = \sum_k r_{jk}\gamma_k, \qquad \boldsymbol{\gamma}_j^{c*} = \sum_k r_{jk}\boldsymbol{\gamma}_k^c, \qquad \alpha_j^* = \sum_k r_{jk}\alpha_k,$$

and

$$u_j^* = \sum_k r_{jk}u_k, \qquad \boldsymbol{u}_j^{c*} = \sum_k r_{jk}\boldsymbol{u}_k^c, \qquad v_j^* = \sum_k r_{jk}v_k.$$

Writing

$$\boldsymbol{\eta}_j^* = \begin{pmatrix}\gamma_j^*\\ \boldsymbol{\gamma}_j^{c*}\\ \alpha_j^*\end{pmatrix}, \qquad \boldsymbol{\omega}_j^* = \begin{pmatrix}u_j^*\\ \boldsymbol{u}_j^{c*}\\ v_j^*\end{pmatrix},$$

we obtain the LD-adjusted form

$$\hat{\mathbf{b}}_j = Z_j\mathbf{A}_\beta\boldsymbol{\eta}_j^* + \boldsymbol{\omega}_j^* + \mathbf{e}_j. \tag{3}$$

Under the MRMU assumptions,

$$\boldsymbol{\eta}_j \sim \mathcal{N}(\mathbf{0}, \boldsymbol{\Sigma}), \qquad \boldsymbol{\omega}_j \sim \mathcal{N}(\mathbf{0}, \boldsymbol{\Omega}), \qquad \mathbf{e}_j \sim \mathcal{N}(\mathbf{0}, \mathbf{S}_j),$$

with independence across SNPs. Therefore, using

$$\ell_j = \sum_k r_{jk}^2,$$

we have

$$\mathrm{Cov}(\boldsymbol{\eta}_j^*) = \sum_k r_{jk}^2 \boldsymbol{\Sigma} = \ell_j \boldsymbol{\Sigma},$$

and hence

$$\mathrm{Cov}(\mathbf{A}_\beta \boldsymbol{\eta}_j^*) = \ell_j \mathbf{A}_\beta \boldsymbol{\Sigma} \mathbf{A}_\beta^T.$$

Similarly,

$$\mathrm{Cov}(\boldsymbol{\omega}_j^*) = \sum_k r_{jk}^2 \boldsymbol{\Omega} = \ell_j \boldsymbol{\Omega}.$$

It follows that the LD-adjusted conditional distribution of the observed summary statistics is

$$\hat{\mathbf{b}}_j \mid Z_j = 1 \sim \mathcal{N}\big(\mathbf{0}, \ell_j \mathbf{A}_\beta \boldsymbol{\Sigma} \mathbf{A}_\beta^T + \ell_j \boldsymbol{\Omega} + \mathbf{S}_j\big)\,, \tag{4}$$

and

$$\hat{\mathbf{b}}_j \mid Z_j = 0 \sim \mathcal{N}(\mathbf{0}, \ell_j \boldsymbol{\Omega} + \mathbf{S}_j)\,. \tag{5}$$

Marginalizing over $Z_j \sim \mathrm{Bernoulli}(\pi)$ yields the observed-data mixture model

$$\begin{aligned} &\Pr(\hat{\gamma}_j, \hat{\boldsymbol{\gamma}}_j^c, \hat{\Gamma}_j \mid \boldsymbol{\Omega}, \mathbf{C}, \hat{\mathbf{S}}_j, \beta, \boldsymbol{\beta}^c) \\ &= \pi \mathcal{N}\left( \begin{pmatrix} \hat{\gamma}_j \\ \hat{\boldsymbol{\gamma}}_j^c \\ \hat{\Gamma}_j \end{pmatrix} \middle| \mathbf{0}, \ell_j \mathbf{A}_\beta \boldsymbol{\Sigma} \mathbf{A}_\beta^T + \ell_j \boldsymbol{\Omega} + \mathbf{S}_j \right) \\ &\quad + (1-\pi) \mathcal{N}\left( \begin{pmatrix} \hat{\gamma}_j \\ \hat{\boldsymbol{\gamma}}_j^c \\ \hat{\Gamma}_j \end{pmatrix} \middle| \mathbf{0}, \ell_j \boldsymbol{\Omega} + \mathbf{S}_j \right). \end{aligned} \tag{6}$$

This derivation shows that, under LD, the inferential covariance is propagated jointly through the exposure, the measured covariates, and the outcome via $\ell_j \mathbf{A}_\beta \boldsymbol{\Sigma} \mathbf{A}_\beta^T$, while the background polygenic covariance is captured by $\ell_j \boldsymbol{\Omega}$.

#### 1.2 Derivation of the conditional likelihood after IV selection

We derive the likelihood conditional on the IV selection rule. The selected IV dataset $D_t$ contains only SNPs satisfying

$$\left| \frac{\hat{\gamma}_j}{\hat{s}_{X,j}} \right| \geq t.$$

Therefore, the likelihood should be constructed from the conditional distribution of

$$\begin{pmatrix} \hat{\gamma}_j \\ \hat{\boldsymbol{\gamma}}_j^c \\ \hat{\Gamma}_j \end{pmatrix}$$

given the IV selection rule.

From the marginal mixture model in Eq. (6), before conditioning on IV selection, the density for the case $Z_j = 1$ is

$$\mathcal{N}\left(\begin{pmatrix}\hat{\gamma}_j\\ \hat{\boldsymbol{\gamma}}_j^c\\ \hat{\Gamma}_j\end{pmatrix}\middle|\mathbf{0}, \ell_j\mathbf{A}_\beta\boldsymbol{\Sigma}\mathbf{A}_\beta^T + \ell_j\widehat{\boldsymbol{\Omega}} + \hat{\mathbf{S}}_j\widehat{\mathbf{C}}\hat{\mathbf{S}}_j\right),$$

whereas the density for the case $Z_j = 0$ is

$$\mathcal{N}\left(\begin{pmatrix}\hat{\gamma}_j\\ \hat{\boldsymbol{\gamma}}_j^c\\ \hat{\Gamma}_j\end{pmatrix}\middle|\mathbf{0}, \ell_j\widehat{\boldsymbol{\Omega}} + \hat{\mathbf{S}}_j\widehat{\mathbf{C}}\hat{\mathbf{S}}_j\right).$$

For a selected SNP, the conditional density is obtained by dividing each density by the probability that the SNP passes the IV selection rule. Specifically, for $Z_j = 1$,

$$\begin{aligned}&\Pr\left(\hat{\gamma}_j, \hat{\boldsymbol{\gamma}}_j^c, \hat{\Gamma}_j \mid \left|\frac{\hat{\gamma}_j}{\hat{s}_{X,j}}\right| \geq t, Z_j = 1; \widehat{\boldsymbol{\Omega}}, \widehat{\mathbf{C}}, \hat{\mathbf{S}}_j, \ell_j, \boldsymbol{\theta}, \boldsymbol{\beta}^c\right)\\ =&\frac{\mathcal{N}\left(\begin{pmatrix}\hat{\gamma}_j\\ \hat{\boldsymbol{\gamma}}_j^c\\ \hat{\Gamma}_j\end{pmatrix}\middle|\mathbf{0}, \ell_j\mathbf{A}_\beta\boldsymbol{\Sigma}\mathbf{A}_\beta^T + \ell_j\widehat{\boldsymbol{\Omega}} + \hat{\mathbf{S}}_j\widehat{\mathbf{C}}\hat{\mathbf{S}}_j\right)}{\Pr\left(\left|\frac{\hat{\gamma}_j}{\hat{s}_{X,j}}\right| \geq t \mid Z_j = 1; \widehat{\boldsymbol{\Omega}}, \widehat{\mathbf{C}}, \hat{\mathbf{S}}_j, \ell_j, \boldsymbol{\theta}, \boldsymbol{\beta}^c\right)}.\end{aligned}$$

Similarly, for $Z_j = 0$,

$$\begin{aligned}&\Pr\left(\hat{\gamma}_j, \hat{\boldsymbol{\gamma}}_j^c, \hat{\Gamma}_j \mid \left|\frac{\hat{\gamma}_j}{\hat{s}_{X,j}}\right| \geq t, Z_j = 0; \widehat{\boldsymbol{\Omega}}, \widehat{\mathbf{C}}, \hat{\mathbf{S}}_j, \ell_j\right)\\ =&\frac{\mathcal{N}\left(\begin{pmatrix}\hat{\gamma}_j\\ \hat{\boldsymbol{\gamma}}_j^c\\ \hat{\Gamma}_j\end{pmatrix}\middle|\mathbf{0}, \ell_j\widehat{\boldsymbol{\Omega}} + \hat{\mathbf{S}}_j\widehat{\mathbf{C}}\hat{\mathbf{S}}_j\right)}{\Pr\left(\left|\frac{\hat{\gamma}_j}{\hat{s}_{X,j}}\right| \geq t \mid Z_j = 0; \widehat{\boldsymbol{\Omega}}, \widehat{\mathbf{C}}, \hat{\mathbf{S}}_j, \ell_j\right)}.\end{aligned}$$

Because the selection rule is two-sided, the selection probability for $Z_j = 1$ is

$$\begin{aligned}&\Pr\left(\left|\frac{\hat{\gamma}_j}{\hat{s}_{X,j}}\right| \geq t \mid Z_j = 1; \widehat{\boldsymbol{\Omega}}, \widehat{\mathbf{C}}, \hat{\mathbf{S}}_j, \ell_j, \boldsymbol{\theta}, \boldsymbol{\beta}^c\right)\\ =&2\Phi\left(\frac{-t\hat{s}_{X,j}}{\sqrt{\ell_j\sigma_X^2 + \ell_j\widehat{\Omega}_{11} + \widehat{C}_{11}\hat{s}_{X,j}^2}}\right) = 2\widehat{\mathcal{T}}_{1,j}.\end{aligned}$$

Similarly, the selection probability for $Z_j = 0$ is

$$\begin{aligned}&\Pr\left(\left|\frac{\hat{\gamma}_j}{\hat{s}_{X,j}}\right| \geq t \mid Z_j = 0; \widehat{\boldsymbol{\Omega}}, \widehat{\mathbf{C}}, \hat{\mathbf{S}}_j, \ell_j\right)\\ =&2\Phi\left(\frac{-t\hat{s}_{X,j}}{\sqrt{\ell_j\widehat{\Omega}_{11} + \widehat{C}_{11}\hat{s}_{X,j}^2}}\right) = 2\widehat{\mathcal{T}}_{0,j}.\end{aligned}$$

After IV selection, $\pi_t$ denotes the conditional probability that a selected SNP has $Z_j = 1$:

$$\pi_t = \Pr\left(Z_j = 1 \mid \left|\frac{\hat{\gamma}_j}{\hat{s}_{X,j}}\right| \geq t\right).$$

Therefore, for the $j$-th selected SNP, the conditional mixture density is

$$\begin{aligned}
&\Pr\left(\hat{\gamma}_j, \hat{\boldsymbol{\gamma}}_j^c, \hat{\Gamma}_j \mid \left|\frac{\hat{\gamma}_j}{\hat{s}_{X,j}}\right| \geq t; \widehat{\boldsymbol{\Omega}}, \widehat{\mathbf{C}}, \hat{\mathbf{S}}_j, \ell_j, \boldsymbol{\theta}, \boldsymbol{\beta}^c\right) \\
&= \pi_t \frac{\mathcal{N}\left(\begin{pmatrix}\hat{\gamma}_j \\ \hat{\boldsymbol{\gamma}}_j^c \\ \hat{\Gamma}_j\end{pmatrix} \middle| \mathbf{0}, \ell_j \mathbf{A}_\beta \boldsymbol{\Sigma} \mathbf{A}_\beta^T + \ell_j \widehat{\boldsymbol{\Omega}} + \hat{\mathbf{S}}_j \widehat{\mathbf{C}} \hat{\mathbf{S}}_j\right)}{2\widehat{\mathcal{T}}_{1,j}} \\
&\quad + (1 - \pi_t) \frac{\mathcal{N}\left(\begin{pmatrix}\hat{\gamma}_j \\ \hat{\boldsymbol{\gamma}}_j^c \\ \hat{\Gamma}_j\end{pmatrix} \middle| \mathbf{0}, \ell_j \widehat{\boldsymbol{\Omega}} + \hat{\mathbf{S}}_j \widehat{\mathbf{C}} \hat{\mathbf{S}}_j\right)}{2\widehat{\mathcal{T}}_{0,j}}.
\end{aligned}$$

Assuming that the selected SNPs are approximately independent after clumping, the conditional likelihood over all $M_t$ selected SNPs is the product of the above conditional densities. Since $\boldsymbol{\beta}^c$ is modeled as a random effect with prior

$$\boldsymbol{\beta}^c \mid \lambda_0 \sim \mathcal{N}(\mathbf{0}, \lambda_0^{-1} \mathbf{I}_K),$$

and

$$\lambda_0 \sim \text{Gam}(\lambda_0 \mid a_0, b_0),$$

we integrate over $\boldsymbol{\beta}^c$ and $\lambda_0$. This gives the conditional likelihood after IV selection:

$$\begin{aligned}
\mathcal{L}(\boldsymbol{\theta} \mid D_t) = \int \prod_{j=1}^{M_t} &\Pr\left(\hat{\gamma}_j, \hat{\boldsymbol{\gamma}}_j^c, \hat{\Gamma}_j \mid \left|\frac{\hat{\gamma}_j}{\hat{s}_{X,j}}\right| \geq t; \widehat{\boldsymbol{\Omega}}, \widehat{\mathbf{C}}, \hat{\mathbf{S}}_j, \ell_j, \boldsymbol{\theta}, \boldsymbol{\beta}^c\right) \\
&\times p(\boldsymbol{\beta}^c \mid \lambda_0) p(\lambda_0 \mid a_0, b_0)\, d\boldsymbol{\beta}^c\, d\lambda_0.
\end{aligned}$$

Substituting the conditional mixture density into the above expression yields

$$\begin{aligned}
\mathcal{L}(\boldsymbol{\theta} \mid D_t) = \int \prod_{j=1}^{M_t} \Bigg[ &\pi_t \frac{\mathcal{N}\left(\begin{pmatrix}\hat{\gamma}_j \\ \hat{\boldsymbol{\gamma}}_j^c \\ \hat{\Gamma}_j\end{pmatrix} \middle| \mathbf{0}, \ell_j \mathbf{A}_\beta \boldsymbol{\Sigma} \mathbf{A}_\beta^T + \ell_j \widehat{\boldsymbol{\Omega}} + \hat{\mathbf{S}}_j \widehat{\mathbf{C}} \hat{\mathbf{S}}_j\right)}{2\widehat{\mathcal{T}}_{1,j}} \\
&+ (1 - \pi_t) \frac{\mathcal{N}\left(\begin{pmatrix}\hat{\gamma}_j \\ \hat{\boldsymbol{\gamma}}_j^c \\ \hat{\Gamma}_j\end{pmatrix} \middle| \mathbf{0}, \ell_j \widehat{\boldsymbol{\Omega}} + \hat{\mathbf{S}}_j \widehat{\mathbf{C}} \hat{\mathbf{S}}_j\right)}{2\widehat{\mathcal{T}}_{0,j}} \Bigg] \\
&\times p(\boldsymbol{\beta}^c \mid \lambda_0) p(\lambda_0 \mid a_0, b_0)\, d\boldsymbol{\beta}^c\, d\lambda_0.
\end{aligned}$$

This likelihood differs from the ordinary likelihood in Eq. (6) by the selection normalizing constants $2\widehat{\mathcal{T}}_{1,j}$ and $2\widehat{\mathcal{T}}_{0,j}$, corresponding to the cases $Z_j = 1$ and $Z_j = 0$, respectively. These terms account for the fact that the observed SNPs in $D_t$ are selected based on their exposure association statistics, and therefore correct the winner's curse induced by exposure-based IV selection.

#### 1.3 The variational EM algorithm

For ease of reference, we first recall the notation used in MRMU. Let $K$ denote the number of measured covariates, let $\beta$ denote the causal effect of the exposure on the outcome, and let

$$\boldsymbol{\beta}^c = (\beta_1^c, \ldots, \beta_K^c)^\top$$

denote the effects of the measured covariates on the outcome.

Let

$$\mathbf{V}_j = \ell_j \widehat{\boldsymbol{\Omega}} + \hat{\mathbf{S}}_j \widehat{\mathbf{C}} \hat{\mathbf{S}}_j, \qquad \mathbf{R}_j = \mathbf{V}_j^{-1}.$$

We write

$$\boldsymbol{\Sigma} = \begin{pmatrix} \boldsymbol{\Sigma}_X & \mathbf{0}_{(K+1)\times 1} \\ \mathbf{0}_{1\times(K+1)} & \tau^2 \end{pmatrix},$$

where $\boldsymbol{\Sigma}_X$ is the covariance matrix of $(\gamma_j, \boldsymbol{\gamma}_j^{c\top})^\top$.

Define

$$\mathbf{A}(\beta, \boldsymbol{\beta}^c) = \mathbf{I}_{K+2} + \beta \mathbf{A}_1 + \sum_{k=1}^{K} \beta_k^c \mathbf{A}_{k+1}, \qquad \mathbf{A}_k = \mathbf{e}_{K+2} \mathbf{e}_k^\top, \quad k = 1, \ldots, K+1,$$

where $\mathbf{e}_{K+2}$ is the $(K+2)$-dimensional vector with the last element equal to one and all other elements equal to zero, and $\mathbf{e}_k$ is the $k$-th canonical basis vector in $\mathbb{R}^{K+2}$. Thus,

$$\mathbf{A}(\beta, \boldsymbol{\beta}^c) \begin{pmatrix} \gamma_j^* \\ \boldsymbol{\gamma}_j^{c*} \\ \alpha_j^* \end{pmatrix} = \begin{pmatrix} \gamma_j^* \\ \boldsymbol{\gamma}_j^{c*} \\ \beta\gamma_j^* + (\boldsymbol{\beta}^c)^\top \boldsymbol{\gamma}_j^{c*} + \alpha_j^* \end{pmatrix}.$$

The latent variables are

$$\mathcal{Z} = \left\{ Z_j, \gamma_j^*, \boldsymbol{\gamma}_j^{c*}, \alpha_j^* : j = 1, \ldots, M_t \right\} \cup \left\{ \boldsymbol{\beta}^c, \lambda_0 \right\},$$

and the unknown model parameters are

$$\boldsymbol{\theta} = \{\beta, \boldsymbol{\Sigma}_X, \tau^2, \pi_t\}.$$

Here $Z_j \in \{0, 1\}$ indicates whether SNP $j$ belongs to the component with nonzero IV strength after IV selection. We use $\widehat{\mathcal{T}}_{1,j}$ and $\widehat{\mathcal{T}}_{0,j}$ to denote the selection-rule normalizing constants for the cases $Z_j = 1$ and $Z_j = 0$, respectively:

$$\widehat{\mathcal{T}}_{1,j} = \Phi\left( \frac{-t\hat{s}_{X,j}}{\sqrt{\ell_j \sigma_X^2 + \ell_j \widehat{\Omega}_{11} + \widehat{C}_{11} \hat{s}_{X,j}^2}} \right), \qquad \widehat{\mathcal{T}}_{0,j} = \Phi\left( \frac{-t\hat{s}_{X,j}}{\sqrt{\ell_j \widehat{\Omega}_{11} + \widehat{C}_{11} \hat{s}_{X,j}^2}} \right).$$

Based on Eqs. (6) and (7) of the main text, the evidence lower bound is

$$\begin{aligned}\mathcal{L}(q_{\mathcal{Z}|D_t}, \boldsymbol{\theta}) &= \int \log \frac{\Pr\left(\left\{\hat{\gamma}_j, \widehat{\boldsymbol{\gamma}}_j^c, \hat{\Gamma}_j\right\}_{j=1}^{M_t}, \mathcal{Z} \mid D_t; \boldsymbol{\theta}\right)}{q(\mathcal{Z} \mid D_t)} q(\mathcal{Z} \mid D_t)\, d\mathcal{Z} \\ &= \mathbb{E}_q\left[\log \Pr\left(\left\{\hat{\gamma}_j, \widehat{\boldsymbol{\gamma}}_j^c, \hat{\Gamma}_j\right\}_{j=1}^{M_t}, \mathcal{Z} \mid D_t; \boldsymbol{\theta}\right)\right] - \mathbb{E}_q\left[\log q(\mathcal{Z} \mid D_t)\right].\end{aligned} \tag{7}$$

The complete-data conditional log-likelihood is

$$\begin{aligned}&\log \Pr\left(\left\{\hat{\gamma}_j, \widehat{\boldsymbol{\gamma}}_j^c, \hat{\Gamma}_j\right\}_{j=1}^{M_t}, \mathcal{Z} \mid D_t; \boldsymbol{\theta}\right) \\ =& \sum_{j=1}^{M_t} \log \mathcal{N}\left(\begin{pmatrix}\hat{\gamma}_j \\ \widehat{\boldsymbol{\gamma}}_j^c \\ \hat{\Gamma}_j\end{pmatrix}; Z_j \mathbf{A}(\beta, \boldsymbol{\beta}^c)\begin{pmatrix}\gamma_j^* \\ \boldsymbol{\gamma}_j^{c*} \\ \alpha_j^*\end{pmatrix}, \mathbf{V}_j\right) \\ &+ \sum_{j=1}^{M_t} \log \mathcal{N}\left(\begin{pmatrix}\gamma_j^* \\ \boldsymbol{\gamma}_j^{c*} \\ \alpha_j^*\end{pmatrix}; \mathbf{0}, \ell_j \boldsymbol{\Sigma}\right) + \log \mathcal{N}\left(\boldsymbol{\beta}^c; \mathbf{0}, \lambda_0^{-1}\mathbf{I}_K\right) + \log \mathrm{Gam}(\lambda_0; a_0, b_0) \\ &+ \sum_{j=1}^{M_t}\left\{Z_j \log \pi_t + (1 - Z_j)\log(1 - \pi_t)\right\} - \sum_{j=1}^{M_t}\left\{Z_j \log(2\widehat{\mathcal{T}}_{1,j}) + (1 - Z_j)\log(2\widehat{\mathcal{T}}_{0,j})\right\}.\end{aligned} \tag{8}$$

Equivalently, after expanding the Gaussian and Gamma densities,

$$\begin{aligned}&\log \Pr\left(\left\{\hat{\gamma}_j, \widehat{\boldsymbol{\gamma}}_j^c, \hat{\Gamma}_j\right\}_{j=1}^{M_t}, \mathcal{Z} \mid D_t; \boldsymbol{\theta}\right) \\ =& -\frac{(K+2)M_t}{2}\log(2\pi) - \frac{1}{2}\sum_{j=1}^{M_t}\log\det(\mathbf{V}_j) \\ &- \frac{1}{2}\sum_{j=1}^{M_t}\left\{\begin{pmatrix}\hat{\gamma}_j \\ \widehat{\boldsymbol{\gamma}}_j^c \\ \hat{\Gamma}_j\end{pmatrix} - Z_j \mathbf{A}(\beta, \boldsymbol{\beta}^c)\begin{pmatrix}\gamma_j^* \\ \boldsymbol{\gamma}_j^{c*} \\ \alpha_j^*\end{pmatrix}\right\}^T \mathbf{R}_j \left\{\begin{pmatrix}\hat{\gamma}_j \\ \widehat{\boldsymbol{\gamma}}_j^c \\ \hat{\Gamma}_j\end{pmatrix} - Z_j \mathbf{A}(\beta, \boldsymbol{\beta}^c)\begin{pmatrix}\gamma_j^* \\ \boldsymbol{\gamma}_j^{c*} \\ \alpha_j^*\end{pmatrix}\right\} \\ &- \frac{(K+2)M_t}{2}\log(2\pi) - \frac{1}{2}\sum_{j=1}^{M_t}\log\det(\ell_j\boldsymbol{\Sigma}) \\ &- \frac{1}{2}\sum_{j=1}^{M_t}\begin{pmatrix}\gamma_j^* \\ \boldsymbol{\gamma}_j^{c*} \\ \alpha_j^*\end{pmatrix}^T (\ell_j\boldsymbol{\Sigma})^{-1}\begin{pmatrix}\gamma_j^* \\ \boldsymbol{\gamma}_j^{c*} \\ \alpha_j^*\end{pmatrix} \\ &- \frac{K}{2}\log(2\pi) + \frac{K}{2}\log\lambda_0 - \frac{\lambda_0}{2}(\boldsymbol{\beta}^c)^\top\boldsymbol{\beta}^c - \log\Gamma(a_0) + a_0\log b_0 + (a_0 - 1)\log\lambda_0 - b_0\lambda_0 \\ &+ \sum_{j=1}^{M_t}\left\{Z_j \log \pi_t + (1 - Z_j)\log(1 - \pi_t)\right\} - \sum_{j=1}^{M_t}\left\{Z_j \log(2\widehat{\mathcal{T}}_{1,j}) + (1 - Z_j)\log(2\widehat{\mathcal{T}}_{0,j})\right\}.\end{aligned} \tag{9}$$

The variational distribution is assumed to factorize as

$$q(\mathcal{Z} \mid D_t) = q(\lambda_0 \mid D_t) q(\boldsymbol{\beta}^c \mid D_t) \prod_{j=1}^{M_t} q(\gamma_j^*, \boldsymbol{\gamma}_j^{c*}, \alpha_j^* \mid Z_j, D_t) q(Z_j \mid D_t), \tag{10}$$

where

$$q(Z_j \mid D_t) = \omega_j^{Z_j}(1-\omega_j)^{1-Z_j}, \qquad \omega_j = q(Z_j = 1 \mid D_t). \tag{11}$$

**E step**

In the E-step, we update $q^*(\mathcal{Z} \mid D_t)$ and then evaluate the ELBO in Eq. (7). The optimal variational distribution has the same factorized form,

$$q^*(\mathcal{Z} \mid D_t) = q^*(\lambda_0 \mid D_t) q^*(\boldsymbol{\beta}^c \mid D_t) \prod_{j=1}^{M_t} q^*(\gamma_j^*, \boldsymbol{\gamma}_j^{c*}, \alpha_j^* \mid Z_j, D_t) q^*(Z_j \mid D_t). \tag{12}$$

**Update $q^*(\gamma_j^*, \boldsymbol{\gamma}_j^{c*}, \alpha_j^* \mid Z_j, D_t)$.** Keeping only the terms in Eq. (9) that depend on $(\gamma_j^*, \boldsymbol{\gamma}_j^{c*}, \alpha_j^*)$ gives

$$\begin{aligned} &\log q(\gamma_j^*, \boldsymbol{\gamma}_j^{c*}, \alpha_j^* \mid Z_j, D_t) \\ =&\mathbb{E}_{q(\boldsymbol{\beta}^c)}\Bigg[ -\frac{1}{2} \left\{ \begin{pmatrix} \hat{\gamma}_j \\ \widehat{\boldsymbol{\gamma}}_j^c \\ \hat{\Gamma}_j \end{pmatrix} - Z_j \mathbf{A}(\beta, \boldsymbol{\beta}^c) \begin{pmatrix} \gamma_j^* \\ \boldsymbol{\gamma}_j^{c*} \\ \alpha_j^* \end{pmatrix} \right\}^T \mathbf{R}_j \left\{ \begin{pmatrix} \hat{\gamma}_j \\ \widehat{\boldsymbol{\gamma}}_j^c \\ \hat{\Gamma}_j \end{pmatrix} - Z_j \mathbf{A}(\beta, \boldsymbol{\beta}^c) \begin{pmatrix} \gamma_j^* \\ \boldsymbol{\gamma}_j^{c*} \\ \alpha_j^* \end{pmatrix} \right\} \\ &\qquad - \frac{1}{2} \begin{pmatrix} \gamma_j^* \\ \boldsymbol{\gamma}_j^{c*} \\ \alpha_j^* \end{pmatrix}^T (\ell_j \boldsymbol{\Sigma})^{-1} \begin{pmatrix} \gamma_j^* \\ \boldsymbol{\gamma}_j^{c*} \\ \alpha_j^* \end{pmatrix} \Bigg] + \text{const}\,. \end{aligned} \tag{13}$$

Let

$$\overline{\mathbf{A}} = \mathbb{E}_q\{\mathbf{A}(\beta, \boldsymbol{\beta}^c)\} = \mathbf{A}(\beta, \boldsymbol{\mu}_{\beta^c}),$$

and

$$\mathbf{Q}_j = \mathbb{E}_q\left\{\mathbf{A}(\beta, \boldsymbol{\beta}^c)^T \mathbf{R}_j \mathbf{A}(\beta, \boldsymbol{\beta}^c)\right\}.$$

Then

$$\mathbf{Q}_j = \overline{\mathbf{A}}^T \mathbf{R}_j \overline{\mathbf{A}} + \sum_{k=1}^{K} \sum_{l=1}^{K} (\boldsymbol{\Sigma}_{\beta^c})_{kl} \mathbf{A}_{k+1}^T \mathbf{R}_j \mathbf{A}_{l+1}. \tag{14}$$

For $Z_j = 1$, Eq. (13) is a quadratic function of $(\gamma_j^*, \boldsymbol{\gamma}_j^{c*}, \alpha_j^*)$, and hence

$$q^*(\gamma_j^*, \boldsymbol{\gamma}_j^{c*}, \alpha_j^* \mid Z_j = 1, D_t) = \mathcal{N}(\boldsymbol{\mu}_j, \boldsymbol{\Lambda}_j^{-1}), \tag{15}$$

where

$$\boldsymbol{\Lambda}_j = \mathbf{Q}_j + (\ell_j \boldsymbol{\Sigma})^{-1}, \qquad \boldsymbol{\mu}_j = \boldsymbol{\Lambda}_j^{-1} \overline{\mathbf{A}}^T \mathbf{R}_j \begin{pmatrix} \hat{\gamma}_j \\ \widehat{\boldsymbol{\gamma}}_j^c \\ \hat{\Gamma}_j \end{pmatrix}. \tag{16}$$

For $Z_j = 0$, Eq. (13) reduces to

$$\log q(\gamma_j^*, \boldsymbol{\gamma}_j^{c*}, \alpha_j^* \mid Z_j = 0, D_t) = -\frac{1}{2} \begin{pmatrix} \gamma_j^* \\ \boldsymbol{\gamma}_j^{c*} \\ \alpha_j^* \end{pmatrix}^T (\ell_j \boldsymbol{\Sigma})^{-1} \begin{pmatrix} \gamma_j^* \\ \boldsymbol{\gamma}_j^{c*} \\ \alpha_j^* \end{pmatrix} + \text{const},$$

and hence

$$q^*(\gamma_j^*, \boldsymbol{\gamma}_j^{c*}, \alpha_j^* \mid Z_j = 0, D_t) = \mathcal{N}(\mathbf{0}, \ell_j \boldsymbol{\Sigma}). \tag{17}$$

In the implementation of the E-step, $\boldsymbol{\Sigma}$ in Eq. (17) is evaluated at the current value from the previous iteration, denoted by $\boldsymbol{\Sigma}^{(old)}$ when needed.

**Update** $q^*(\boldsymbol{\beta}^c \mid D_t)$**.** Define

$$\mathbf{M}_j = \mathbb{E}_q\left[\left(\begin{matrix}\gamma_j^*\\ \boldsymbol{\gamma}_j^{c*}\\ \alpha_j^*\end{matrix}\right)\left(\begin{matrix}\gamma_j^*\\ \boldsymbol{\gamma}_j^{c*}\\ \alpha_j^*\end{matrix}\right)^T\middle| Z_j=1, D_t\right] = \boldsymbol{\Lambda}_j^{-1} + \boldsymbol{\mu}_j\boldsymbol{\mu}_j^T.$$

Keeping only the terms that depend on $\boldsymbol{\beta}^c$ yields

$$\begin{aligned}\log q(\boldsymbol{\beta}^c \mid D_t) = &\sum_{j=1}^{M_t}\omega_j\boldsymbol{\mu}_j^T\left(\sum_{k=1}^{K}\beta_k^c\mathbf{A}_{k+1}\right)^T\mathbf{R}_j\left(\begin{matrix}\hat{\gamma}_j\\ \widehat{\boldsymbol{\gamma}}_j^c\\ \hat{\Gamma}_j\end{matrix}\right)\\ &-\sum_{j=1}^{M_t}\omega_j\,\mathrm{Tr}\left[(\mathbf{I}_{K+2}+\beta\mathbf{A}_1)^T\mathbf{R}_j\left(\sum_{k=1}^{K}\beta_k^c\mathbf{A}_{k+1}\right)\mathbf{M}_j\right]\\ &-\frac{1}{2}\sum_{j=1}^{M_t}\omega_j\,\mathrm{Tr}\left[\left(\sum_{k=1}^{K}\beta_k^c\mathbf{A}_{k+1}\right)^T\mathbf{R}_j\left(\sum_{l=1}^{K}\beta_l^c\mathbf{A}_{l+1}\right)\mathbf{M}_j\right]\\ &-\frac{1}{2}\mathbb{E}_q(\lambda_0)(\boldsymbol{\beta}^c)^\top\boldsymbol{\beta}^c + \text{const}.\end{aligned}\tag{18}$$

Equivalently,

$$\log q(\boldsymbol{\beta}^c \mid D_t) = \mathbf{g}^\top\boldsymbol{\beta}^c - \frac{1}{2}(\boldsymbol{\beta}^c)^\top\left(\mathbf{H}+\frac{a_\lambda}{b_\lambda}\mathbf{I}_K\right)\boldsymbol{\beta}^c + \text{const},\tag{19}$$

where $a_\lambda/b_\lambda = \mathbb{E}_q(\lambda_0)$,

$$\begin{aligned}g_k &= \sum_{j=1}^{M_t}\omega_j\left[\boldsymbol{\mu}_j^\top\mathbf{A}_{k+1}^T\mathbf{R}_j\left(\begin{matrix}\hat{\gamma}_j\\ \widehat{\boldsymbol{\gamma}}_j^c\\ \hat{\Gamma}_j\end{matrix}\right) - \mathrm{Tr}\left\{(\mathbf{I}_{K+2}+\beta\mathbf{A}_1)^T\mathbf{R}_j\mathbf{A}_{k+1}\mathbf{M}_j\right\}\right],\\ H_{kl} &= \sum_{j=1}^{M_t}\omega_j\,\mathrm{Tr}\left(\mathbf{A}_{k+1}^T\mathbf{R}_j\mathbf{A}_{l+1}\mathbf{M}_j\right),\qquad k,l=1,\ldots,K.\end{aligned}\tag{20}$$

Thus,

$$q^*(\boldsymbol{\beta}^c \mid D_t) = \mathcal{N}(\boldsymbol{\mu}_{\beta^c}, \boldsymbol{\Sigma}_{\beta^c}),\qquad \boldsymbol{\Sigma}_{\beta^c}^{-1} = \mathbf{H}+\frac{a_\lambda}{b_\lambda}\mathbf{I}_K,\qquad \boldsymbol{\mu}_{\beta^c} = \boldsymbol{\Sigma}_{\beta^c}\mathbf{g}.\tag{21}$$

For efficient computation,

$$\mathrm{Tr}(\mathbf{A}_{k+1}^T\mathbf{B}) = B_{K+2,k+1},$$

and

$$H_{kl}^{(j)} = \mathrm{Tr}\left(\mathbf{A}_{k+1}^T\mathbf{R}_j\mathbf{A}_{l+1}\mathbf{M}_j\right) = R_{j,K+2,K+2}(\mathbf{M}_j)_{l+1,k+1}.$$

**Update** $q^*(\lambda_0 \mid D_t)$**.** Keeping only terms that depend on $\lambda_0$ yields

$$\log q(\lambda_0 \mid D_t) = \left(a_0 - 1 + \frac{K}{2}\right)\log\lambda_0 - \left\{b_0 + \frac{1}{2}\mathbb{E}_q\left((\boldsymbol{\beta}^c)^\top\boldsymbol{\beta}^c\right)\right\}\lambda_0 + \text{const}.$$

Therefore,

$$q^*(\lambda_0 \mid D_t) = \mathrm{Gam}(\lambda_0; a_\lambda, b_\lambda),\qquad a_\lambda = a_0 + \frac{K}{2},\qquad b_\lambda = b_0 + \frac{1}{2}\mathrm{Tr}\left(\boldsymbol{\mu}_{\beta^c}\boldsymbol{\mu}_{\beta^c}^T + \boldsymbol{\Sigma}_{\beta^c}\right).\tag{22}$$

Thus,

$$\mathbb{E}_q(\lambda_0) = \frac{a_\lambda}{b_\lambda},\qquad \mathbb{E}_q(\log\lambda_0) = \psi(a_\lambda) - \log b_\lambda.$$

**Evaluate the ELBO at the current E-step.** The ELBO used to monitor the variational EM iterations is

$$\mathcal{L}(q_{\mathcal{Z}|D_t}, \boldsymbol{\theta}) = \mathbb{E}_q\left[\log\Pr\left(\left\{\hat{\gamma}_j, \widehat{\boldsymbol{\gamma}}_j^c, \hat{\Gamma}_j\right\}_{j=1}^{M_t}, \mathcal{Z} \mid D_t; \boldsymbol{\theta}\right)\right] - \mathbb{E}_q\left[\log q(\mathcal{Z} \mid D_t)\right].$$

The first term is

$$\begin{aligned}
&\mathbb{E}_q\left[\log\Pr\left(\left\{\hat{\gamma}_j, \widehat{\boldsymbol{\gamma}}_j^c, \hat{\Gamma}_j\right\}_{j=1}^{M_t}, \mathcal{Z} \mid D_t; \boldsymbol{\theta}\right)\right] \\
=& -\frac{(K+2)M_t}{2}\log(2\pi) - \frac{1}{2}\sum_{j=1}^{M_t}\log\det(\mathbf{V}_j) \\
&- \frac{1}{2}\sum_{j=1}^{M_t}\Bigg[(1-\omega_j)\begin{pmatrix}\hat{\gamma}_j \\ \widehat{\boldsymbol{\gamma}}_j^c \\ \hat{\Gamma}_j\end{pmatrix}^T \mathbf{R}_j \begin{pmatrix}\hat{\gamma}_j \\ \widehat{\boldsymbol{\gamma}}_j^c \\ \hat{\Gamma}_j\end{pmatrix} \\
&\qquad + \omega_j\left\{\begin{pmatrix}\hat{\gamma}_j \\ \widehat{\boldsymbol{\gamma}}_j^c \\ \hat{\Gamma}_j\end{pmatrix}^T \mathbf{R}_j \begin{pmatrix}\hat{\gamma}_j \\ \widehat{\boldsymbol{\gamma}}_j^c \\ \hat{\Gamma}_j\end{pmatrix} - 2\boldsymbol{\mu}_j^T\overline{\mathbf{A}}^T\mathbf{R}_j\begin{pmatrix}\hat{\gamma}_j \\ \widehat{\boldsymbol{\gamma}}_j^c \\ \hat{\Gamma}_j\end{pmatrix} + \mathrm{Tr}(\mathbf{Q}_j\mathbf{M}_j)\right\}\Bigg] \\
&- \frac{(K+2)M_t}{2}\log(2\pi) - \frac{1}{2}\sum_{j=1}^{M_t}\log\det(\ell_j\boldsymbol{\Sigma}) \\
&- \frac{1}{2}\sum_{j=1}^{M_t}\left[\omega_j\ell_j^{-1}\,\mathrm{Tr}(\boldsymbol{\Sigma}^{-1}\mathbf{M}_j) + (1-\omega_j)\,\mathrm{Tr}\left\{\boldsymbol{\Sigma}^{-1}\boldsymbol{\Sigma}^{(old)}\right\}\right] \\
&- \frac{K}{2}\log(2\pi) + \frac{K}{2}\{\psi(a_\lambda) - \log b_\lambda\} - \frac{1}{2}\frac{a_\lambda}{b_\lambda}\,\mathrm{Tr}\left(\boldsymbol{\mu}_{\beta^c}\boldsymbol{\mu}_{\beta^c}^T + \boldsymbol{\Sigma}_{\beta^c}\right) \\
&- \log\Gamma(a_0) + a_0\log b_0 + (a_0-1)\{\psi(a_\lambda) - \log b_\lambda\} - b_0\frac{a_\lambda}{b_\lambda} \\
&+ \sum_{j=1}^{M_t}\{\omega_j\log\pi_t + (1-\omega_j)\log(1-\pi_t)\} \\
&- \sum_{j=1}^{M_t}\left\{\omega_j\log(2\widehat{\mathcal{T}}_{1,j}) + (1-\omega_j)\log(2\widehat{\mathcal{T}}_{0,j})\right\}.
\end{aligned} \tag{23}$$

The entropy term is

$$\begin{aligned}
-\mathbb{E}_q\left[\log q(\mathcal{Z} \mid D_t)\right] =& \sum_{j=1}^{M_t}\omega_j\left\{\frac{1}{2}\log\det(\boldsymbol{\Lambda}_j^{-1}) + \frac{K+2}{2}(1+\log 2\pi)\right\} \\
&+ \sum_{j=1}^{M_t}(1-\omega_j)\left\{\frac{1}{2}\log\det(\ell_j\boldsymbol{\Sigma}^{(old)}) + \frac{K+2}{2}(1+\log 2\pi)\right\} \\
&- \sum_{j=1}^{M_t}\{\omega_j\log\omega_j + (1-\omega_j)\log(1-\omega_j)\} \\
&+ \frac{1}{2}\log\det(\boldsymbol{\Sigma}_{\beta^c}) + \frac{K}{2}(1+\log 2\pi) \\
&+ \log\Gamma(a_\lambda) - (a_\lambda - 1)\psi(a_\lambda) - \log b_\lambda + a_\lambda.
\end{aligned} \tag{24}$$

Equations (23)–(24) are used at each iteration after replacing the variational parameters by the current optimal values. In particular,

$$\boldsymbol{\Sigma}^{(old)} = \begin{pmatrix} \boldsymbol{\Sigma}_X^{(old)} & \mathbf{0}_{(K+1)\times 1} \\ \mathbf{0}_{1\times(K+1)} & \tau^{2(old)} \end{pmatrix}$$

is used in $q^*(\gamma_j^*, \boldsymbol{\gamma}_j^{c*}, \alpha_j^* \mid Z_j = 0, D_t)$ during the E-step, while $\boldsymbol{\Lambda}_j$, $\boldsymbol{\mu}_j$, $\boldsymbol{\mu}_{\beta^c}$, $\boldsymbol{\Sigma}_{\beta^c}$, $a_\lambda$, $b_\lambda$, and $\omega_j$ are updated using the current parameter values.

**Update $q^*(Z_j \mid D_t)$.** To obtain the optimal $\omega_j$, retain the terms in the ELBO that depend on $\omega_j$:

$$\begin{aligned} \mathcal{L}(\omega_j) =& \omega_j \left\{ \frac{1}{2}\boldsymbol{\mu}_j^\top \boldsymbol{\Lambda}_j \boldsymbol{\mu}_j + \frac{1}{2}\log\det(\boldsymbol{\Lambda}_j^{-1}) - \frac{1}{2}\log\det(\ell_j \boldsymbol{\Sigma}^{(old)}) + \log \pi_t - \log(2\widehat{\mathcal{T}}_{1,j}) \right\} \\ & + (1-\omega_j)\left\{ \log(1-\pi_t) - \log(2\widehat{\mathcal{T}}_{0,j}) \right\} \\ & - \omega_j \log \omega_j - (1-\omega_j)\log(1-\omega_j) + \text{const}\,. \end{aligned} \tag{25}$$

Setting $\partial \mathcal{L}(\omega_j)/\partial \omega_j = 0$ gives

$$\omega_j = \frac{1}{1+\exp(-b_j)}, \tag{26}$$

where

$$b_j = \frac{1}{2}\boldsymbol{\mu}_j^\top \boldsymbol{\Lambda}_j \boldsymbol{\mu}_j + \log\frac{\pi_t}{1-\pi_t} + \frac{1}{2}\log\frac{\det(\boldsymbol{\Lambda}_j^{-1})}{\det(\ell_j \boldsymbol{\Sigma}^{(old)})} - \log\frac{\widehat{\mathcal{T}}_{1,j}}{\widehat{\mathcal{T}}_{0,j}}. \tag{27}$$

**M step**

In the M-step, we maximize $\mathcal{L}(q_{\mathcal{Z}|D_t}, \boldsymbol{\theta})$ with respect to $\boldsymbol{\theta}$, holding the variational distribution obtained in the E-step fixed.

**Update $\beta$.** Let

$$\overline{\mathbf{A}}_c = \sum_{k=1}^{K} \mu_{\beta^c,k} \mathbf{A}_{k+1}.$$

Collecting terms that depend on $\beta$ gives

$$\begin{aligned} \mathcal{L}(\beta) =& \sum_{j=1}^{M_t} \omega_j \boldsymbol{\mu}_j^\top (\beta \mathbf{A}_1)^\top \mathbf{R}_j \begin{pmatrix} \hat{\gamma}_j \\ \widehat{\boldsymbol{\gamma}}_j^c \\ \hat{\Gamma}_j \end{pmatrix} \\ & - \frac{1}{2}\sum_{j=1}^{M_t} \omega_j \operatorname{Tr}\left[ (\beta\mathbf{A}_1)^\top \mathbf{R}_j \left( \beta \mathbf{A}_1 + \overline{\mathbf{A}}_c + \mathbf{I}_{K+2} \right) \mathbf{M}_j \right] + \text{const}\,. \end{aligned} \tag{28}$$

Taking the derivative with respect to $\beta$ and setting it to zero yields

$$\beta = \frac{\sum_{j=1}^{M_t} \omega_j \left[ \boldsymbol{\mu}_j^\top \mathbf{A}_1^\top \mathbf{R}_j \begin{pmatrix} \hat{\gamma}_j \\ \widehat{\boldsymbol{\gamma}}_j^c \\ \hat{\Gamma}_j \end{pmatrix} - \operatorname{Tr}\left\{ \mathbf{A}_1^\top \mathbf{R}_j (\mathbf{I}_{K+2} + \overline{\mathbf{A}}_c) \mathbf{M}_j \right\} \right]}{\sum_{j=1}^{M_t} \omega_j \operatorname{Tr}\left( \mathbf{A}_1^\top \mathbf{R}_j \mathbf{A}_1 \mathbf{M}_j \right)}. \tag{29}$$

**Update $\pi_t$.** The terms depending on $\pi_t$ are

$$\mathcal{L}(\pi_t) = \sum_{j=1}^{M_t} \{\omega_j \log \pi_t + (1-\omega_j)\log(1-\pi_t)\} .$$

Thus,

$$\pi_t = \frac{1}{M_t}\sum_{j=1}^{M_t} \omega_j. \tag{30}$$

**Update $\mathbf{\Sigma}_X$.** Partition

$$\boldsymbol{\mu}_j = \begin{pmatrix} \boldsymbol{\mu}_{X,j} \\ \mu_{\alpha,j} \end{pmatrix}, \qquad \mathbf{\Lambda}_j^{-1} = \begin{pmatrix} \mathbf{\Lambda}_{X,j}^{-1} & \mathbf{\Lambda}_{X\alpha,j}^{-1} \\ \mathbf{\Lambda}_{\alpha X,j}^{-1} & \sigma_{\alpha,j}^2 \end{pmatrix},$$

where $\boldsymbol{\mu}_{X,j}$ contains the components corresponding to $(\gamma_j^*, \boldsymbol{\gamma}_j^{c*\top})^\top$. Define

$$\mathbf{M}_{X,j} = \boldsymbol{\mu}_{X,j}\boldsymbol{\mu}_{X,j}^T + \mathbf{\Lambda}_{X,j}^{-1}.$$

The terms in the ELBO that depend on $\mathbf{\Sigma}_X$ are

$$\begin{aligned} \mathcal{L}(\mathbf{\Sigma}_X) = &-\frac{M_t}{2}\log\det(\mathbf{\Sigma}_X) - \frac{1}{2}\sum_{j=1}^{M_t}\omega_j \ell_j^{-1}\operatorname{Tr}(\mathbf{\Sigma}_X^{-1}\mathbf{M}_{X,j}) \\ &-\frac{1}{2}\sum_{j=1}^{M_t}(1-\omega_j)\operatorname{Tr}\left\{\mathbf{\Sigma}_X^{-1}\mathbf{\Sigma}_X^{(old)}\right\} - \sum_{j=1}^{M_t}\omega_j \log\widehat{\mathcal{T}}_{1,j} + \text{const}\,. \end{aligned} \tag{31}$$

If $t=0$, the truncation term does not depend on $\mathbf{\Sigma}_X$, and the update is

$$\mathbf{\Sigma}_X = \frac{1}{M_t}\sum_{j=1}^{M_t}\left\{\omega_j\ell_j^{-1}\mathbf{M}_{X,j} + (1-\omega_j)\mathbf{\Sigma}_X^{(old)}\right\}. \tag{32}$$

If $t \neq 0$, directly maximizing Eq. (31) is difficult because $\widehat{\mathcal{T}}_{1,j}$ depends on the first diagonal element of $\mathbf{\Sigma}_X$. We instead maximize the following lower bound:

$$\begin{aligned} \mathcal{L}(\mathbf{\Sigma}_X) \geq &-\frac{M_t}{2}\log\det(\mathbf{\Sigma}_X^{(old)}) - \frac{M_t}{2}\operatorname{Tr}\left\{(\mathbf{\Sigma}_X^{(old)})^{-1}(\mathbf{\Sigma}_X - \mathbf{\Sigma}_X^{(old)})\right\} \\ &-\frac{1}{2}\sum_{j=1}^{M_t}\operatorname{Tr}\left[\mathbf{\Sigma}_X^{-1}\left\{\omega_j\ell_j^{-1}\mathbf{M}_{X,j} + (1-\omega_j)\mathbf{\Sigma}_X^{(old)}\right\}\right] \\ &-\frac{1}{2}\sum_{j=1}^{M_t}\omega_j\ell_j t\hat{s}_{X,j}\frac{\dot{\widehat{\mathcal{T}}}_{1,j}^{(old)}}{\widehat{\mathcal{T}}_{1,j}^{(old)}}\left\{\ell_j\sigma_X^{2(old)} + \ell_j\widehat{\Omega}_{11} + \widehat{C}_{11}\hat{s}_{X,j}^2\right\}^{-3/2}\mathbf{e}_1^\top(\mathbf{\Sigma}_X - \mathbf{\Sigma}_X^{(old)})\mathbf{e}_1 + \text{const}, \end{aligned} \tag{33}$$

where $\mathbf{e}_1 = (1,0,\ldots,0)^\top \in \mathbb{R}^{K+1}$, $\sigma_X^{2(old)} = \mathbf{e}_1^\top\mathbf{\Sigma}_X^{(old)}\mathbf{e}_1$, and $\dot{\widehat{\mathcal{T}}}_{1,j}^{(old)}$ denotes the derivative of $\widehat{\mathcal{T}}_{1,j}$ evaluated at $\mathbf{\Sigma}_X^{(old)}$.

Let

$$\mathbf{S}_X = \sum_{j=1}^{M_t} \left\{ \omega_j \ell_j^{-1} \mathbf{M}_{X,j} + (1-\omega_j) \mathbf{\Sigma}_X^{(old)} \right\},$$

$$\mathbf{G}_X = \sum_{j=1}^{M_t} \omega_j \ell_j t \hat{s}_{X,j} \frac{\dot{\widehat{\mathcal{T}}}_{1,j}^{(old)}}{\widehat{\mathcal{T}}_{1,j}^{(old)}} \left\{ \ell_j \sigma_X^{2(old)} + \ell_j \widehat{\Omega}_{11} + \widehat{C}_{11} \hat{s}_{X,j}^2 \right\}^{-3/2} \mathbf{e}_1 \mathbf{e}_1^\top. \tag{34}$$

Taking the derivative of the lower bound with respect to $\mathbf{\Sigma}_X$ gives

$$-\frac{M_t}{2} (\mathbf{\Sigma}_X^{(old)})^{-1} + \frac{1}{2} \mathbf{\Sigma}_X^{-1} \mathbf{S}_X \mathbf{\Sigma}_X^{-1} - \frac{1}{2} \mathbf{G}_X = \mathbf{0}. \tag{35}$$

Equivalently,

$$\mathbf{\Sigma}_X^{-1} \mathbf{S}_X \mathbf{\Sigma}_X^{-1} = M_t (\mathbf{\Sigma}_X^{(old)})^{-1} + \mathbf{G}_X.$$

Let

$$\mathbf{B}_X = M_t (\mathbf{\Sigma}_X^{(old)})^{-1} + \mathbf{G}_X,$$

and let $\mathbf{L}$ be the Cholesky factor such that $\mathbf{B}_X = \mathbf{L}\mathbf{L}^\top$. Then the update is

$$\mathbf{\Sigma}_X = \mathbf{L}^{-\top} \left( \mathbf{L}^\top \mathbf{S}_X \mathbf{L} \right)^{1/2} \mathbf{L}^{-1}. \tag{36}$$

**Update $\tau^2$.** The terms depending on $\tau^2$ are

$$\mathcal{L}(\tau^2) = \sum_{j=1}^{M_t} \left\{ -\frac{1}{2} \log(\tau^2) - \frac{1}{2} \omega_j \ell_j^{-1} \frac{\mu_{\alpha,j}^2 + \sigma_{\alpha,j}^2}{\tau^2} - \frac{1}{2} (1-\omega_j) \frac{\tau^{2(old)}}{\tau^2} \right\} + \text{const}. \tag{37}$$

Maximizing Eq. (37) gives

$$\tau^2 = \frac{1}{M_t} \sum_{j=1}^{M_t} \left\{ \omega_j \ell_j^{-1} (\mu_{\alpha,j}^2 + \sigma_{\alpha,j}^2) + (1-\omega_j) \tau^{2(old)} \right\}. \tag{38}$$

#### 1.4 Simulation settings

##### 1.4.1 Simulation I

We conducted simulations by generating summary statistics under an MVMR setting with one exposure variable, one measured covariate, and one outcome variable to investigate the performance of MRMU and other SVMR and MVMR methods. Because MVMR methods select IVs based on multiple exposures, we modified the inferential component of MRMU to include IVs for both variables. Specifically, we considered the following model for generating summary statistics of $M = 60{,}000$ SNPs for an exposure $X$, a measured covariate $X^c$, and an outcome $Y$:

$$\begin{pmatrix} \hat{\gamma}_j \\ \hat{\gamma}_j^c \\ \hat{\Gamma}_j \end{pmatrix} = \begin{pmatrix} \gamma_j \\ \gamma_j^c \\ \beta\gamma_j + \beta^c \gamma_j^c + \alpha_j \end{pmatrix} + \begin{pmatrix} u_j \\ u_j^c \\ v_j \end{pmatrix} + \begin{pmatrix} \epsilon_j \\ \epsilon_j^c \\ \xi_j \end{pmatrix}, \qquad j = 1, \cdots, M. \tag{39}$$

Here, $\beta$ is the causal effect of $X$ on $Y$ and is the parameter of interest, whereas $\beta^c$ is the effect of the measured covariate $X^c$ on $Y$. Similar to the MRMU model, we decomposed the estimated effects into the inferential model, the polygenic effect model, and the estimation error model. To account for unmeasured confounders, we adopted the same polygenic-effect and estimation-error assumptions as in Eq. (2) of main text and specified

$$\mathbf{\Omega} = \frac{h^2}{M}\begin{pmatrix} 1 & 0.3 & 0.3 \\ 0.3 & 1 & 0.3 \\ 0.3 & 0.3 & 1 \end{pmatrix}, \qquad \mathbf{C} = \begin{pmatrix} 1 & 0.15 & 0.15 \\ 0.15 & 1 & 0.15 \\ 0.15 & 0.15 & 1 \end{pmatrix}, \tag{40}$$

where $h^2/M$ is the per-SNP heritability of the polygenic effects, and we set $h^2 = 0.2$. The sample size was set to $N = 20{,}000$, so that $\hat{s}_{X,j} = \hat{s}^c_j = \hat{s}_{Y,j} = 1/\sqrt{N}$ for $j = 1, \cdots, M$.

For the inferential model, we considered two scenarios for the measured covariate $X^c$: (i) $X^c$ acts as a confounder of the relationship between $X$ and $Y$; and (ii) $X^c$ acts as a mediator of the relationship between $X$ and $Y$. Let $\tilde{\gamma}_j$ and $\tilde{\gamma}^c_j$ denote the direct effects of SNP $G_j$ on $X$ and $X^c$, respectively.

Under the confounder scenario, we assume that $X^c$ causally affects $X$. Let $\theta$ denote the effect of $X^c$ on $X$. Then the marginal SNP effects on $X$ and $X^c$ are given by

$$\gamma_j = \tilde{\gamma}_j + \theta\tilde{\gamma}^c_j, \qquad \gamma^c_j = \tilde{\gamma}^c_j. \tag{41}$$

Under the mediator scenario, we assume that $X$ causally affects $X^c$. Let $\theta_m$ denote the effect of $X$ on $X^c$. Then the marginal SNP effects on $X$ and $X^c$ are given by

$$\gamma_j = \tilde{\gamma}_j, \qquad \gamma^c_j = \tilde{\gamma}^c_j + \theta\tilde{\gamma}_j. \tag{42}$$

In both scenarios, the outcome summary statistics were generated according to Eq. (39), where $\beta$ denotes the direct effect of $X$ on $Y$ conditional on $X^c$, and $\beta^c$ denotes the effect of $X^c$ on $Y$. Under the mediator scenario, the total effect of $X$ on $Y$ is therefore $\beta + \theta\beta^c$.

To generate $(\gamma_j, \gamma^c_j)$, we set $\theta = 0.4$ in both scenarios and made the following distributional assumptions on the direct effects $(\tilde{\gamma}_j, \tilde{\gamma}^c_j)$: (1) $\tilde{\gamma}_j \sim \mathcal{N}\left(0, 20\frac{h^2}{M}\right), \tilde{\gamma}^c_j \sim \mathcal{N}\left(0, 20\frac{h^2}{M}\right)$; (2) $\tilde{\gamma}_j \sim \mathcal{N}\left(0, 20\frac{h^2}{M}\right), \tilde{\gamma}^c_j = 0$; (3)$\tilde{\gamma}_j = 0, \tilde{\gamma}^c_j \sim \mathcal{N}\left(0, 20\frac{h^2}{M}\right)$; (4) $\tilde{\gamma}_j = 0, \tilde{\gamma}^c_j = 0$. Here, the variances of the direct effects of IVs on $X$ and $X^c$ were set to 20 times the polygenic variance. We set the proportions of the four SNP groups as $q_1 = 0.003, q_2 = 0.003, q_3 = 0.003, q_4 = 0.991$. We generated $\alpha_j$, the direct effect on $Y$, from $\alpha_j \sim \mathcal{N}\left(0, \frac{h^2}{M}\right)$. The LD scores for the $M$ SNPs were randomly sampled from the European 1000 Genomes LD score reference panel. Because MRMU and the compared methods use GWAS summary statistics of approximately independent SNPs as input, each of the $M$ simulated SNPs was assumed to represent an LD block, and thus no LD was explicitly modeled among simulated SNPs. To evaluate type I error control, power, and point estimation, we fixed $\beta^c = 0.3$ and varied $\beta \in \{0, 0.1, 0.2, 0.3\}$.

We compared the performance of MRMU with six SVMR methods (IVW, Egger, RAPS, CAUSE, Weighted-mode, and Weighted-median) and five MVMR methods (MVMR-IVW, MVMR-Egger, MVMR-Median, MVMR-Lasso, and MVMR-Robust). Before conducting MR analysis, all of these methods required IV selection. For the SVMR methods that require strong IVs, including IVW, Egger, RAPS, Weighted-mode, and Weighted-median, we employed a $p$-value threshold of $5 \times 10^{-8}$ on the summary statistics of $X$ to select SNPs significantly associated with $X$. For CAUSE, we used the default IV threshold of $1 \times 10^{-3}$. For MRMU, we specified an IV threshold of $5 \times 10^{-8}$. To select IVs for the five MVMR methods, we set the IV threshold at $5 \times 10^{-8}$ and took the union of IVs selected by both $X$ and $X^c$.

##### 1.4.2 Simulation II

We conducted simulations to evaluate the performance of MRMU in scenarios involving multiple measured covariates. We considered a case with $K = 9$ measured covariates. To simulate realistic settings in which not all adjusted covariates are truly relevant to the outcome, we set $\boldsymbol{\beta}^c = (0.1,\ 0.2,\ 0.2,\ 0.05,\ 0,\ 0,\ 0,\ 0,\ 0)^T$, so that five of the nine measured covariates had no effect on the outcome. We generated summary statistics for $M = 60{,}000$ SNPs under the MRMU model in Eq. (6). We first specified the correlation matrices for the polygenic effects $(u_j, \boldsymbol{u}_j^c, v_j)$ and the inferential-model effects $(\gamma_j, \boldsymbol{\gamma}_j^c, \alpha_j)$, denoted by $\mathbf{R}_g$ and $\mathbf{R}$, respectively, together with the matrix $\mathbf{C}$ for estimation errors, using a real-data example (given below). Given $\mathbf{R}_g$, we set $\boldsymbol{\Omega} = \frac{h^2}{M}\mathbf{R}_g$, with $h^2 = 0.2$. Thus, the variances of the polygenic effects for the exposure, the measured covariates, and the outcome were all set to $h^2/M$. For the inferential model, we set the variance of $\gamma_j$ to be $\sigma_X^2 = 20\frac{h^2}{M}$, that is, 20 times the polygenic variance. The variance ratios for the IV effects on the nine measured covariates relative to the exposure were specified as $\left(\sigma_{c,1}^2/\sigma_X^2, \ldots, \sigma_{c,9}^2/\sigma_X^2\right) = (0.84,\ 0.3,\ 0.15,\ 0.53,\ 0.57,\ 1.34,\ 0.74,\ 0.39,\ 0.033)$. The variance of $\alpha_j$ was set to $\tau^2 = \frac{h^2}{M}$. Using these variances together with $\mathbf{R}$, we obtained the covariance matrix $\boldsymbol{\Sigma}$. We set $\pi = 0.5\%$, indicating that only a small fraction of SNPs had nonzero IV strength. For the estimation errors $(\epsilon_j, \boldsymbol{\epsilon}_j^c, \xi_j)$, we set $\hat{s}_{X,j}^2 = \hat{s}_{1,j}^{c\,2} = \cdots = \hat{s}_{9,j}^{c\,2} = \hat{s}_{Y,j}^2 = \frac{1}{N}$, where the GWAS sample size was set to $N = 20{,}000$. To examine point estimation, type I error control, and power, we varied $\beta \in \{0,\ 0.05,\ 0.1,\ 0.15,\ 0.2,\ 0.25,\ 0.3\}$.

$$\mathbf{R}_g = \begin{pmatrix}
1.00000 & 0.37416 & 0.04067 & 0.30328 & 0.29800 & 0.36778 & -0.58968 & -0.09359 & 0.41212 & -0.22587 & 0.30066 \\
0.37416 & 1.00000 & 0.02134 & 0.06685 & 0.21797 & 0.21104 & -0.00521 & -0.12116 & 0.15441 & -0.09099 & 0.26263 \\
0.04067 & 0.02134 & 1.00000 & -0.04067 & 0.08176 & 0.13363 & -0.01924 & -0.07651 & 0.07037 & -0.10472 & 0.34701 \\
0.30328 & 0.06685 & -0.04067 & 1.00000 & 0.25701 & 0.31457 & -0.37836 & -0.12083 & 0.39444 & -0.26327 & 0.29346 \\
0.29800 & 0.21797 & 0.08176 & 0.25701 & 1.00000 & 0.25084 & -0.18267 & -0.04685 & 0.07224 & -0.14488 & 0.26260 \\
0.36778 & 0.21104 & 0.13363 & 0.31457 & 0.25084 & 1.00000 & -0.30506 & -0.10325 & 0.31121 & -0.17655 & 0.33966 \\
-0.58968 & -0.00521 & -0.01924 & -0.37836 & -0.18267 & -0.30506 & 1.00000 & 0.00177 & -0.35368 & 0.22051 & -0.29435 \\
-0.09359 & -0.12116 & -0.07651 & -0.12083 & -0.04685 & -0.10325 & 0.00177 & 1.00000 & -0.08969 & 0.15594 & -0.13436 \\
0.41212 & 0.15441 & 0.07037 & 0.39444 & 0.07224 & 0.31121 & -0.35368 & -0.08969 & 1.00000 & -0.19402 & 0.29430 \\
-0.22587 & -0.09099 & -0.10472 & -0.26327 & -0.14488 & -0.17655 & 0.22051 & 0.15594 & -0.19402 & 1.00000 & -0.23521 \\
0.30066 & 0.26263 & 0.34701 & 0.29346 & 0.26260 & 0.33966 & -0.29435 & -0.13436 & 0.29430 & -0.23521 & 1.00000
\end{pmatrix}.$$

$$\mathbf{R} = \begin{pmatrix}
1.00000 & 0.42110 & 0.18732 & -0.12510 & 0.19101 & 0.26643 & -0.49825 & -0.17546 & 0.45505 & -0.09041 & 0.00000 \\
0.42110 & 1.00000 & 0.07417 & -0.14446 & 0.16235 & 0.22637 & 0.03890 & -0.16981 & 0.19071 & 0.02451 & 0.00000 \\
0.18732 & 0.07417 & 1.00000 & -0.17659 & 0.11468 & 0.26406 & -0.14688 & -0.26474 & 0.25588 & -0.08442 & 0.00000 \\
-0.12510 & -0.14446 & -0.17659 & 1.00000 & -0.09533 & -0.17314 & -0.00310 & 0.03171 & -0.05143 & -0.22237 & 0.00000 \\
0.19101 & 0.16235 & 0.11468 & -0.09533 & 1.00000 & 0.27208 & -0.16324 & -0.06140 & 0.27661 & -0.07723 & 0.00000 \\
0.26643 & 0.22637 & 0.26406 & -0.17314 & 0.27208 & 1.00000 & -0.17057 & -0.21920 & 0.29419 & -0.30654 & 0.00000 \\
-0.49825 & 0.03890 & -0.14688 & -0.00310 & -0.16324 & -0.17057 & 1.00000 & -0.01796 & -0.32957 & 0.13115 & 0.00000 \\
-0.17546 & -0.16981 & -0.26474 & 0.03171 & -0.06140 & -0.21920 & -0.01796 & 1.00000 & -0.11214 & 0.26812 & 0.00000 \\
0.45505 & 0.19071 & 0.25588 & -0.05143 & 0.27661 & 0.29419 & -0.32957 & -0.11214 & 1.00000 & -0.16429 & 0.00000 \\
-0.09041 & 0.02451 & -0.08442 & -0.22237 & -0.07723 & -0.30654 & 0.13115 & 0.26812 & -0.16429 & 1.00000 & 0.00000 \\
0.00000 & 0.00000 & 0.00000 & 0.00000 & 0.00000 & 0.00000 & 0.00000 & 0.00000 & 0.00000 & 0.00000 & 1.00000
\end{pmatrix}.$$

$$\mathbf{C} = \begin{pmatrix}
1.16853 & 0.43815 & 0.07295 & 0.32301 & 0.19418 & 0.32595 & -0.56178 & -0.04885 & 0.35967 & -0.07028 & 0.05594 \\
0.43815 & 1.13243 & 0.04848 & 0.12197 & 0.10066 & 0.16111 & -0.00989 & -0.04966 & 0.12270 & -0.02211 & 0.02217 \\
0.07295 & 0.04848 & 1.19669 & -0.01197 & 0.02428 & 0.08161 & -0.00082 & -0.06145 & 0.09800 & -0.04720 & 0.06147 \\
0.32301 & 0.12197 & -0.01197 & 1.26122 & 0.17997 & 0.26272 & -0.33783 & -0.12236 & 0.46705 & -0.11981 & 0.09026 \\
0.19418 & 0.10066 & 0.02428 & 0.17997 & 1.20608 & 0.14194 & -0.18765 & -0.02331 & 0.03954 & -0.05970 & 0.07906 \\
0.32595 & 0.16111 & 0.08161 & 0.26272 & 0.14194 & 1.11483 & -0.18422 & -0.04681 & 0.27459 & -0.02929 & 0.01577 \\
-0.56178 & -0.00989 & -0.00082 & -0.33783 & -0.18765 & -0.18422 & 1.17670 & -0.01237 & -0.23411 & 0.06716 & -0.09941 \\
-0.04885 & -0.04966 & -0.06145 & -0.12236 & -0.02331 & -0.04681 & -0.01237 & 1.86435 & -0.06001 & 0.16470 & -0.05573 \\
0.35967 & 0.12270 & 0.09800 & 0.46705 & 0.03954 & 0.27459 & -0.23411 & -0.06001 & 1.30434 & -0.08385 & 0.08913 \\
-0.07028 & -0.02211 & -0.04720 & -0.11981 & -0.05970 & -0.02929 & 0.06716 & 0.16470 & -0.08385 & 1.14353 & -0.05561 \\
0.05594 & 0.02217 & 0.06147 & 0.09026 & 0.07906 & 0.01577 & -0.09941 & -0.05573 & 0.08913 & -0.05561 & 1.01368
\end{pmatrix}.$$

#### IV selection and LD clumping across MR methods

IV selection was performed according to the modeling framework and recommended settings of each MR method. For MRMU and SVMR methods, IVs were selected based on their associations with the exposure of interest, because these methods estimate the causal effect of one exposure at a time. For MVMR methods, IVs were selected by taking the union of variants associated with any exposure included in the multivariable model, including the primary exposure and measured covariates.

For MRMU and MRAPSS, IVs were selected using a suggestive genome-wide threshold of $P < 5 \times 10^{-5}$. For standard SVMR methods, including IVW, MR-Egger, RAPS, weighted median, and weighted mode, IVs were selected using the conventional genome-wide significance threshold of $P < 5 \times 10^{-8}$. For CAUSE, IVs were selected using $P < 1 \times 10^{-3}$, following its recommended setting. For MVMR methods, IVs were selected at $P < 5 \times 10^{-8}$ for each exposure or measured covariate, and the union of these variants was used as the IV set.

To ensure approximate independence among selected IVs, LD clumping was applied using PLINK. Unless otherwise specified by the software or method recommendation, clumping was performed with an LD threshold of $r^2 < 0.01$ within a 1 Mb window. CAUSE was run using its recommended clumping setting with $r^2 < 0.01$. The same IV selection and clumping strategy was applied consistently across simulations, negative control analyses, and real-data applications, unless otherwise stated.

#### 1.5 Real-data negative control analysis

We conducted a real-data negative control analysis using adult body mass index (adult BMI) as the exposure and height at age 10 as the outcome. GWAS summary statistics for adult BMI were obtained from a large meta-analysis of the GIANT consortium and UK Biobank, including 681,275 participants [2]. This dataset represents a meta-analysis of GIANT studies in approximately 250,000 participants and a GWAS of BMI in approximately 450,000 UK Biobank participants. The dataset is representative of adult BMI because the mean age of GIANT participants was 55.5 years, and UK Biobank participants were aged 37–73 years, with 99.5% between 40 and 69 years [2, 3].

Height at age 10 was a self-reported UK Biobank trait measuring participants' height relative to their peers at age 10 (Phenotype code: 1697; $N = 332,021$), with GWAS summary statistics obtained from the Neale Lab UK Biobank resource.

We identified 13 complex traits as candidate measured covariates representing potential pleiotropic pathways in this negative control analysis. These traits were grouped into four domains: socioeconomic factors, including educational attainment and household income; diet-related traits, including fat, protein, carbohydrate, and sugar intake; pubertal timing and reproductive-related traits, including age at voice break, age at menarche, age at first birth, and age at first sex; and lifestyle behaviors, including alcohol consumption, smoking initiation age, and number of sexual partners. These covariates were selected to capture plausible measured pathways that may link adult BMI-associated variants to childhood height, despite the absence of a plausible direct effect of adult BMI on height at age 10. GWAS summary statistics for these covariates were obtained from UK Biobank or external consortia, with details summarized in Supplementary Table S2. We applied the same preprocessing and harmonization procedures to all GWAS summary statistics as in previous studies [1, 4].

#### 1.6 MRMU workflow for the CAD real-data analysis

We applied MRMU to investigate potential causal risk factors for coronary artery disease (CAD), and the overall analytical workflow is illustrated in Supplementary Figure ??. First, we conducted a comprehensive literature review and collected 88 complex traits as candidate risk factors for CAD. These traits were broadly grouped into multiple categories, including autoimmune diseases, blood cell traits, socioeconomic status, reproductive traits, gastrointestinal diseases, cancer, mental health and brain development, anthropometric traits, other diseases, and cardiometabolic traits. The cardiometabolic category included biomarkers, Lifestyle, and metabolic traits. Detailed information on the GWAS sources for all candidate traits is provided in Supplementary Table S3.

For the outcome, we used CAD GWAS summary statistics from the UK Biobank (UKB; 60,801 cases and 123,504 controls) in the discovery stage. After collecting the GWAS summary statistics for both candidate risk factors and the outcome, we performed standard data preprocessing. Specifically, we conducted SNP-level quality control by removing variants that were not in HapMap3, duplicated SNPs, SNPs with missing information, variants in the major histocompatibility complex region, ambiguous SNPs, poorly imputed SNPs, and SNPs with minor allele frequency less than 0.01. We then harmonized the summary statistics so that the SNP effect estimates for each risk factor and the outcome were aligned to the same effect allele.

Next, we performed a prescreening analysis to prioritize candidate risk factors for downstream MRMU analysis. For each candidate trait, instrumental variables were selected using a GWAS significance threshold of $P < 5 \times 10^{-5}$, followed by LD clumping in PLINK with $r^2 = 0.001$ and a 1 Mb window to ensure independence among instruments. We then applied MRAPSS to evaluate the association between each candidate trait and CAD, and traits with an MRAPSS test $P$-value less than 0.05 were retained for further analysis. This prescreening step resulted in 37 candidate risk factors for downstream MRMU analysis.

We subsequently applied MRMU to estimate the causal effects of the prescreened risk factors on CAD. In this analysis, cardiometabolic traits were included as measured covariates to account for potential confounding from established metabolic and lifestyle-related pathways relevant to CAD. For each target risk factor, MRMU was fitted while adjusting for these measured covariates, thereby improving the robustness of causal effect estimation.

Finally, to validate the findings from the discovery stage, we conducted replication analyses using two independent CAD GWAS datasets, including CARDIoGRAMplusC4D and a CAD Meta-Analysis (122,733 cases and 424,528 controls). Consistency of the results across the discovery and replication datasets was used to assess the robustness of the identified causal relationships. For analyses involving multiple outcome datasets, All GWAS summary statistics were first subjected to standard preprocessing and harmonization before IV selection. Specifically, effect alleles were aligned across exposure, measured covariate, and outcome GWAS datasets, and variants with ambiguous alleles, incompatible allele coding, or missing required summary statistics were removed. IVs were then selected from the harmonized GWAS data according to the method-specific rules described above. This preprocessing order ensured that, for a given exposure, the same set of IVs was used across different CAD outcome datasets whenever the variants were available in all harmonized datasets. Therefore, differences in MRMU estimates across CAD datasets were not driven by changes in the IV set, but instead reflected differences in the outcome GWAS summary statistics.

#### 1.7 Definition of a defined difference in pathway-related covariate analyses

For each exposure, let $\hat{\beta}^{(0)}$ denote the MR-APSS estimate for the exposure effect on CAD without adjustment for measured covariates, and let $\hat{\beta}^{(g)}$ denote the MRMU estimate after adjustment for covariate group $g$. Let $\mathrm{se}^{(0)}$ and $\mathrm{se}^{(g)}$ denote the corresponding standard errors.

We defined the change in effect estimate after covariate adjustment as

$$\Delta^{(g)} = \hat{\beta}^{(g)} - \hat{\beta}^{(0)}.$$

A significant change in the estimated effect was assessed using

$$Z^{(g)} = \frac{\hat{\beta}^{(g)} - \hat{\beta}^{(0)}}{\sqrt{\left(\mathrm{se}^{(g)}\right)^2 + \left(\mathrm{se}^{(0)}\right)^2}},$$

with the corresponding two-sided $P$-value

$$P_{\Delta}^{(g)} = 2\left\{1 - \Phi\left(\left|Z^{(g)}\right|\right)\right\},$$

where $\Phi(\cdot)$ denotes the cumulative distribution function of the standard normal distribution.

An exposure–CAD pair was marked as having a "defined difference" after adjustment for covariate group $g$ if either of the following criteria was satisfied:

$$P_{\Delta}^{(g)} < 0.05,$$

or

$$P^{(0)} < 0.05 \quad \text{and} \quad P^{(g)} \geq 0.05,$$

where $P^{(0)}$ is the MR-APSS $P$-value and $P^{(g)}$ is the MRMU $P$-value after adjustment for covariate group $g$. These exposure–CAD pairs were marked by † in Fig. 5A–C.

Because MR-APSS and MRMU both use exposure-selected IVs, this comparison was designed to evaluate changes associated with measured covariate adjustment while reducing the influence of differences in IV selection. The test above treats the two estimates as approximately independent and is used as a diagnostic criterion for identifying covariate-dependent changes, rather than as a formal mediation test.

### Supplementary figures

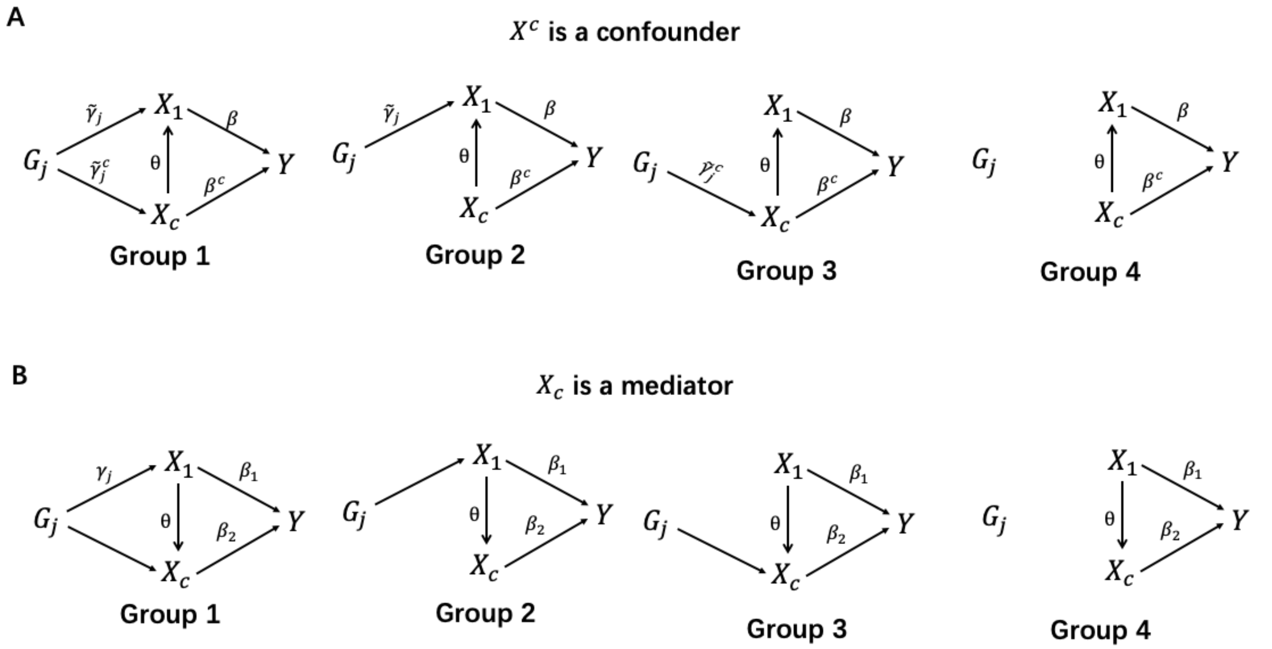

**Figure** S1: **Four-group IV simulation model with measured covariates.** (**A**) $X^c$ acts as a measured confounder of the exposure–outcome relationship. (**B**) $X^c$ acts as a mediator on the pathway from the exposure $X$ to the outcome $Y$. In both settings, genetic variants were classified according to their direct effects on $X$ and $X^c$: Group 1 variants have direct effects on both $X$ and $X^c$; Group 2 variants have a direct effect on $X$ but no effect on $X^c$; Group 3 variants have a direct effect on $X^c$ and an indirect effect on $X$ through $X^c$; and Group 4 variants have no effect on either $X$ or $X^c$.

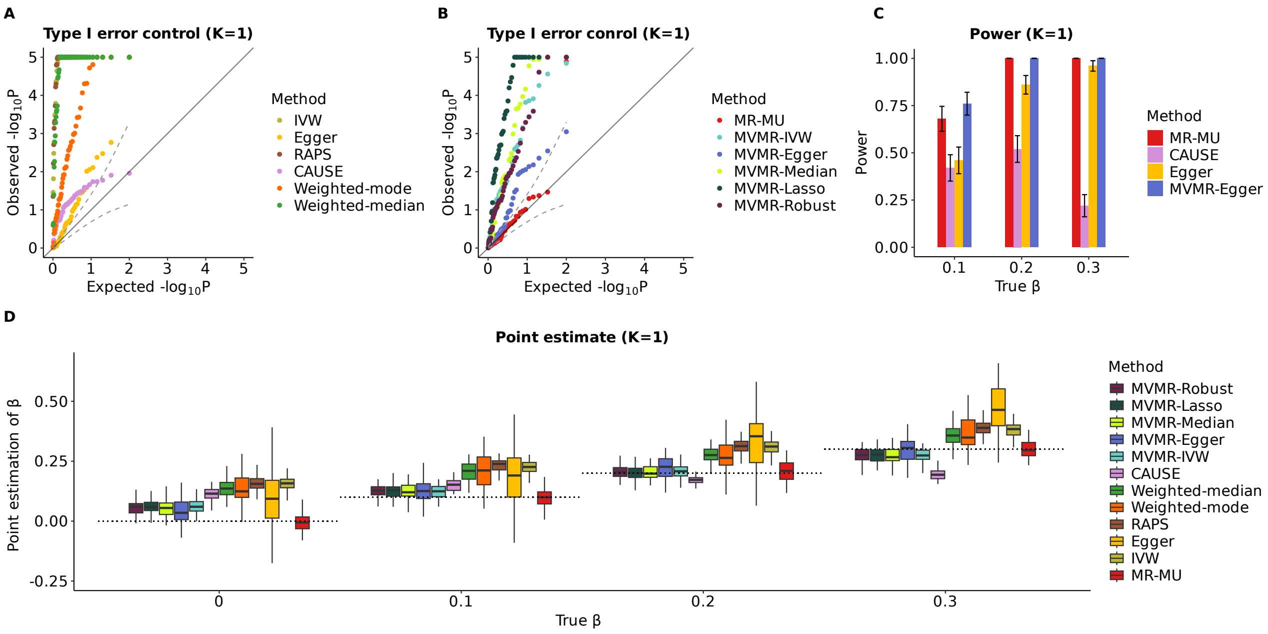

**Figure** S2: **Results under the simulation setting with one measured mediator** ($K = 1$) (**A**, **B**) Quantile–quantile plots of $-\log_{10}(P)$ values under the null setting ($\beta_1 = 0$) for MRMU and SVMR methods (**A**) and for MRMU and MVMR methods (**B**). MRMU, MR-Egger, CAUSE, and MVMR-Egger showed well-calibrated type I error. (**C**) Statistical power of MRMU, MR-Egger, CAUSE, and MVMR-Egger under alternative settings with $\beta_1 \in \{0.1, 0.2, 0.3\}$. (**D**) Causal effect estimates across methods under settings with $\beta_1 \in \{0, 0.1, 0.2, 0.3\}$; dotted horizontal lines indicate the true causal effects. Results for each setting were summarized from 50 replications.

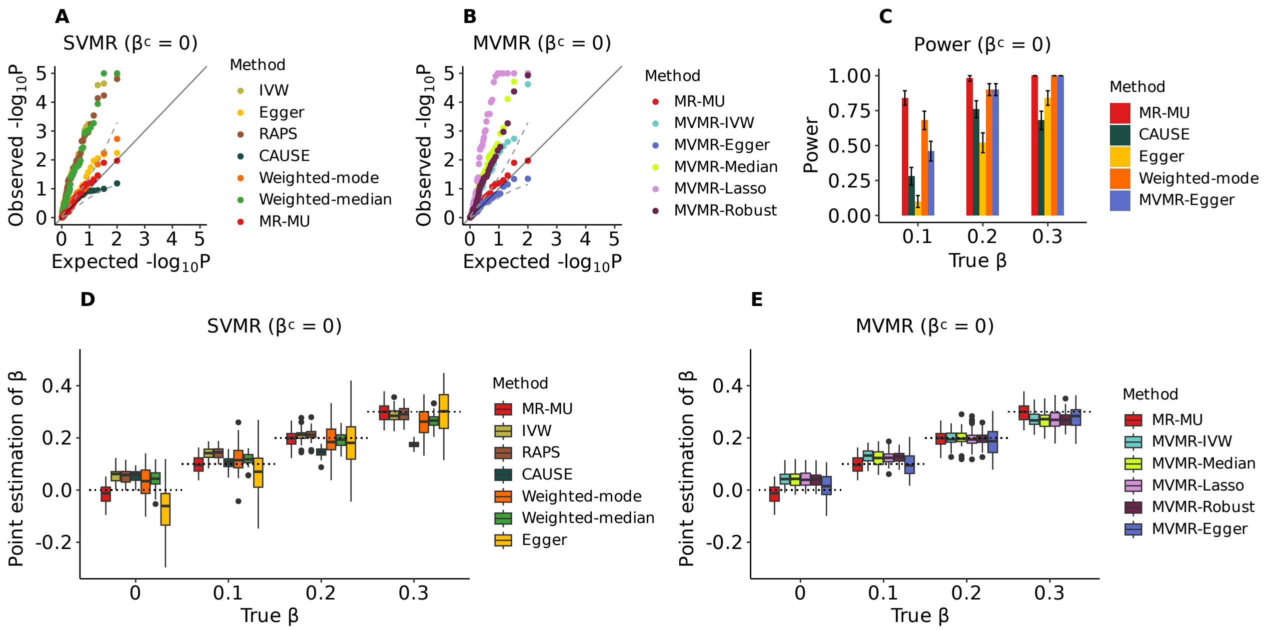

**Figure** S3: **Results under the simulation setting with one measured confounder with** $\beta^c = 0$ (**A**, **B**) Quantile–quantile plots of $-\log_{10}(P)$ values under the null setting ($\beta = 0$) for MRMU and SVMR methods (**A**) and for MRMU and MVMR methods (**B**). MRMU, MR-Egger, CAUSE, and MVMR-Egger showed well-calibrated type I error. (**C**) Statistical power of MRMU, MR-Egger, CAUSE, and MVMR-Egger under alternative settings with $\beta_1 \in \{0.1, 0.2, 0.3\}$. (**D-E**) Causal effect estimates across SVMR methods (**D**) and MVMR methods (**E**) under settings with $\beta_1 \in \{0, 0.1, 0.2, 0.3\}$; dotted horizontal lines indicate the true causal effects. Results for each setting were summarized from 50 replications.

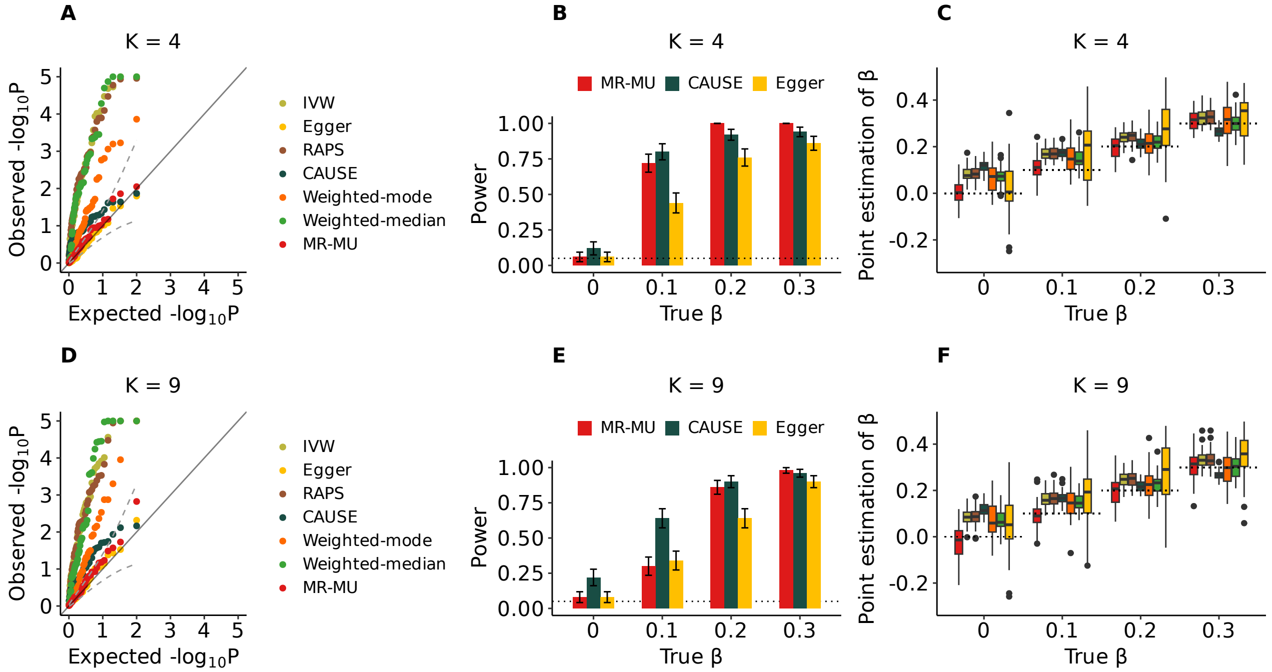

**Figure** S4: Performance of MRMU and SVMR methods in simulation settings with multiple measured covariates ($K = 4$ or $K = 9$). (**A**) Quantile–quantile plots of $-\log_{10}(P)$ values under the null setting with $K = 4$. (**B**) Statistical power of MRMU, CAUSE, and MR-Egger under alternative settings with the causal effect $\beta$ ranging from 0.05 to 0.3 for $K = 4$. (**C**) Causal effect estimates from MRMU and the compared SVMR methods under the setting with $K = 4$. (**D**) Quantile–quantile plots of $-\log_{10}(P)$ values under the null setting with $K = 9$. (**E**) Statistical power of MRMU, CAUSE, and MR-Egger under alternative settings with the causal effect $\beta$ ranging from 0.05 to 0.3 for $K = 9$. (**F**) Causal effect estimates from MRMU and the compared SVMR methods under the setting with $K = 9$. Results for each setting were summarized over 50 replications.

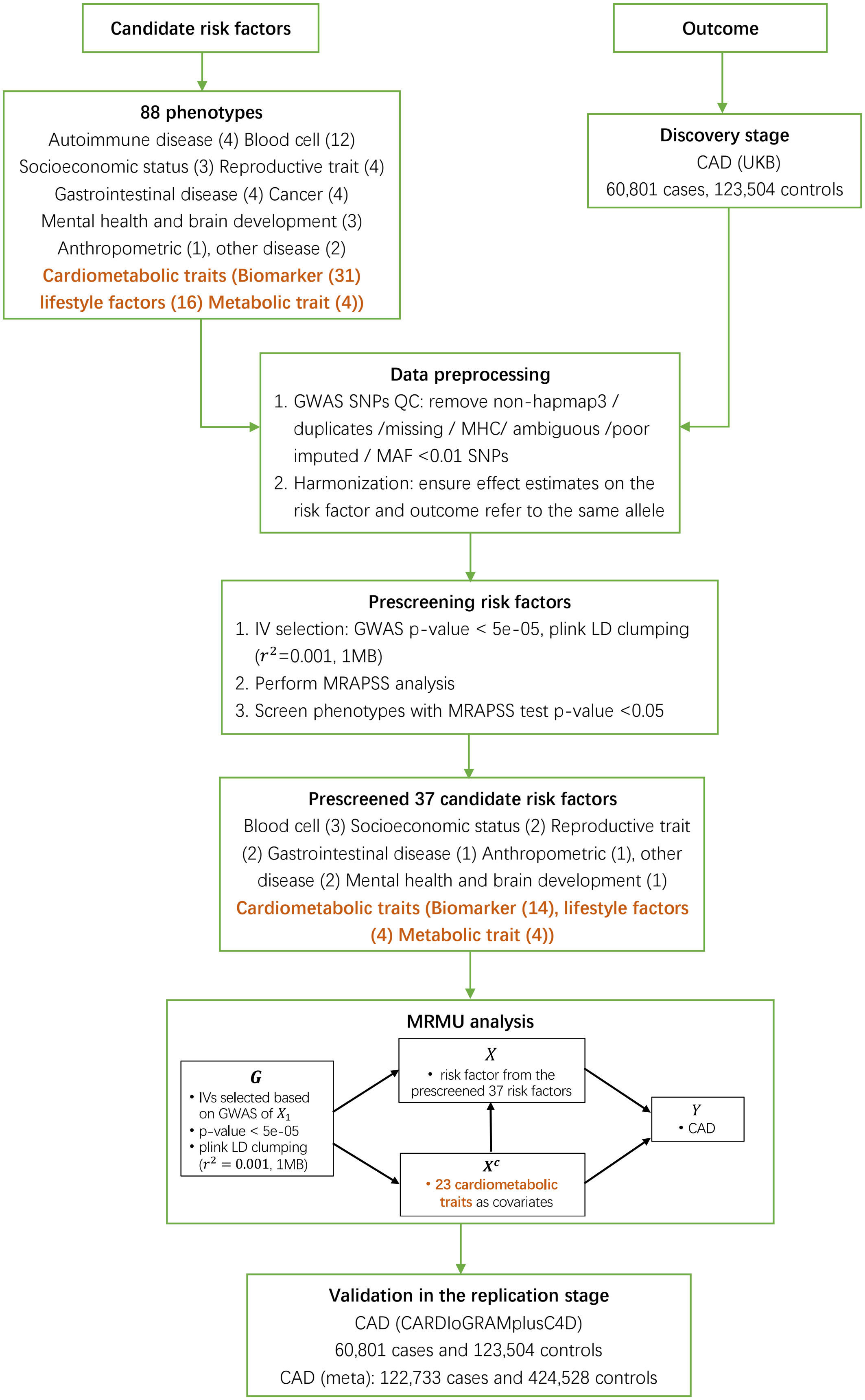

**Figure** S5: MRMU workflow for the CAD real-data analysis

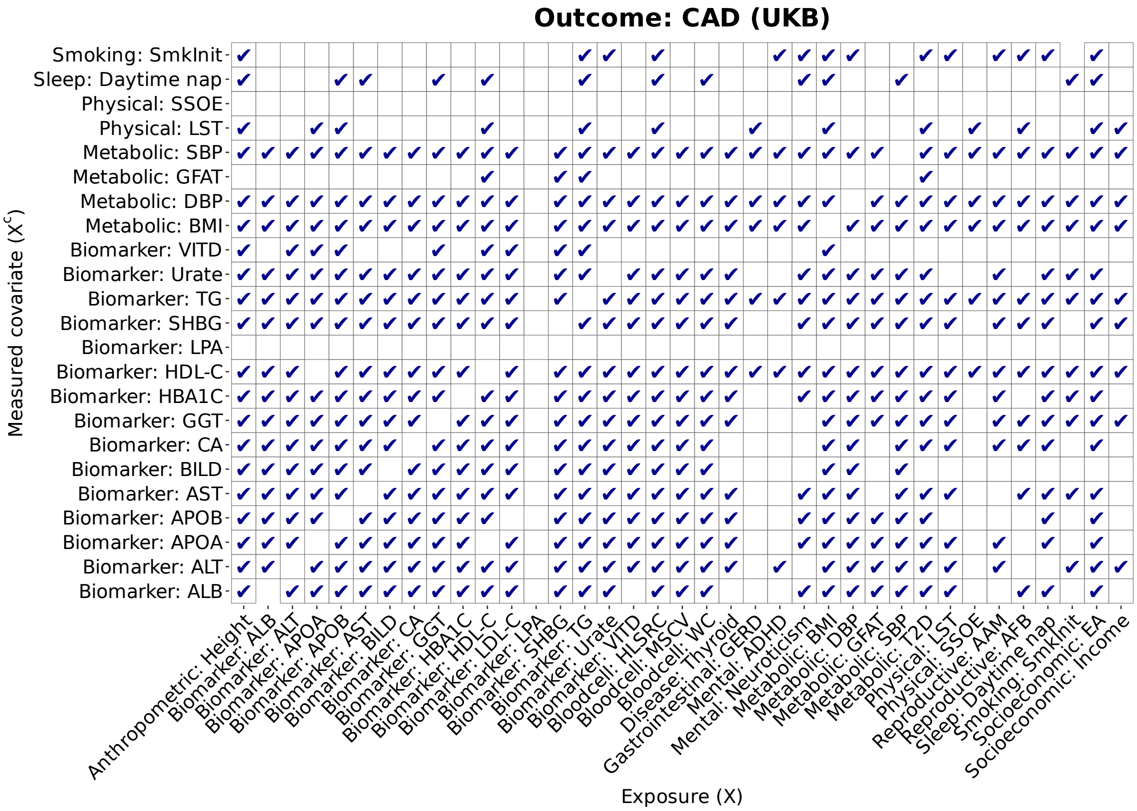

**Figure** S6: **Exposure-specific selection of measured covariates for MRMU analysis of CAD using the UK Biobank GWAS as the outcome dataset.** Columns represent candidate exposures and rows represent measured covariates. A check mark indicates that the corresponding covariate was retained for adjustment for a given exposure. For each exposure, IVs were first selected from the exposure GWAS, and each candidate covariate was evaluated by counting the number of exposure-selected IVs that reached genome-wide significance ($P < 5 \times 10^{-8}$) in the covariate GWAS. Covariates with fewer than five such variants were excluded from the adjustment set.

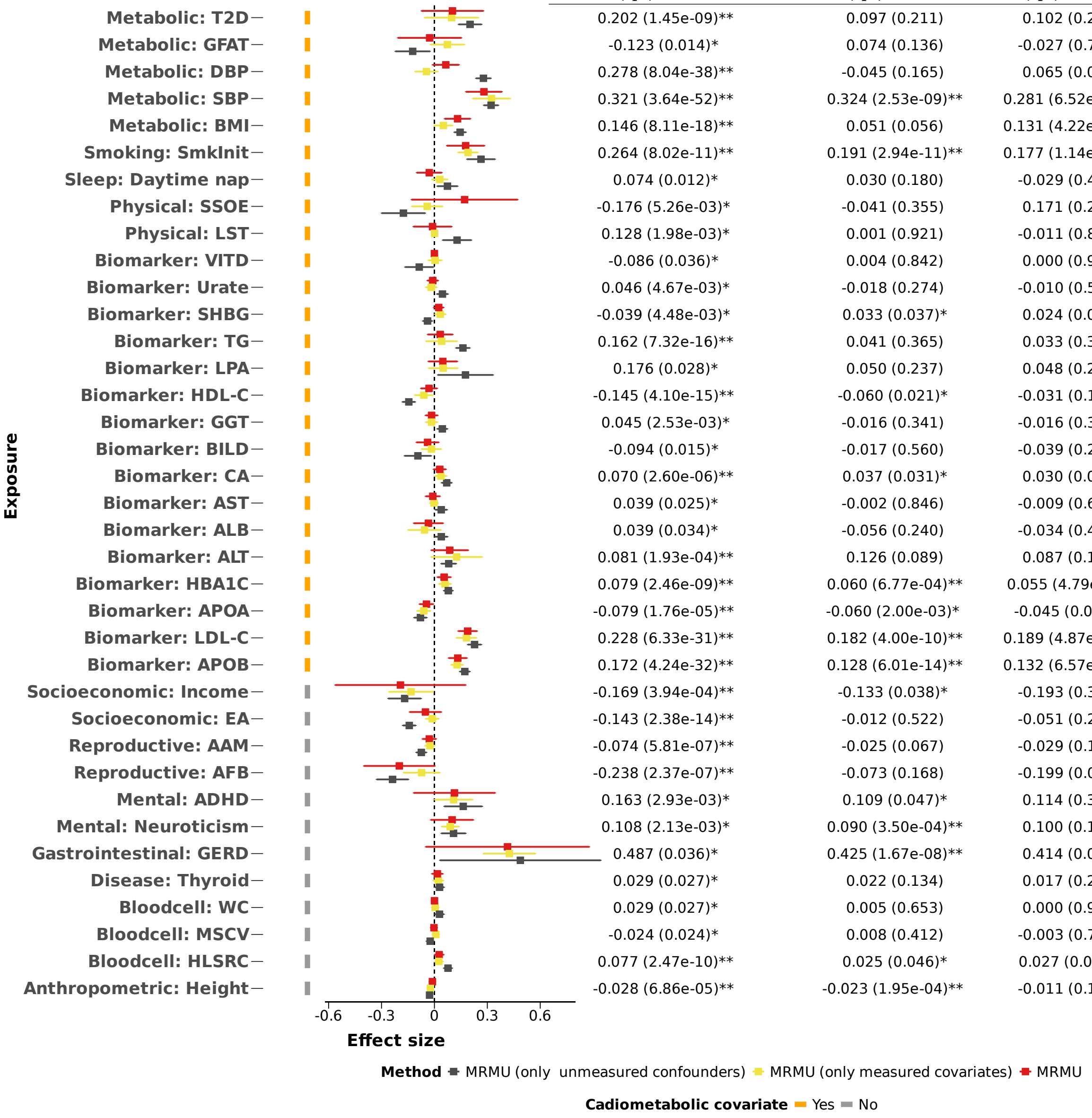

| Exposure | MRMU (only unmeasured confounders ) $\hat{\beta}_1$ (p-value) | MRMU (only measured covariates) $\hat{\beta}_1$ (p-value) | MRMU $\hat{\beta}_1$ (p-value) |
|---|---|---|---|
| Metabolic: T2D | 0.202 (1.45e-09)** | 0.097 (0.211) | 0.102 (0.252) |
| Metabolic: GFAT | -0.123 (0.014)* | 0.074 (0.136) | -0.027 (0.767) |
| Metabolic: DBP | 0.278 (8.04e-38)** | -0.045 (0.165) | 0.065 (0.091) |
| Metabolic: SBP | 0.321 (3.64e-52)** | 0.324 (2.53e-09)** | 0.281 (6.52e-08)** |
| Metabolic: BMI | 0.146 (8.11e-18)** | 0.051 (0.056) | 0.131 (4.22e-04)** |
| Smoking: SmkInit | 0.264 (8.02e-11)** | 0.191 (2.94e-11)** | 0.177 (1.14e-03)** |
| Sleep: Daytime nap | 0.074 (0.012)* | 0.030 (0.180) | -0.029 (0.422) |
| Physical: SSOE | -0.176 (5.26e-03)* | -0.041 (0.355) | 0.171 (0.265) |
| Physical: LST | 0.128 (1.98e-03)* | 0.001 (0.921) | -0.011 (0.848) |
| Biomarker: VITD | -0.086 (0.036)* | 0.004 (0.842) | 0.000 (0.985) |
| Biomarker: Urate | 0.046 (4.67e-03)* | -0.018 (0.274) | -0.010 (0.546) |
| Biomarker: SHBG | -0.039 (4.48e-03)* | 0.033 (0.037)* | 0.024 (0.090) |
| Biomarker: TG | 0.162 (7.32e-16)** | 0.041 (0.365) | 0.033 (0.364) |
| Biomarker: LPA | 0.176 (0.028)* | 0.050 (0.237) | 0.048 (0.256) |
| Biomarker: HDL-C | -0.145 (4.10e-15)** | -0.060 (0.021)* | -0.031 (0.195) |
| Biomarker: GGT | 0.045 (2.53e-03)* | -0.016 (0.341) | -0.016 (0.366) |
| Biomarker: BILD | -0.094 (0.015)* | -0.017 (0.560) | -0.039 (0.231) |
| Biomarker: CA | 0.070 (2.60e-06)** | 0.037 (0.031)* | 0.030 (0.095) |
| Biomarker: AST | 0.039 (0.025)* | -0.002 (0.846) | -0.009 (0.661) |
| Biomarker: ALB | 0.039 (0.034)* | -0.056 (0.240) | -0.034 (0.423) |
| Biomarker: ALT | 0.081 (1.93e-04)** | 0.126 (0.089) | 0.087 (0.105) |
| Biomarker: HBA1C | 0.079 (2.46e-09)** | 0.060 (6.77e-04)** | 0.055 (4.79e-03)* |
| Biomarker: APOA | -0.079 (1.76e-05)** | -0.060 (2.00e-03)* | -0.045 (0.026)* |
| Biomarker: LDL-C | 0.228 (6.33e-31)** | 0.182 (4.00e-10)** | 0.189 (4.87e-12)** |
| Biomarker: APOB | 0.172 (4.24e-32)** | 0.128 (6.01e-14)** | 0.132 (6.57e-07)** |
| Socioeconomic: Income | -0.169 (3.94e-04)** | -0.133 (0.038)* | -0.193 (0.307) |
| Socioeconomic: EA | -0.143 (2.38e-14)** | -0.012 (0.522) | -0.051 (0.266) |
| Reproductive: AAM | -0.074 (5.81e-07)** | -0.025 (0.067) | -0.029 (0.141) |
| Reproductive: AFB | -0.238 (2.37e-07)** | -0.073 (0.168) | -0.199 (0.052) |
| Mental: ADHD | 0.163 (2.93e-03)* | 0.109 (0.047)* | 0.114 (0.334) |
| Mental: Neuroticism | 0.108 (2.13e-03)* | 0.090 (3.50e-04)** | 0.100 (0.103) |
| Gastrointestinal: GERD | 0.487 (0.036)* | 0.425 (1.67e-08)** | 0.414 (0.080) |
| Disease: Thyroid | 0.029 (0.027)* | 0.022 (0.134) | 0.017 (0.293) |
| Bloodcell: WC | 0.029 (0.027)* | 0.005 (0.653) | 0.000 (0.933) |
| Bloodcell: MSCV | -0.024 (0.024)* | 0.008 (0.412) | -0.003 (0.768) |
| Bloodcell: HLSRC | 0.077 (2.47e-10)** | 0.025 (0.046)* | 0.027 (0.042)* |
| Anthropometric: Height | -0.028 (6.86e-05)** | -0.023 (1.95e-04)** | -0.011 (0.178) |

**Figure** S7: Comparison of MRMU model variants in the CAD risk factor analysis. Effect estimates are shown for three MRMU variants across the 37 prescreened candidate traits in the CAD (UKB) discovery dataset: adjustment for unmeasured confounders only, adjustment for measured cardiometabolic covariates only, and joint adjustment for both components. Points and horizontal bars show estimated effects and 95% confidence intervals, with effect estimates and p-values listed on the right. Orange and gray side bars denote cardiometabolic and non-cardiometabolic traits, respectively.$*$ denotes nominal significance ($P < 0.05$), and $**$ denotes significance after Bonferroni correction.

### Supplementary tables

**Table** S1: Instrument variable selection and LD clumping settings across MR methods.

| **Method** | **IV selection rule** | **P-value threshold** | **LD clumping** |
|---|---|---|---|
| MRMU/MR-APSS | Variants associated with the exposure | $P < 5 \times 10^{-5}$ | $r^2 < 0.01$ within a 1 Mb window |
| IVW/Egger/RAPS/Weighted-mode/Weighted-median | Variants associated with the exposure | $P < 5 \times 10^{-8}$ | $r^2 < 0.01$ within a 1 Mb window |
| CAUSE | Variants associated with the exposure | $P < 1 \times 10^{-3}$ | Recommended CAUSE clumping setting with $r^2 < 0.01$ |
| MVMR-IVW/MVMR-Egger/MVMR-Robust/MVMR-Lasso/MVMR-Median | Union of variants associated with any traits, except the outcome, included in the multivariable model | $P < 5 \times 10^{-8}$ | $r^2 < 0.01$ within a 1 Mb window |

**Table** S2: GWAS sources for traits in the negative control analysis.

| Trait role | Label in figures | Trait description | Sample size | PMID / Source | Consortium / GWAS catalog |
|---|---|---|---|---|---|
| exposure | Adult BMI | Adult body mass index, calculated as weight divided by height squared. | ∼700,000 | 30124842 | GIANT |
| covariate | Educational attainment | Years of schooling or highest educational level attained. | 765,283 | 35361970 | https://thessgac.com/papers/ |
| covariate | Household income | Self-reported total household income. | 286,301 | 31844048 | GCST009523 |
| covariate | Age at first sex | Self-reported age at first sexual intercourse. | 397,338 | 34211149 | GCST90000047 |
| covariate | Age at first birth | Age at which an individual had their first child. | 542,901 | 34211149 | GCST90000050 |
| covariate | Age at menarche | Self-reported age at first menstrual period. | ∼370,000 | 28436984 | http://www.reprogen.org/ |
| covariate | Age at voice break | Self-reported timing of voice breaking relative to peers. | 154,459 | UK Biobank / Neale Lab (UKB data field 2385) | https://broad-ukb-sumstats-us-east-1 s3.amazonaws.com/round2/additive-tsvs/2385.gwas.imputed_v3.male.tsv.bgz |
| covariate | Age of initiation of Smoking | Age at which an individual started smoking regularly. | 323,386 | 30643251 | AgeSmk https://doi.org/10.13020/przg-dp88 |
| covariate | Drinks per week | Average number of alcoholic drinks consumed per week. | 941,280 | 30643251 | DrnkWk https://doi.org/10.13020/przg-dp88 |
| covariate | Number of sexual partners | Self-reported lifetime number of sexual partners. | ∼378,882 | 30643258 | https://thessgac.com/papers/ |
| covariate | Diet: fat | Proportion of total energy intake derived from fat. | ∼235,000 | 32393786 | https://thessgac.com/papers/ |
| covariate | Diet: protein | Proportion of total energy intake derived from protein. | ∼235,000 | 32393786 | https://thessgac.com/papers/ |
| covariate | Diet: carbs | Proportion of total energy intake derived from carbohydrates. | ∼235,000 | 32393786 | https://thessgac.com/papers/ |
| covariate | Diet: sugar | Proportion of total energy intake derived from sugar. | ∼235,000 | 32393786 | https://thessgac.com/papers/ |
| outcome | Height at age 10 | Self-reported height at age 10 relative to peers. | 355,331 | UK Biobank / Neale Lab (UKB data field 1697) | https://broad-ukb-sumstats-us-east-1 s3.amazonaws.com/round2/additive-tsvs/1697.gwas.imputed_v3.both_sexes.tsv.bgz |

**Table** S3: GWAS sources for candidate traits used in the CAD real-data analysis.

| **Category** | **Label in figures** | **Trait** | **Sample size** | **PMID** |
|---|---|---|---|---|
| Anthropometric trait | Anthropometric: Height | Height | ∼700,000 | 30124842 |
| Autoimmune disease | Autoimmune: Asthma | Asthma (Ast) | 393,859 | 32296059 |
| Autoimmune disease | Autoimmune: Ecz | Atopic dermatitis/ eczema (Ecz) | 796,661 | 34454985 |
| Autoimmune disease | Autoimmune: RA | Rheumatoid arthritis | 97,173 | 36333501 |
| Autoimmune disease | Autoimmune: T1D | Type 1 diabetes | 520,580 | 34012112 |
| Biomarker | Biomarker: ALB | Albumin | 331,979 | 33462484 |
| Biomarker | Biomarker: ALP | Alkaline phosphatase | 363,228 | 33462484 |
| Biomarker | Biomarker: ALT | Alanine aminotransferase | 363,054 | 33462484 |
| Biomarker | Biomarker: APOA | Apolipoprotein A | 330,515 | 33462484 |
| Biomarker | Biomarker: APOB | Apolipoprotein B | 361,372 | 33462484 |
| Biomarker | Biomarker: AST | Aspartate aminotransferase | 361,854 | 33462484 |
| Biomarker | Biomarker: BILD | Direct bilirubin | 308,695 | 33462484 |
| Biomarker | Biomarker: BUN | Urea | 362,973 | 33462484 |
| Biomarker | Biomarker: CA | Calcium | 332,355 | 33462484 |
| Biomarker | Biomarker: CRE | Creatinine | 363,070 | 33462484 |
| Biomarker | Biomarker: CRP | C-reactive protein | 575,531 | 35459240 |
| Biomarker | Biomarker: CYS | Cystatin C | 363,090 | 33462484 |
| Biomarker | Biomarker: EGFR | eGFR | 363,070 | 33462484 |
| Biomarker | Biomarker: GGT | Gamma glutamyltransferase | 363,026 | 33462484 |
| Biomarker | Biomarker: HBA1C | HbA1c | 345,814 | 33462484 |
| Biomarker | Biomarker: HDL-C | HDL cholesterol | 332,323 | 33462484 |
| Biomarker | Biomarker: IGF1 | Igf-1 | 361,107 | 33462484 |
| Biomarker | Biomarker: LDL-C | LDL cholesterol | 362,516 | 33462484 |
| Biomarker | Biomarker: LPA | Lipoprotein A | 290,497 | 33462484 |
| Biomarker | Biomarker: NAP | Non-albumin protein | 331,979 | 33462484 |
| Biomarker | Biomarker: PHOS | Phosphate | 331,826 | 33462484 |
| Biomarker | Biomarker: SHBG | Shbg | 329,122 | 33462484 |
| Biomarker | Biomarker: TBIL | Total bilirubin | 361,681 | 33462484 |
| Biomarker | Biomarker: TES | Testosterone | 329,274 | 33462484 |
| Biomarker | Biomarker: TG | Triglycerides | 362,907 | 33462484 |
| Biomarker | Biomarker: TP | Total protein | 331,979 | 33462484 |
| Biomarker | Biomarker: UCR | Creatinine in urine | 352,922 | 33462484 |
| Biomarker | Biomarker: URK | Potassium in urine | 352,180 | 33462484 |
| Biomarker | Biomarker: URNA | Sodium in urine | 352,164 | 33462484 |
| Biomarker | Biomarker: Urate | Urate | 362,754 | 33462484 |
| Biomarker | Biomarker: VITD | Vitamin D | 346,130 | 33462484 |
| Blood cell | Bloodcell: EC | Eosinophil count | 459,000 | 29892013 |
| Blood cell | Bloodcell: HLSRC | High light scatter reticulocyte count | 459,000 | 29892013 |
| Blood cell | Bloodcell: LC | Lymphocyte count | 459,000 | 29892013 |
| Blood cell | Bloodcell: MC | Monocyte count | 459,000 | 29892013 |
| Blood cell | Bloodcell: MCH | Mean corpuscular hemoglobin | 459,000 | 29892013 |
| Blood cell | Bloodcell: MPV | Mean platelet volume | 459,000 | 29892013 |
| Blood cell | Bloodcell: MSCV | Mean sphered cell volume | 459,000 | 29892013 |
| Blood cell | Bloodcell: PC | Platelet count | 459,000 | 29892013 |
| Blood cell | Bloodcell: PDW | Platelet distribution width | 459,000 | 29892013 |
| Blood cell | Bloodcell: RC | Red blood cell count | 459,000 | 29892013 |
| Blood cell | Bloodcell: RDW | RBC distribution width | 459,000 | 29892013 |
| Blood cell | Bloodcell: WC | White blood cell count | 459,000 | 29892013 |
| Cancer | Breast cancer | Breast cancer | 13,778/183,466 | 37340002 |
| Cancer | Colorectal cancer | Colorectal cancer | 7,194/334,343 | 37340002 |
| Cancer | Pan cancer | Meta-analysis of all cancer types | 43,098/334,343 | 37340002 |

*Continued on next page*

*Continued from previous page*

| **Category** | **Label** | **Trait** | **Sample size** | **PMID** |
|---|---|---|---|---|
| Cancer | Prostate cancer | Prostate cancer | 10,739/150,877 | 37340002 |
| Disease | Disease: Thyroid | thyroid-stimulating hormone | 247,107 | 37872160 |
| Gastrointestinal disease | Gastrointestinal: CD | Crohn's disease | 40,266 | 28067908 |
| Gastrointestinal disease | Gastrointestinal: GERD | Gastroesophageal reflux disease | 385,276 | 31527586 |
| Gastrointestinal disease | Gastrointestinal: IBD | Inflammatory bowel disease | 59,957 | 28067908 |
| Gastrointestinal disease | Gastrointestinal: UC | Ulcerative colitis | 45,975 | 28067908 |
| Lifestyle (Physical activity) | Physical: AA | Average acceleration | 91,084 | 29899525 |
| Lifestyle (Physical activity) | Physical: Computer | Leisure computer use | 408,815 | 32317632 |
| Lifestyle (Physical activity) | Physical: LST | Leisure screen time | 526,725 | 36071172 |
| Lifestyle (Physical activity) | Physical: SSOE | Strenuous sports or other exercises: $\geq$2–3 vs. 0 days/week | 238,132 | 29899525 |
| Lifestyle (Physical activity) | Physical: VPA | VPA: $\geq$3 vs. 0 days/week | 261,055 | 29899525 |
| Lifestyle (Sleep behavior) | Sleep: Chronotype | Chronotype | 449,734 | 30696823 |
| Lifestyle (Sleep behavior) | Sleep: Daytime nap | Daytime napping | 452,633 | 33568662 |
| Lifestyle (Sleep behavior) | Sleep: Daytime sleepiness | Daytime sleepiness | 452,071 | 31409809 |
| Lifestyle (Sleep behavior) | Sleep: Insomnia | Insomnia | 453,379 | 30804566 |
| Lifestyle (Sleep behavior) | Sleep: Morning person | Morning person | 403,195 | 30696823 |
| Lifestyle (Sleep behavior) | Sleep: Sleep Episodes | Number of sleep episodes | 84,810 | 30846698 |
| Lifestyle (Smoking and alcohol drinking) | Alcohol: DrnkWk | Drinks per week | 941,280 | 30643251 |
| Lifestyle (Smoking and alcohol drinking) | Alcohol: PAU | Problematic alcohol use | 903,147 | 38062264 |
| Lifestyle (Smoking and alcohol drinking) | Smoking: AgeSmk | Age at initiation of regular smoking | 341,427 | 30643252 |
| Lifestyle (Smoking and alcohol drinking) | Smoking: CigDay | Heaviness of smoking | 337,334 | 30643254 |
| Lifestyle (Smoking and alcohol drinking) | Smoking: SmkInit | Ever smoked regularly | 1,232,091 | 30643253 |
| Mental health and brain development | Mental: ADHD | Attention-Deficit/Hyperactivity Disorder | 225,534 | 36702997 |
| Mental health and brain development | Mental: Cerebellar volume | Cerebellar volume | 27,486 | 35842455 |
| Mental health and brain development | Mental: Neuroticism | Neuroticism | 682,688 | 38293137 |
| Metabolic trait | Metabolic: BMI | Body mass index | $\sim$700,000 | 30124842 |
| Metabolic trait | Metabolic: DBP | Diastolic blood pressure | 1,028,980 | 38689001 |
| Metabolic trait | Metabolic: GFAT | Gluteofemoral | 38,965 | 35773277 |
| Metabolic trait | Metabolic: SBP | Systolic blood pressure | 1,028,980 | 38689001 |
| Metabolic trait | Metabolic: T2D | Type 2 diabetes | 933,970 | 35551307 |
| Reproductive traits | Reproductive: AAM | Age at menarche | $\sim$370,000 | 28436984 |
| Reproductive traits | Reproductive: AFB | Age at first birth | 542,901 | 34211149 |
| Reproductive traits | Reproductive: ANM | Age at menopause | $\sim$200,000 | 34349265 |
| Reproductive traits | Reproductive: AVB | Relative age voice broke | 154,459 | 34662886 |
| Socioeconomic status | Socioeconomic: EA | Educational attainment | 765,283 | 35361970 |
| Socioeconomic status | Socioeconomic: Income | Household income | 497,413 | 39875632 |
| Socioeconomic status | Socioeconomic: Intelligence | Intelligence | 269,867 | 29942086 |
